# Genome-wide association study of borderline personality disorder identifies 11 loci and highlights shared risk with mental and somatic disorders

**DOI:** 10.1101/2024.11.12.24316957

**Authors:** Fabian Streit, Swapnil Awasthi, Alisha SM Hall, Alice Braun, Maria Niarchou, Eirini Marouli, Oladapo Babajide, Josef Frank, Lea Zillich, Carolin M Callies, Diana Avetyan, Eric Zillich, Joonas Naamanka, Jean Gonzalez, Arvid Harder, Yi Lu, Zouhair Aherrahrou, Zain-Ul-Abideen Ahmad, Helga Ask, Anthony Batzler, Michael E Benros, Odette M Brand-de Wilde, Søren Brunak, Mie T Bruun, Lea AN Christoffersen, Lucía Colodro-Conde, Brandon J Coombes, Elizabeth C Corfield, Norbert Dahmen, Maria Didriksen, Khoa M Dinh, Srdjan Djurovic, Joseph Dowsett, Ole Kristian Drange, Helene Dukal, Susanne Edelmann, Christian Erikstrup, Mariana K Espinola, Eva Fassbinder, Annika Faucon, Diana S Ferreira de Sá, Jerome C Foo, Maria Gilles, Alfonso Gutiérrez-Zotes, Thomas F Hansen, Magnus Haraldsson, R. Patrick Harper, Alexandra Havdahl, Urs Heilbronner, Stefan Herms, Henrik Hjalgrim, Christopher Hübel, Gitta A Jacob, Bitten Aagaard, Anders Jorgensen, Martin Jungkunz, Nikolaus Kleindienst, Nora Knoblich, Stefanie Koglin, Julia Kraft, Kristi Krebs, Christopher W Lee, Yuhao Lin, Stefanie Lis, Amanda Lisoway, Ioannis A Malogiannis, Amy Martinsen, Tolou Maslahati, Katharina Merz, Andreas Meyer-Lindenberg, Susan Mikkelsen, Christina Mikkelsen, Arian Mobascher, Gerard Muntané, Asmundur Oddsson, Sisse R Ostrowski, Teemu Palviainen, Ole BV Pedersen, Geir Pedersen, Liam Quinn, Matthias A Reinhard, Florian A Ruths, Björn H Schott, Michael Schredl, Emanuel Schwarz, Cornelia E Schwarze, Michael Schwinn, Tabea Send, Engilbert Sigurdsson, Katja Simon-Keller, Astros T Skuladottir, Joaquim Soler, Anne Sonley, Erik Sørensen, Hreinn Stefansson, Peter Straub, Jaana Suvisaari, Martin Tesli, Jacob Træholt, Henrik Ullum, Maja P Völker, G Bragi Walters, Rujia Wang, Christian C Witt, Gerhard Zarbock, Peter Zill, John-Anker Zwart, DBDS Genomic Consortium, Estonian Biobank Research Team, the GLAD Study, HUNT All-In Psychiatry, Ole A Andreassen, Arnoud Arntz, Joanna M Biernacka, Martin Bohus, Gerome Breen, Alexander L Chapman, Sven Cichon, Lea K Davis, Michael Deuschle, Sebastian Euler, Sabine C Herpertz, Benjamin Hummelen, Andrea Jobst, Jaakko Kaprio, James L Kennedy, Kelli Lehto, Klaus Lieb, Lourdes Martorell, Shelley McMain, Richard Musil, Vanessa Nieratschker, Markus M Nöthen, Frank Padberg, Aarno Palotie, Juan C Pascual, Nader Perroud, Josep A Ramos-Quiroga, Ted Reichborn-Kjennerud, Marta Ribases, Stefan Roepke, Dan Rujescu, Sandra Sanchez-Roige, Claudia Schilling, Christian Schmahl, Kari Stefansson, Thorgeir E Thorgeirsson, Gustavo Turecki, Elisabet Vilella, Thomas Werge, Bendik S Winsvold, Johannes Wrege, Marcella Rietschel, Stephan Ripke, Stephanie H Witt

## Abstract

We conducted the largest genome-wide meta-analysis of borderline personality disorder (BPD) to date, with a discovery sample of 12,339 cases and 1,041,717 controls, and a replication study of 685 cases and 107,750 controls (all participants of European ancestry). We identified 11 independent associated genomic loci, and nine risk genes in the gene-based analysis. We observed a single-nucleotide polymorphism (SNP) heritability of 17.3% and derived polygenic scores (PGS) predicted 4.6% of the phenotypic variance in BPD on the liability scale. BPD showed the strongest positive genetic correlations with GWAS of posttraumatic stress disorder, depression, attention deficit hyperactivity disorder, antisocial behavior, and measures of suicide and self-harm. Phenome-wide association analyses using BPD-PGS confirmed these associations and additionally revealed associations with general medical conditions including obstructive pulmonary disease and diabetes. The present analyses highlight BPD as a polygenic disorder, with the genetic risk showing substantial overlap with psychiatric and physical health conditions.

## Main text

Borderline Personality Disorder (BPD) is a mental disorder characterized by pervasive instability in emotions, interpersonal relationships, and self-image, and by impulsive behavior (symptoms see Supplementary Methods)^1,2^. BPD has a prevalence of 0.92−1.90% in Western countries^3^, with symptom onset typically occurring during adolescence. Women are more frequently diagnosed with BPD than men by a ratio of ∼3:1, for which a substantial contribution of diagnostic as well as selection bias has been postulated^4,5^. Individuals with BPD display high rates of self-harm, suicidal ideation, and suicide attempts. BPD shows substantial symptom overlap and comorbidity with other mental disorders^5,6^, and comorbidity with neurological and somatic health conditions^6,7^. While some psychotherapies are effective in treating BPD^8^, no psychopharmacological treatments are FDA□approved specifically for BPD^2,9^.

In addition to environmental risk factors such as early interpersonal trauma^10,11^, genetic factors substantially contribute to disorder risk. Twin and family studies estimate the heritability of BPD to be 46−69%^12,13^, and demonstrate that the genetic risk for BPD is partially shared with other mental disorders, but as well as with continuous traits, e.g. the Big Five personality traits^14,15^. However, a systematic assessment of shared genetic risk with a broad range of disorders and traits is missing.

For many mental disorders, genome-wide association studies (GWAS) meta-analyzing genetic data from tens or hundreds of thousands of cases and controls have successfully identified up to hundreds of genetic risk loci^16^. In contrast, genetic research on BPD lags behind^17^: In the thus far only GWAS of BPD conducted in 998 cases and 1,545 controls^18^, no single genome-wide significant variants were identified, but significant genetic correlations (*r*_*g*_) of BPD were observed with bipolar disorder (BIP), schizophrenia (SCZ), and major depressive disorder^18^. While these findings indicate the potential of using genetic approaches to investigate BPD, research based on those results is limited by the large uncertainties in the estimated effect sizes. Additionally, the extent to which sex-specific genetic effects contribute to the observed sex differences in the prevalence and clinical characteristics of BPD remains unclear.

The main aims of the present study were to a) identify novel genetic risk loci for BPD to improve the understanding of the underlying molecular mechanisms, and b) systematically assess the shared genetic risk between BPD and a broad range of related traits and disorders.

## Results

### Genome-wide association study

We performed a discovery GWAS meta-analysis including 6,043,895 genetic markers in 12,339 BPD cases and 1,041,717 controls of European ancestry (overview Fig. 1). In total, 17 studies contributed individual-level data for a total of 2,705 cases meeting DSM-IV criteria for BPD and 4,600 controls, which were combined into five datasets. Additionally, 10 large-scale biobank or cohort studies provided summary statistics from GWAS of BPD (*N*_cases_=9,634, *N*_controls_=1,037,117 controls). In these, BPD status was assessed using ICD codes except for the Genetic Links to Anxiety and Depression (GLAD) study, which used a self-reported diagnosis. Details on ascertainment, and inclusion and exclusion criteria are documented in Supplementary Table S1. Sample sizes, basic demographics (sex and age), depression comorbidity, and analysis details are provided in Supplementary Table S2 and the Supplementary Methods. Additionally, we performed meta-analyses stratified by sex (female-only; male-only) as well as a sensitivity analysis excluding subjects with a history of BIP or SCZ—conditions commonly excluded in BPD studies—from the studies providing summary statistics (Supplementary Tables S3−S5). To assess replication, the lead SNPs and PGS derived from the discovery analysis were tested in two independent datasets (total N_cases_=685, total N_controls_=107,750). Additionally, SNPs with p<1×10^−6^ in the discovery GWAS were analyzed in a combined meta-analysis (*N*_*cases*_=13,024, *N*_*controls*_=1,149,467).

**Figure 1:**
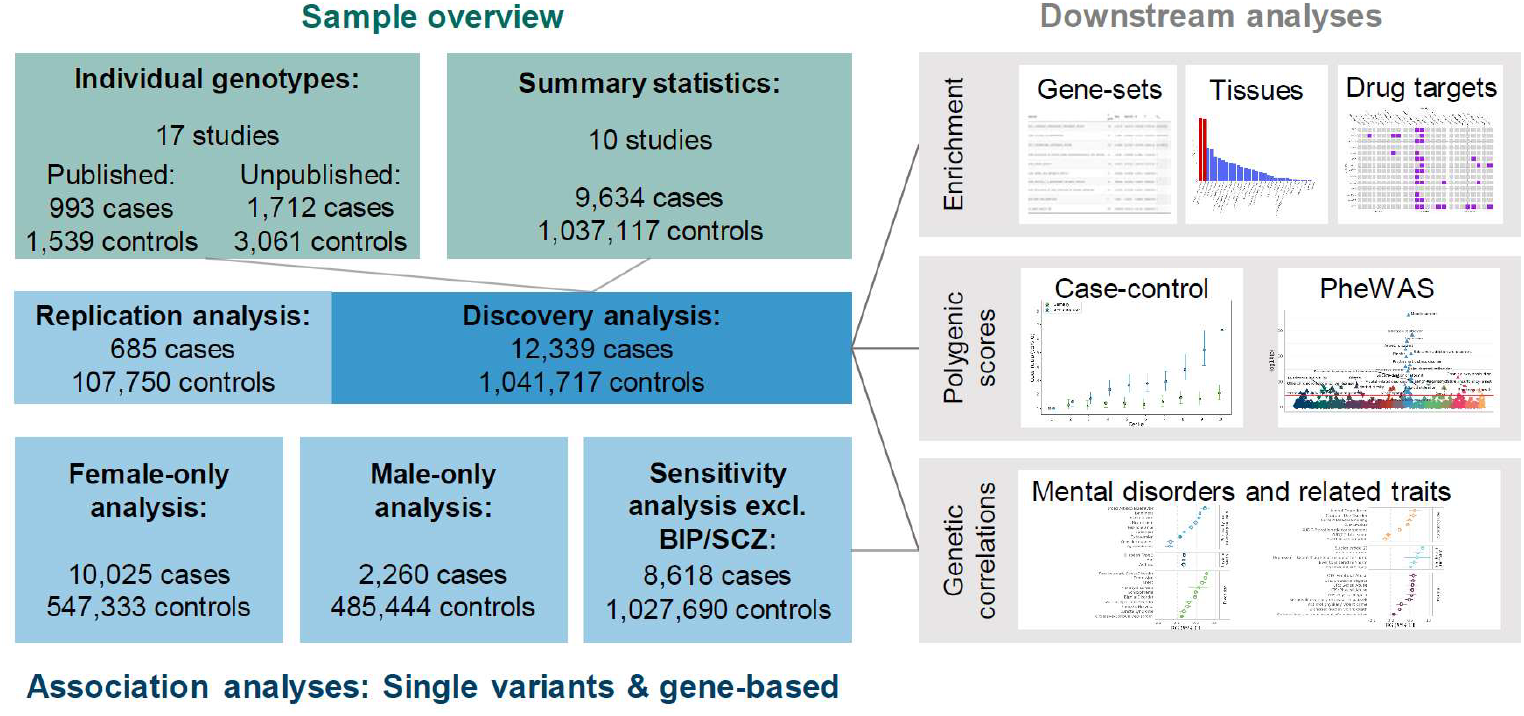
Schematic overview of the genome-wide association study (GWAS) of borderline personality disorder (BPD). A total of 17 studies providing individual-level data (analyzed in 5 datasets: see text) and 10 studies providing summary statistics were included in the discovery meta-analysis. Genetic associations were tested at the single-variant and gene level. Gene enrichment analyses were applied to test for enrichment in gene-sets and tissue-specific gene expression data from 53 human tissues (GTEx), from 29 different ages and 11 general developmental stages (BrainSpan), single-cell based cell types, and drug-target gene set. Polygenic scores were calculated to assess the prediction of BPD case-control status in the analysis datasets with available individual-level data and to test the association of the genetic liability for BPD in phenome-wide association studies (PheWAS) with phecodes in two biobanks (BioVU, UK Biobank). Genetic correlations were calculated between the results of the GWAS of BPD and the GWAS of 50 disorders and traits of interest. BIP=bipolar disorder; SCZ=schizophrenia.

The discovery GWAS had power over 80% to detect effects of 1.1 for variants with an allele frequency between 0.3−0.7^19^ (Supplementary Table S6, Supplementary Figure S1).

We observed an SNP-heritability of 28.4% (95% confidence interval (CI) [24.9%, 31.8%]), corresponding to an SNP-heritability of 17.2% (95%-CI [15.2%, 19.4%]) on the liability scale (assuming a population prevalence of 1.5%)^3^. The linkage disequilibrium score regression (LDSC)^20^ intercept was 1.03 (standard error (SE)=0.0085), with an attenuation ratio of 0.11 (SE=0.031), and we observed a genomic inflation factor (λ_GC_) of 1.22 (λ_1000_=1.01). There was a high genetic correlation between the studies with individual-level data and those providing summary statistics (*r*_*g*_=0.85, 95%-CI [0.68, 1.03]), which was significantly smaller than 1 (*p*=0.038).

SNP association analysis revealed six independent genome-wide significant loci (*p*<5×10^−8^, Fig. 2; Table 1; Supplementary Figs. S2−S14; Supplementary Table S7), and no marker showed significant heterogeneity between studies after multiple testing correction (smallest heterogeneity p-value=0.023). All six genome-wide significant lead SNPs (6/6, *p*=0.016, Tables 1), and 77% of the lead SNPs associated with *p*<1×10^−6^ (24/31, *p*=0.0017) showed effects in the same direction in the replication analysis. In the combined analysis, the six loci remained genome-wide significant, and five additional loci with p<5×10^−8^ were observed (Figure1, Table 1, Supplementary Figs. S2−S24).

**Table 1:**
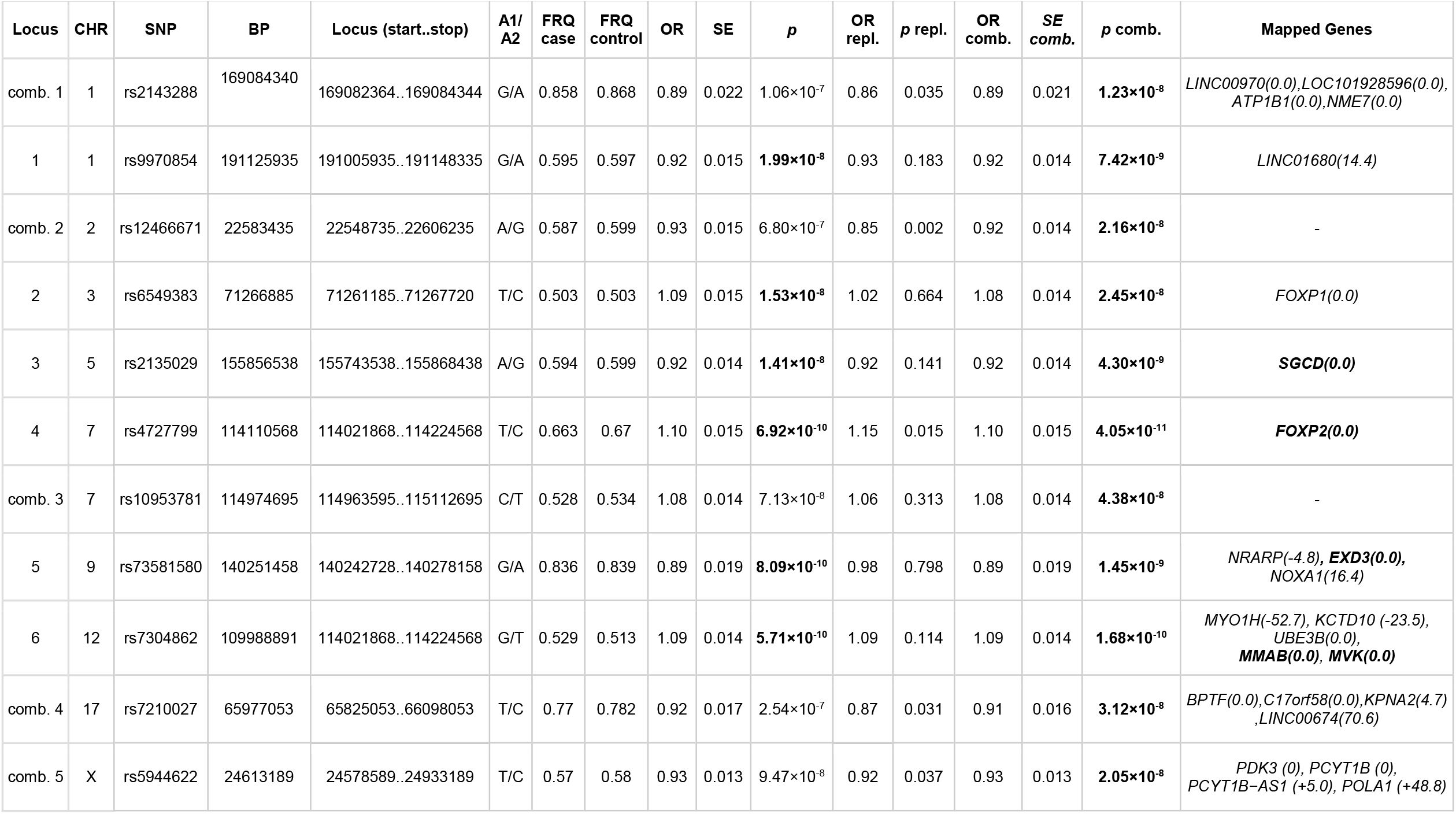
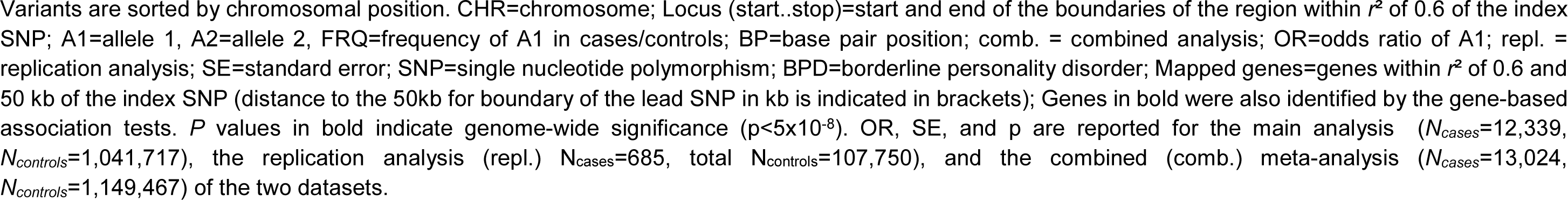
Lead 11 genome-wide significant SNPs associated (p<5×10^−8^) with BPD.

**Figure 2:**
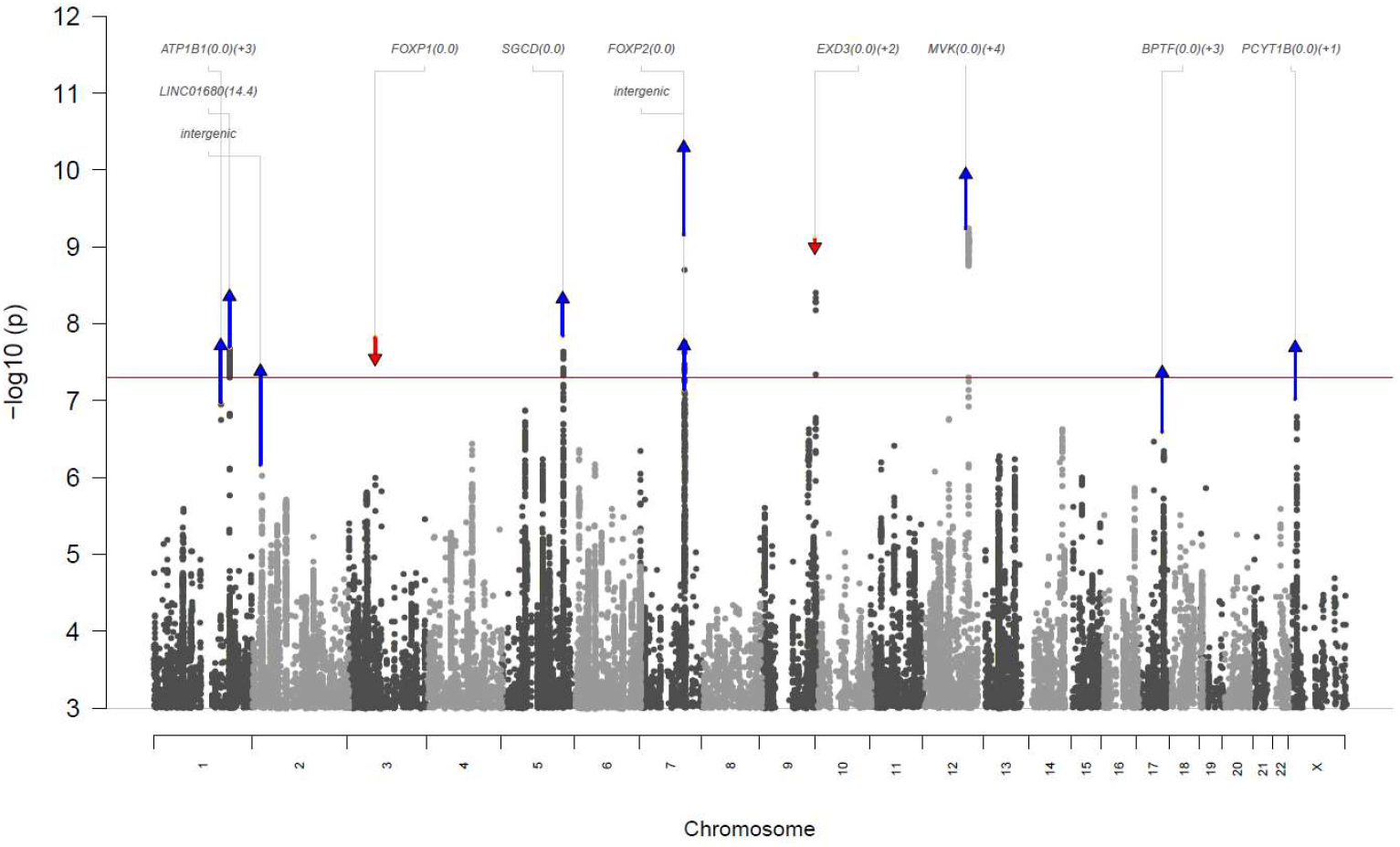
Manhattan plot of the genome-wide association study (GWAS) of borderline personality disorder (BPD). The -log10 *p-*value for each single nucleotide polymorphism (SNP) in the discovery GWAS (*N*_*cases*_=12,339, *N*_*controls*_=1,041,717) is indicated on the y-axis (chromosomal position shown on the x-axis). The red line horizontal indicates genome-wide significance (*p*<5×10^−8^). Index SNPs representing independent genome-wide significant associations either in the discovery GWAS or the combined analysis (*N*_*cases*_=13,024, *N*_*controls*_=1,149,467) are highlighted (upward-pointing blue triangle: increased significance in combined analysis, downward-pointing red triangle: reduced significance in combined analysis). Combined p-values are plotted for lead loci and vertical lines indicate the p-value change from the discovery to the combined analysis.

Gene-based associations using MAGMA (version 1.07)^21,22^ indicated nine genes, including *CCDC71* and *DEPDC1B*; in addition to *SGCD, FOXP2, EXD3, MVK, MMAB, PCYT1B*, and *BPTF* already indicated by the genome-wide SNP associations (Extended Data Fig. 1; Supplementary Fig. S25; Supplementary Table S8).

Among the genes implicated by proximity to lead SNPs or by the gene-based analysis, the highest Polygenic Priority Scores (PoPS)^23^ were observed for *FOXP2* (score=0.96, rank=1), and *SGCD* (score=0.74, rank=2), with *FOXP1* and *DEPDC1B* also ranking in the top 1% (Supplementary Table S9). Of the genes in proximity to loci that reached significance in the combined analysis, *NME7* and *KPNA2* ranked highest (top 5%).

### Sex-stratified GWAS

We carried out sex-stratified GWAS meta-analyses for male (*N*_cases_=2,260, *N*_controls_=485,444) and female subjects (*N*_cases_=10,025, *N*_controls_=547,333) (Supplementary Tables S3&S4).

We used sex-specific population prevalences of 2.25% for females and 0.75% for males to convert heritability estimates to the liability scale^4,5^. For females, we observed an SNP-heritability of 30.3% (95%-CI [26.1%, 34.5%]), corresponding to 20.5% (95%-CI [17.7%, 23,4%]) on the liability scale. For males, the observed SNP-heritability was 19,8% (95%-CI [7.9%, 31.6%]), corresponding to 10.2% (95%-CI [4.1%, 16.3%] on the liability scale, which was significantly lower than in females (*p*=0.00090).

There was a high genetic correlation between the two analyses (*r*_*g*_=0.80, 95%-CI [0.53, 1.07]), which did not differ from 1 (*p*=0.055).

The female-only analysis identified one genome-wide significant risk locus on chromosome 9 (rs73581580; *p*=2.83×10^−8^; locus 5 identified in the main analysis (*EXD3*), and a second locus on chromosome 7 (rs10227454, *p*=4.99×10^−8^), ∼1 Mb from locus 4 (*FOXP2, R2*=0.005, *D’*=0.25; Extended Data Fig. 2; Supplementary Figs. S26−S30, Supplementary Table S10). The gene-based analysis identified genome-wide associations for *DEPDC1B, SGCD, MVK*, and *MMAB*, all significant in the main analysis (Extended Data Fig. 3; Supplementary Fig. S31).

The male-only analysis identified two genome-wide significant risk loci that were not genome-wide significant in the main analysis: one on chromosome 2 (rs17757829, *p*=1.02×10^−8^) and one on chromosome 20 (rs6032676, *p*=9.55×10^−9^) (Extended Data Fig. 4; Supplementary Figs. S32−S36, Supplementary Table S11) and no significant genes (Extended Data Fig. 5; Supplementary Fig. S37).

The six lead SNPs of the main GWAS showed comparable effects and at least nominal significance in the smaller, and therefore lower-powered, sex-stratified analyses (Supplementary Table S12; Extended Data Fig. 6). The nine genes significant in the gene-based analysis of the main GWAS all showed nominal significance in the female-only analysis(all *p*_female_<8.15×10^−5^), whereas *CCDC71, DEPDC1B*, and *SGCD* were not significant in the male-only analysis (Supplementary Table S13).

### Sensitivity analysis excluding BIP&SCZ

In the sensitivity analysis excluding subjects with schizophrenia and bipolar disorder (*N*_cases_=8,618, *N*_controls_=1,027,690), locus 4 in *FOXP2* (rs4727799) and locus 5 in *EXD3* (rs73581580) from the main analysis were the only genome-wide significant associations (Extended Data Fig. 7; Supplementary Figs. S38−S42, Supplementary Table S14). All six lead SNPs of the main GWAS (all *p*_sensitivity_<3.32×10^−5^; Extended Data Fig. 6; Supplementary Table S12). The nine significant genes in the gene-based analysis of the main analysis, all were nominally significant in the sensitivity analysis (*p*_sensitivity_<0.0021; Supplementary Table S13) with *FOXP2, EXD3, MMAB, MVK* reaching genome-wide significance, and *ZNF626* being the only additional genome-wide significant gene (Extended Data Fig. 8; Supplementary Fig. S43).

### Enrichment analysis

Enrichment of gene-sets and tissue expression using Genotype-Tissue Expression (GTEx; version 8) data for 53 tissue types^24^ and expression data from 29 different ages and 11 general developmental stages (BrainSpan)^25^ was tested with MAGMA (version 1.07)^21^ as implemented in FUMA^22^: One significant gene-set was identified: the S1P-S1P3 (Sphingosine-1-phosphate - sphingosine-1-phosphate receptor 3) pathway^26^ (*p*_*adj*_=0.018; Supplementary Table S15). Nominally significant enrichment was observed in several GTEx tissue types, with the most significant enrichment in cerebellar tissue (not significant after correction for multiple testing; all *p*_*adj*_>0.32; Supplementary Fig. S44, Supplementary Table S16). The analysis using the two BrainSpan data sets indicated the strongest enrichments 8−21 weeks post conception (not significant after correction; all *p*_*adj*_>0.09), and in early to mid prenatal phases respectively (*p*_*adj*_<0.025; Supplementary Fig. S45&46, Supplementary Table S17&18).

Analyses based on Human Brain Atlas single-nucleus RNA sequencing data^27^, showed SNP-h2 enrichment using stratified LDSC^28,29^ in 2 of the 31 tested superclusters (*“medium spiny neurons”*;*”LAMP5-LHX6 and Chandelier cells”*; *p*_*adj*_<0.0064; Supplementary Fig. S47, Supplementary Table S19).

### Drug target analysis

Of the 16 genes priotized from the discovery GWAS meta-analysis via proximity to the 6 GWAS loci or in the gene-based test, none were highlighted as drug targets by Open Targets, but several showed potential tractability (Supplementary Fig. S48). The Genome for REPositioning drugs (GREP) pipeline revealed no significant enrichment for drug targets across any ICD or ATC category. The Drug Gene Interaction Database (DGIdb) highlighted drug-gene interactions for *MYO1H* with lithium, *MVP* with ALENDRONATE SODIUM, and *PCYT1B* with the unapproved substances CT-2584 and SPHINGOSINE. In an additional analysis with DRUGSETS^30^, based on MAGMA, testing the enrichment of the GWAS associations in 735 drug−gene sets, the most significant drugs included medications for neurological conditions (safinamide, gabapentin, lomerizine, oxcarbazepine), pain (prilocaine, tetracaine), and alcohol-dependence (acamprosate), but also medications for metabolic/somatic conditions (Supplementary Table S20; all *p*_*adj*_>0.18).

### Polygenic scoring

BPD-PGS were calculated using PRS-CS^31^ in the datasets where individual-level genotype information was available using leave-one-out summary statistics, excluding the respective sample from the discovery GWAS. In addition, we calculated PGS in the two independent replication data sets (Online Methods). For comparison, PGS were additonally calculated based on the first BPD GWAS from 2017^18^. The explained variance was converted to the liability scale (population prevalence = 1.5%^3^).

PGS explained a weighted average of 4.6% (AUC=66.0%) of the phenotypic variance on the liability scale (assuming a lifetime prevalence of 1.5%^3^) in the five datasets included in the discovery meta-analysis (Fig. 3, Supplementary Table S21). In the independent replication samples, PGS explained 3.1% of the variance (*p*=0.0041, AUC=62.4%) in Spain 2, and 2.6% in All of Us (*p*=2.09×10^−25^, AUC=61.4%). The odds ratio for BPD case-control status comparing the highest PGS decile to the lowest decile was OR=6.61 (95%-CI [5.20, 8.41]) for PGS based on the current meta-analysis, compared to OR=2.09 (95%-CI [1.60, 2.72]) for PGS based on the 2017 GWAS. When comparing to the middle 10% of the distribution, we observed an OR of 2.37 (95%-CI [1.94, 2.89])) for the highest decile for PGS based on the current meta-analysis and an OR of 1.44 (95%-CI [1.05, 1.98]) for PGS based on the 2017 GWAS.

**Figure 3:**
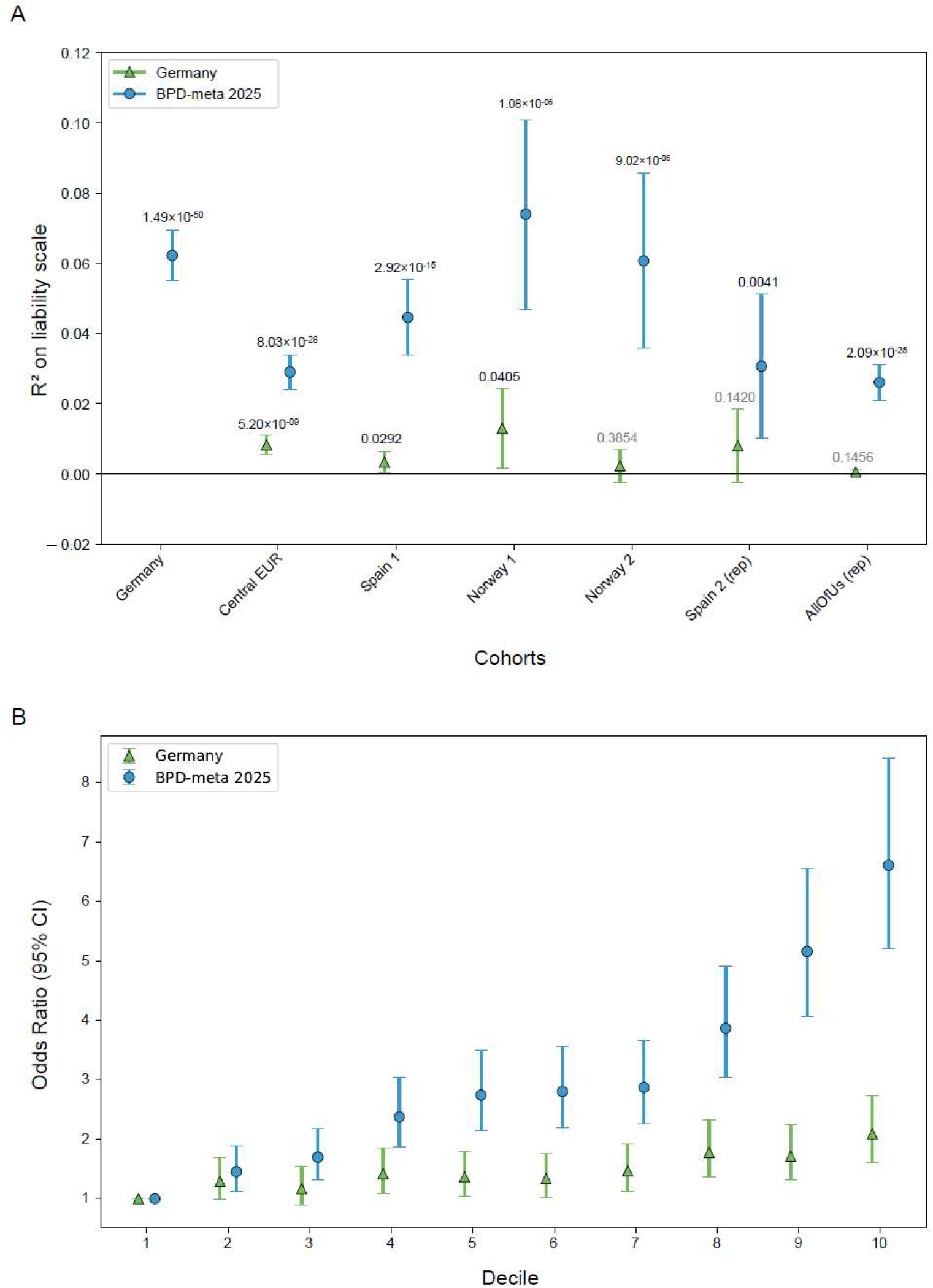
Polygenic score (PGS) analysis. A) The proportion of variance in case-control status explained by the PGS on the liability scale (y-axis; Liability *R2*) B) Odds ratio for BPD by PGS deciles, with decile 1 as reference. Leave-one-out PGS were calculated for the datasets where individual-level genotype data was available, and in an additional independent PGS target sample (Spain2 (rep.), sample information in Supplementary Tables S1&S2) using PRS-CS. For each prediction, the respective dataset was excluded from the used discovery genome-wide association study (GWAS) meta-analysis. PGS in the independent target dataset were based on the complete current GWAS meta-analysis (blue circles: 2025 meta-GWAS). The explained variance was converted to the liability scale of the population assuming a lifetime disease risk of 1.5%. The analysis presented in Panel B excluded the All of Us dataset, as its individual-level data could not be merged with the other data sets due to data protection regulations. To visualize the increase in variance explained by the PGS, we also calculated PGS based on the first published BPD GWAS, consisting of the Germany sample (green triangles: 2017 GWAS) ^18^. Significance: 1^*^: *p*<0.05; 2^*^: *p*<0.01; 3^*^: *p*<0.005; 4^*^: *p*<0.001; 5^*^: *p*<1×10^−4^; 6^*^: *p*<1×10^−8^; 7^*^: *p*<1×10^−12^, 8^*^: *p*<1×10^−20, *^: *p*<1×10^−30^

### Genetic correlations

#### Correlations with disorders and traits of interest

In a targeted approach, we calculated genetic correlations with a selection of 50 GWAS of other disorders and traits relevant to BPD, including mental disorders, suicide, self-harm, trauma, substance use, physical health, pain, sleep, personality traits (Big Five GWAS including data from 23andMe, Inc.), and cognition. After Bonferroni correction for multiple testing (α=0.05/50=0.001), BPD showed significant genetic correlations with 43 of the 50 tested phenotypes. Among the psychiatric disorders, BPD showed the strongest correlations with post-traumatic stress disorder (PTSD) (*r*_*g*_=0.77), depression (*r*_*g*_=0.74), and attention deficit hyperactivity disorder (ADHD) (*r*_*g*_=0.67). In the other domains, substantial genetic correlations with |*r*_*g*_|>0.5 included material deprivation, chronic pain, broad antisocial behavior, loneliness, externalizing traits, measures of suicide, self-harm, and trauma. Lower but significant genetic correlations (*r*_*g*_<0.18) were observed for the somatic disorders type 2 diabetes and asthma, and body mass index (Fig. 4; Supplementary Table S22).

**Figure 4:**
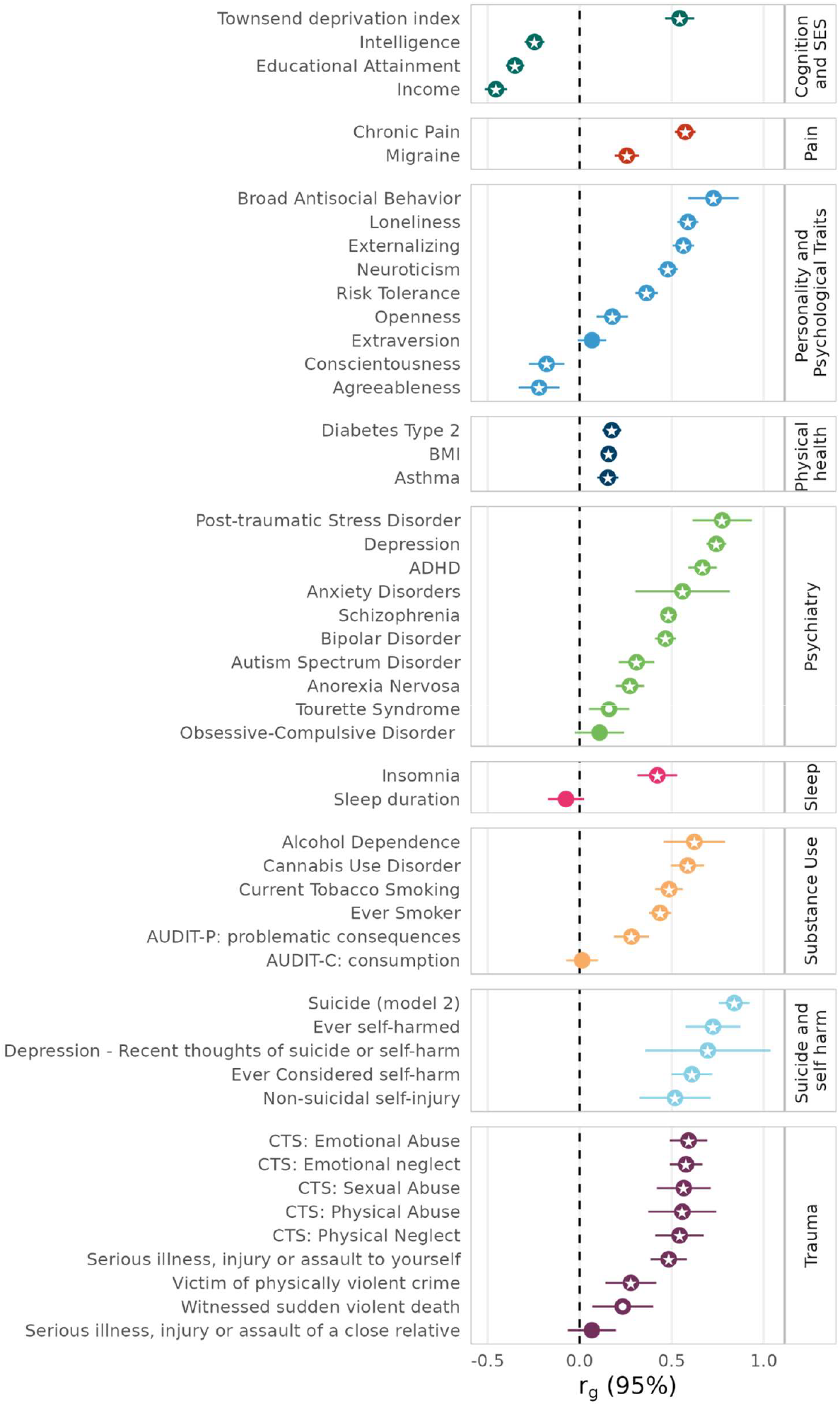
Genetic correlations of borderline personality disorder (BPD) with other phenotypes. Genetic correlations of BPD (total *N*_cases_=12,339, *N*_controls_=1,041,717) with 50 disorders and traits: within each group, disorders, and traits are sorted by their genetic correlation. White dot: *p*<0.05; White star: *p*<0.001 (0.05/50 tested correlations). 95% CI: 95% confidence interval; ADHD: attention deficit hyperactivity disorder; AUDIT: Alcohol Use Disorders Identification Test; BMI: body mass index; CTS: Childhood Trauma Screener; PTSD: post-traumatic stress disorder.

The sensitivity meta-analysis excluding subjects with SCZ/BIP showed comparable genetic correlations (Supplementary Fig. S49, Supplementary Table S23) with the 50 disorders and traits. Notably, lower but still substantial genetic correlations were observed with BIP (*r*_g_=0.35 vs. *r*_g_=0.47) and SCZ (*r*_g_=0.36 vs. *r*_g_=0.48). The genetic correlation of the main meta-analysis and the sensitivity analysis showed a genetic correlation of *r*_g_=1.00 (95%-CI [0,98-1,02]).

**Figure 5:**
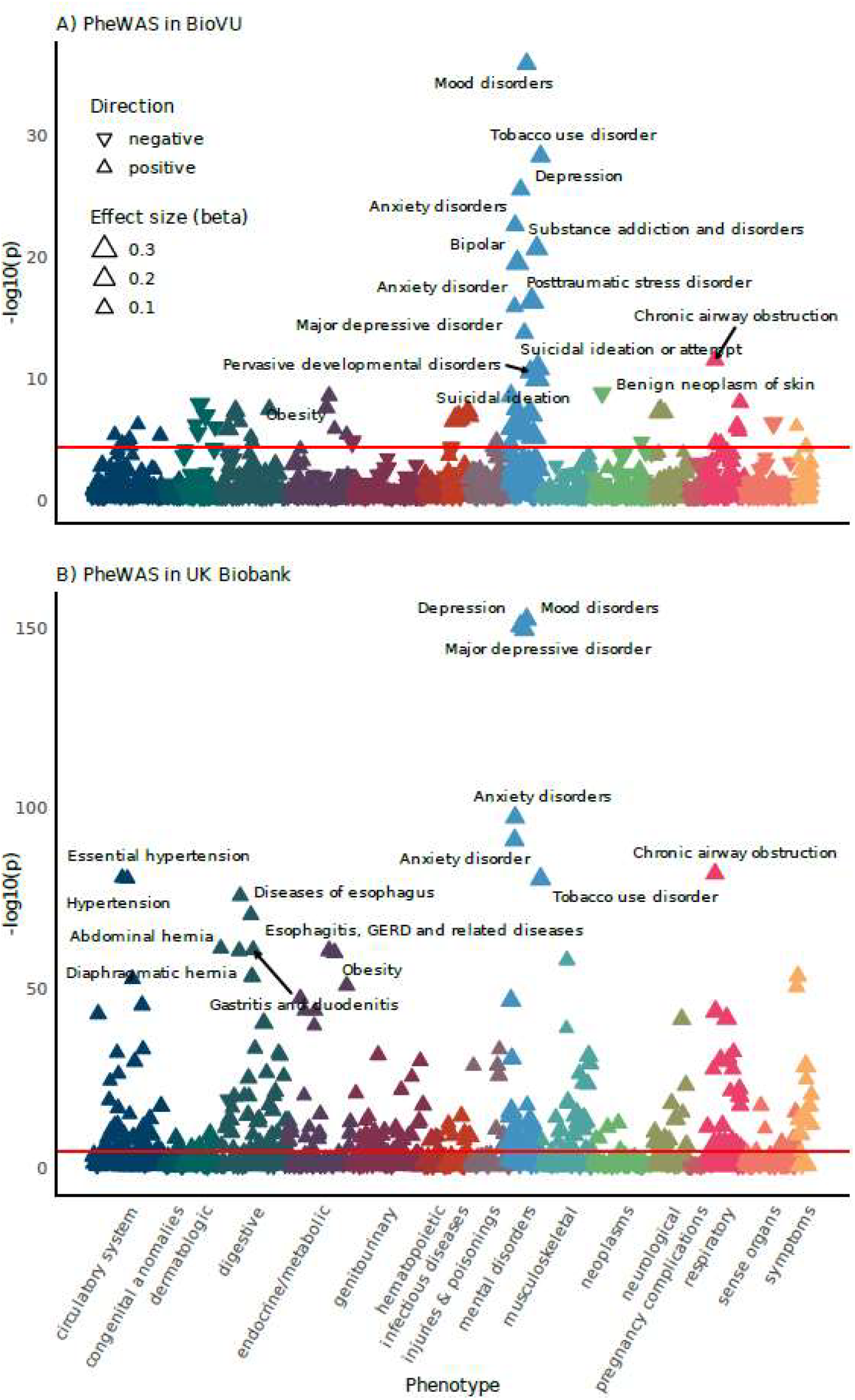
PheWAS of borderline personality disorder polygenic scores (BPD-PGS) in A) BioVU, and B) UKB: Association of BPD-PGS with 1,431 tested phecodes in BioVU and 1,250 Hospital Episode Statistics (HES) phecodes in UKB. Statistical significance (-log10(*p*)) is plotted on the y-axis and diagnoses are grouped by category. The red line indicates the significance threshold for Bonferroni correction for the tested phecodes (BioVU: 0.05/1,431=3.49×10^−5^; UKB: 0.05/1,250=4.00×10^−5^). The top 15 associations of each PheWAS are annotated. Upward-pointing triangles indicate positive associations, and downward-pointing triangles indicate negative associations. The size of the triangles indicates the effect size (beta).

### Phenome-wide association studies (PheWAS)

Complementing the targeted genetic correlation analysis, we characterized the genetic signal identified in the BPD GWAS, by performing two phenome-wide association studies (PheWAS). To this end, we tested the association of BPD-PGS (using PRS-CS^31^; excluding each target sample from discovery) with “phecodes”, *i*.*e*., medical phenotypes based on International Classification of Disease (ICD) diagnoses documented in the electronic health records (EHR), using data from the Vanderbilt University Medical Center Biobank (BioVU)^32^ (66,325 subjects;214 with the phecode 301.20: *“antisocial/borderline personality disorder”*) and Hospital Episode Statistics (HES) in the UK Biobank (UKB) (316,635 subjects; 211 with the phecode 301.20).

#### PheWAS in BioVU (EHR data)

In the PheWAS analysis in BioVU, 69 of the 1,431 tested diagnoses showed an association with BPD-PGS after Bonferroni correction (*p*_*adj*_<0.05; Supplementary Table S24). BPD-PGS showed the most statistically significant associations with codes from the mental disorders category (Extended Data Fig. 9), including mood disorders, substance use disorders, anxiety and PTSD, and suicidal ideation or attempt. Associations were also observed in other categories, including neurological (*e*.*g*., epilepsy) and somatic disorders (*e*.*g*., type 2 diabetes, chronic airway obstruction, hypertension), and pain disorders and symptoms.

#### PheWAS in UKB (HES data)

In the PheWAS analysis in the UKB, 317 of the 1,250 tested diagnoses showed an association with BPD-PGS (*p*_*adj*_<0.05; Supplementary Table S25). BPD-PGS showed the most statistically significant associations with codes from the mental disorders category (Extended Data Fig. 10), including mood disorders, tobacco use disorders, and anxiety disorders. As in BioVU, significant associations after Bonferroni correction were observed in other categories, including respiratory (*e*.*g*., chronic airway obstruction), digestive (*e*.*g*., esophagus, gastroesophageal reflux disease), circulatory, endocrine/metabolic, and neurological diagnoses, and pain disorders and symptoms.

Of the 69 diagnoses that were significant in the BioVU, 56 were also present with sufficient case numbers in the UKB. Of these, 51 showed effects in the same direction and 41 also reached significance in the UKB (*p*_*adj*_<0.05). In both samples, the phecode 301.20 (*“antisocial/borderline personality disorder”*) showed the strongest effect size (OR_BioVU_=1.41; OR_UKB_=1.54).

## Discussion

The present GWAS including 12,339 BPD cases and 1,041,717 controls marks a major advance in BPD genetics, with the demonstration of a substantial SNP-heritability, the identification of genome-wide significant risk loci, pathway and tissue enrichment, and a considerably improved predictive value of derived PGS. The study includes the first sex-stratified and X-chromosomal GWAS for BPD, and the first systematic investigation of shared genetic risk of the disorder. The validity and generalizability of the results is supported by the replication of the lead SNP associations and PGS analyses in independent datasets.

The substantial increase in sample size was achieved by using a strategy combining studies with different ascertainment strategies. The approach of our previous study, which included only clinical studies that had explicitly recruited BPD patients^18^, was extended by also including biobank and cohort studies that mainly linked diagnoses from electronic health records to genetic data. The high genetic correlation indicates the two subsets capture a largely identical genetic architecture.

A substantial proportion of the heritability observed in family and twin studies (46− 69%)^12,13^ was explained by common genetic variation (estimated SNP-heritability of 17%). This is consistent with GWAS in other psychiatric disorders, where SNP-heritability often accounts for approximately a third of the estimates from twin and family studies^16^. The substantial contribution of common genetic variation to BPD is also supported by the leave-one-out PGS analyses, predicting a weighted average of 4.6% of the variance on the liability scale. Importantly, the low LDSC intercept (1.02) and the attenuation ratio of 11% indicate the association is largely driven by the polygenic signal and not by confounding.

In the context of identifying genetic risk variants and genes, the present study is the first to identify six genome-wide loci associated with BPD. The genetic risk loci were located in or near genes including *SGCD, EXD3, FOXP1*, and *FOXP2*, and one locus mapped to several genes: *MVK, MMAB*, and *UBE3B. SGCD* encodes sarcoglycan delta, a component of the sarcoglycan complex, and mutations have been associated with muscular and cardiovascular disorders^33^. Common variation in *SGCD* has also been associated with mental disorders, like SCZ^34,35^ and PTSD^36^, substance use phenotypes^37–39^, and measures of quality of life^40^. Converging findings in rodents and humans further suggest that the sarcoglycan complex is expressed in the CNS in both neurons and astrocytes^41,42^ and may be involved in GABAergic neurotransmission ^41^ and cerebrovascular function^43^.

Among the genes mapped to genes by proximity to the lead SNPs or in the MAGMA gene-based analysis, the highest PoPS scores were observed for *SGCD, FOXP2, FOXP1, DEPDC1B. FOXP1* and *FOXP2* are both members of the Forkhead-box (FOX) transcription factor family that are expressed in the brain and are both associated with speech and language development^44^. The *FOXP2* gene has shown significant associations with externalizing behavior^45,46^ and related traits or disorders such as substance use disorders^47–50^, broad antisocial behavior^51^, and ADHD^52^ in several recent GWAS. Observed associations with childhood maltreatment^53^ and PTSD^36,54,55^ further suggest a trauma-related pathway for BPD, while recent GWAS of the Big Five personality traits showed an association of *FOXP2* with agreeableness^56,57^, conscientiousness^57^, and neuroticism^57^. Of the genes indicated by the combined analysis, *NME7* (NME/NM23 family member 7) and *KPNA2* (Karyopherin Subunit Alpha 2) had the highest PoPS scores: NME7 is a component of the gamma-tubulin ring complex which plays a role in microtubule organization^58^. It has been associated with antidepressant treatment response in a gene expression study^59^ and a GWAS^60^. KPNA2, a key component of the nucleocytoplasmic transport system, has mainly been studied with respect to cancer development, but was recently found with significant brain GWAS/QTL associations in a transcriptomic analysis of MDD^61^.

We present the first GWAS to include both sex-stratified and X-chromosome analyses — an initial step towards investigating the sex-specific genetic architecture of the disorder, which is relevant concerning the sex differences in BPD prevalence and presentation. We observed a substantially higher SNP-heritability in females, which has also been described for the two mental disorders showing the strongest genetic correlation with BPD–PTSD and depression^62^. The lead SNPs of the main GWAS, however, showed the same direction and similar effect sizes in the stratified analyses in both sexes, and the genetic correlation of the sex-stratified GWAS was high and did not differ from 1. However, it must be noted that the sex-stratified analyses had limited statistical power, particularly the male-only analysis, which was also indicated by a z-score of the SNP-heritability under 4^63^. Nevertheless, we consider the sex-stratified analysis an important first step and provide it as a resource. Sex-stratified analyses in larger samples may help to elucidate sex-specific risk factors of BPD^64^. Notably, for the first time, we comprehensively analyzed the genetic variation on the X chromosome regarding personality disorders and identified the gene encoding choline-phosphate cytidylyltransferase B (*PCYT1B*) as significantly associated with BPD in the gene-based analysis, as well as an association locus in the gene with genome-wide significance in the combined analysis. *PCYT1B* is expressed in the brain^65,66^, but has not been previously reported to be associated with mental health phenotypes. However, many previous GWAS analyses have not reported X chromosome data^64^.

This study is the first to systematically use genetic association data to investigate expression enrichment and potential drug repurposing for BPD, offering initial insights: The results suggest the importance of genes expressed in the brain and during early prenatal development. Single-cell analyses revealed significant SNP-heritability enrichment in genes expressed in the clusters *“medium spiny neurons”* as well as *“LAMP5-LHX6 and Chandelier cells”*. Both clusters have previously been demonstrated to be enriched in mental health phenotypes in a systematic analysis, namely in schizophrenia, IQ, educational attainment, and neuroticism^29^, with *“medium spiny neurons”* additionally showing enrichment in MDD^29^, and the *“Lamp5-LHX6 and Chandelier cell”* cluster being significantly enriched in the most recent bipolar disorder GWAS^67^. Together, these findings suggest that specific neuronal cell types with established relevance for psychiatric disorders may also play a role in BPD.

In addition, our analyses identified potential drug targets: Drug target analysis of the 16 prioritized genes highlighted *MYO1H*, which has been associated with the response to treatment with lithium, a mood stabilizer also suggested to reduce the risk of suicide attempts^68^. However, it should be noted that the gene was indicated by proximity only in the lithium GWAS^69^. The drug target analysis also suggested *PCYT1B* as a target of sphingosines. The sphingosine-1-phosphate receptor 3 (S1PR3) regulates various cellular processes when bound to its ligand Sphingosine-1-phosphate (S1P), has been implicated in stress resilience in animal studies, and was found to be downregulated in PTSD^70^. Notably, the S1P-S1P3 pathway was the only significant gene set in the present analysis. The genome-wide DRUGSETS analysis indicated drugs with high biological plausibility, including medication for neurological disorders and pain, albeit our results were not statistically significant after correction for multiple testing and should be interpreted with caution.

Through genetic correlation and PheWAS analyses, we address a critical gap in the literature by systematically and comprehensively assessing the shared genetic architecture of BPD across a broad spectrum of other disorders and traits. We confirmed the genetic correlations observed in the previous BPD GWAS^18^ with depression, BIP, SCZ^18^, neuroticism, openness to experience^71^, and loneliness^72^. With respect to the Big Five personality dimensions, the results highlight the enhanced power of the present BPD GWAS: We now additionally observe negative genetic correlations with agreeableness and conscientiousness, which is consistent with the profiles of the Big Five personality dimensions observed in cases with BPD^73–75^, as well as with data from twin models^76,77^. This suggests that genetic factors underlying variation in personality traits in the general population contribute to the risk for BPD. Among mental disorders, BPD showed the strongest genetic correlations with PTSD, depression, ADHD, and anxiety. Notably, these disorders are frequently observed as preceding or comorbid conditions and share clinical features with BPD^5,7,78^. The associations with suicide and self-harm are consistent with the clinical presentation of BPD^79^, the correlations with trauma phenotypes highlight the role these experiences play in BPD^10^, and the strong correlation of BPD with disorders from the internalizing/externalizing spectrum disorders suggests that BPD risk is influenced by the liability for both dimensions^79,80^.

In both PheWAS samples, and consistent with the genetic correlation results, the BPD-PGS were significantly associated with mood disorders, anxiety disorders, PTSD, substance use disorders, as well as suicidal ideation or suicide attempts. The strongest effect was observed for the phecode *“antisocial/borderline personality disorder”* supporting the specificity of the GWAS signal for BPD. BPD-PGS were also associated with a range of physical health phenotypes, including chronic airway obstruction, type 2 diabetes, obesity, and hypertension, for which an increased risk in BPD patients has been previously described^7^. The relatively small number of BPD cases in the PheWAS target samples makes it likely that these associations are driven by shared genetic risk, and not solely by comorbidities of BPD patients in the target samples. These results provide a promising starting point for the investigation of the shared genetic risks of BPD and somatic health, which is highly relevant as a large share of the reduced life expectancy of people with BPD is related to physical health problems^81^. Further research is warranted to understand the mechanisms through which inter-individual differences in genetic liability for BPD influence the risk for different disorders.

The present analyses cannot dissect to what degree comorbidities, the overlap of symptoms, and potential diagnostic misclassifications might have influenced our results^82^. While we addressed the potential overlap with BIP and SCZ, more fine-grained analyses are needed. In BPD, comorbidity with other mental disorders is the rule rather than the exception^1,5^, for example, approximately 70% of the investigated BPD patients had a history of depression. Restricting GWAS cases to those without psychiatric comorbidity would reduce the number of available subjects and limit the analysis to a less representative (and drastically smaller) subset. Therefore, we consider it a strength of the present analysis that the biobanks and cohorts providing summary statistics performed the main analysis without excluding cases with psychiatric comorbidity. To assess the impact of this strategy, the sensitivity analysis excluded subjects with either a diagnosis of BIP or SCZ, which are common exclusion criteria for dedicated BPD studies, accounting for 30% of the cases. At the level of the single-variant associations, we observed comparable effect sizes in the sensitivity analysis. We, therefore, consider it unlikely that SCZ or BIP comorbidity substantially biased the GWAS hits. Comparing the genetic correlation of the two iterations of the BPD GWAS with the GWAS of BIP and SCZ, we observe an attenuated but still substantial genetic correlation in the sensitivity analysis, suggesting shared genetic risk beyond the co-occurrence of the disorders.

The present study represents substantial progress in the identification of the genetic factors underlying BPD. However, the study has several limitations. First, the sample size is still relatively small compared to those of other mental disorders^16^, and the power analysis indicated that the statistical power was limited, especially for variants with a lower frequency. The inclusion of additional samples will likely lead to a substantial increase in the number of loci identified^83,84^. Furthermore, expanding the analyses to non-European ancestries is necessary to improve the understanding of the underlying genetic architecture and to facilitate the generalizability of the results^85,86^. Additionally, in this study, we investigated BPD as a categorical diagnosis, as it is also commonly used in the clinical context. However, the diagnosis can be heterogeneous and might differ between cohorts. Future studies should consider the heterogeneity of the disorder, and examine clinical BPD at a more fine-grained level, *e*.*g*., by examining individual symptoms or symptom clusters^87,88^. In addition, functional dimensions such as RDoC^89^ and classification systems such as HiTop^90,91^ as well as borderline personality traits or symptom dimensions^2,17,92^ should be incorporated.

In summary, the present GWAS meta-analysis represents a major step forward towards an understanding of the genetic etiology of BPD. It is the first GWAS of a personality disorder to identify genome-wide significant risk loci and genes, and demonstrate that BPD, like other mental disorders, is a complex polygenic disorder.

## Online methods

### Sample description

An overview of the analyses performed can be found in Fig. 1. We conducted a GWAS meta-analysis, including 1,054,056 participants of European ancestry (*N*_cases_=12,339 *N*_controls_=1,041,717, *N*_eff_=2×21,617). In total, 17 studies contributed individual-level data (*N*_cases_=2,705, *N*_controls_=4,600 controls). This included data from the prior BPD GWAS^18^, and data from an additional 1,712 cases and 3,061 controls (meeting DSM-IV criteria for BPD). Furthermore, we included data from 10 large-scale biobank or cohort studies that provided summary statistics from GWAS of BPD (*N*_cases_=9,634, *N*_controls_=1,037,117 controls). Of those, nine cohorts identified BPD status following ICD codes (ICD-10: F60.3, ICD-9: 3018D/301.83, and ICD-8: 3013), while in the GLAD study, a self-reported diagnosis was used.

Details on ascertainment, inclusion and exclusion criteria for cases and controls, and country of origin for each sample are documented in Supplementary Table S1. A detailed description of the study design, ascertainment of cases and controls, and genotyping array for each study, as well as quality control and imputation specific to studies sharing summary statistics are provided in Supplementary Table S2 and the Supplementary Methods. All subjects gave informed consent, and the studies were approved by the respective ethical committees.

### Genotyping, quality control, and imputation

Samples providing genotype data at the individual level were grouped into datasets based on array and ancestry, resulting in five datasets (Supplementary Table S2). Quality control and imputation were carried out using the RICOPILI pipeline ^93^. A detailed description can be found in the Supplementary Methods.

### Genome-wide association study (GWAS)

For the studies providing genotype data at the individual level, GWAS were carried out within each dataset using an additive logistic regression model using PLINK (v1.9)^94^ for imputed genetic dosage data with the relevant ancestry principal components (PCs; details in Supplementary Methods) included as covariates. Details on the association analyses for the cohorts providing summary statistics can be found in Supplementary Table S2 and Supplementary Methods. In addition to the main analysis, we performed sex-stratified analyses. Dedicated studies of BPD often exclude subjects with schizophrenia and bipolar disorder. To explore the influence of comorbidity with these severe mental disorders on our results, as a sensitivity analysis, association analyses were performed excluding subjects with a history of BIP or SCZ from the studies providing summary statistics (sensitivity analysis: *N*_cases_=8,618, *N*_controls_=1,027,690; Supplementary Tables S3–S5). In the case of the cohorts, where the control population was not filtered for BIP and SCZ in the main analysis (Copenhagen Hospital Biobank and Danish Blood Donor Study; deCODE; Mayo Clinic Biobank; FinnGen), subjects with BIP or SCZ were also excluded from the controls for the sensitivity analysis.

### Meta-analysis

Meta-analysis of all samples providing either individual-level data or summary statistics was conducted using the inverse variance weighted fixed effects model in METAL^95^ as implemented in RICOPILI^93^ with a genome-wide significance threshold of 5×10^−8^. To evaluate consistency of effect sizes, we computed Cochran’s Q-statistics and the corresponding heterogeneity p-values and I^2^ statistics, which quantifies the percentage of total variation due to heterogeneity. This procedure excluded single-nucleotide polymorphisms (SNPs) with minor allele frequency (MAF)<1% or an imputation quality score (INFO)<0.60. SNPs were translated to the HG19 genomic build (Human build GRCh37) when necessary, and both SNPs and SNP alleles were aligned to the haplotype reference consortium (HRC) reference genome to ensure standardization for the meta-analysis. To ensure a robust analysis with highly credible SNP sets, we excluded SNPs with highly significant heterogeneity (*p* <0.001) and SNPs with an effective sample size of less than 85%. X chromosome markers were included in the meta-analysis for all samples except the Norwegian Mother, Father and Child Cohort Study (MoBa), where data were not available at the time of analysis. To assess the similarity between the five datasets of studies providing individual-level data and the 10 studies providing summary statistics, separate meta-analyses were calculated for the two subsets.

The statistical power of the main analysis to detect genome-wide significant associations (*p*<5×10^−8^), was calculated using the Genetic Power Calculator^19^. For a range of allele frequencies (0.01–0.99), and an effect size of 1.1, we calculated the power of the main analysis (*N*_*eff*_ = 2×21,617, calculated as described in ^96^), and the required sample size for a power over 80% (details see Supplementary Table S6, Supplementary Figure S1).

SNP-heritability, based on the autosomal markers, was estimated using LDSC^20^, both as the observed SNP-heritability and as the SNP-heritability converted to the liability scale, using the *N*_*eff*_ (2×21,617=43,234)^96^, a corresponding sample prevalence of 0.5, and assuming a population prevalence of 1.5%^3^ for the main analysis. For the sex-stratified GWAS, we used sex-specific population prevalences of 2.25% in females and 0.75% in males for the conversion to the liability scale, corresponding to the previously described female:male ratio of 3:1^4,5^. To assess the contribution of confounding to the observed signal, we calculated the attenuation ratio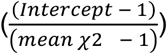.

To test whether the SNP-heritability estimates of the sex-stratified GWAS differed, and to test whether the genetic correlations between GWAS subsets (males vs. females; individual-level vs summary cohorts) differed from 1, we used the block jackknife extension of LDSC^20^, which has previously been described^97^.

To assess replication of the observed SNP associations, lead SNPs for the thresholds *p*<1×10^−6^ and *p*<5×10^−8^ were tested for replication in a meta-analysis of two independent datasets (Spain 2 & All Of Us; total N_cases_ = 685, total N_controls_ = 107,750). Additionally, for SNPs with *p*<1×10^−6^ in the discovery GWAS analysis, discovery and replication datasets were analyzed in a combined GWAS meta-analysis, and loci with *p*<5×10^−8^ in the combined analysis are reported.

### Gene-based and gene-set tests

Gene-based, gene-set, and tissue enrichment tests were carried out using MAGMA (version 1.07) ^21^ as implemented in FUMA (version 1.5.2) ^22^. Gene-based tests were performed using the summary statistics of the GWAS meta-analysis for a total of 19,843 genes, with SNPs assigned to genes based on their physical position. SNPs were included in the analysis using boundaries of 35 kilobases (kb) upstream and 10kb downstream of the genes. Gene-sets were tested by analyzing a total of 9,237 gene-sets (MSigDB v2023.1Hs; limited to gene sets with at least 20 mapped genes). For both gene-based and gene-set tests, a Bonferroni *p*-value threshold, corrected for the number of respective tests, was applied (gene-based α=0.05/19,843=2.5×10^−6^; gene-set α=0.05/9,237 =5.4×10^−6^).

Additionally, Polygenic Priority Scores (PoPS) ^23^ were calculated, which prioritize genes at GWAS loci using MAGMA gene-level association tests and over 57,000 gene features such as cell-type specific expression gene expression, biological pathways, and protein-protein interactions. PoPS scores were available for 17,702 autosomal genes, and PoPS scores and ranks are reported.

Tissue enrichment expression was carried out using Genotype-Tissue Expression (GTEx; version 8) data for 53 tissue types^24^, and data from BrainSpan, representing 29 different ages and 11 general developmental stages^25^ as implemented in FUMA (version 1.5.2)^22^. In addition, we utilized the Human Brain Atlas dataset containing single-nucleus RNA sequencing data from approximately 3.369 million nuclei derived from adult post-mortem brain samples, covering 106 anatomical sections^27^. We used the cell-types at the level of 31 “superclusters”, and maintained the existing nomenclature for cell-types. We calculated expression proportion values for each cell-type (“gene specificity”, described in detail in^29^ and^98^), resulting in ∼1300 genes per cell-type. We subsequently tested the enrichment of heritability in these gene sets using stratified LDSC ^28^, adjusting for the 53 baseline annotations. To adjust for multiple hypothesis testing in the tissue expression analyses, we applied a Bonferroni p-value threshold adjusted of the respective number of tested tissues.

### Drug target analysis

For the drug target analysis, we prioritized genes that either mapped to the genome-wide risk loci (Table 1) or were significant in the gene-based analysis. To identify drug targets among these genes, we extracted data from the Open Targets platform using the GraphSGL API. The information provided by Open Targets is based on the ChEMBL database ^99^. Additionally, we performed a tractability analysis (*i*.*e*., the potential to be modulated by a drug) to assess small molecule binding, the presence of accessible epitopes for antibody-based therapy, relevant data for using Proteolysis Targeting Chimeras (PROTACs), and the presence of compounds in clinical trials with modalities other than small molecules or antibodies. Databases were queried on 2024-06-24. Furthermore, we used the Genome for REPositioning drugs (GREP; https://github.com/saorisakaue/GREP) pipeline to test for drug target enrichment across clinical ATC or ICD categories ^100^. Finally, we queried the Drug Gene Interaction Database (DGIdb) ^101^.

In addition to these approaches focussing on the prioritized 16 genes, we explored potential drug targets with the DRUGSETS approach^30^, which integrates the genome-wide association signal. Using this approach we tested drug–gene sets, compiled from the Clue Repurposing Hub and the Drug–Gene Interaction Database, each comprising the genes whose protein products are known to be targeted by or to interact with a given drug. We then conducted a competitive gene-set analysis in MAGMA v1.08, conditioning on a background set of all 2,281 drug-target genes, to identify drug-gene sets significantly associated with BPD. To account for multiple testing across the 735 gene sets analyzed, we applied a Bonferroni threshold of *p*<0.05/735 = 6.80×10_-5_.

### Polygenic scores

We applied polygenic scoring (PGS) to predict case-control status in the datasets where individual-level genotype information was available. Polygenic scores were calculated using PRS-CS, a method that uses Bayesian regression to calculate updated (posterior) effect sizes by applying continuous shrinkage to the initial (prior) effect sizes from the discovery dataset using linkage disequilibrium (LD) information^31^. The posterior effect sizes account for the LD between SNPs using external LD reference panels constructed from the 1000 Genomes Project Phase 3 European samples. PGS were calculated based on both the present meta-analysis and also the prior BPD GWAS from 2017 (consisting of the Germany dataset) ^18^ for comparison. Using leave-one-out results of the present meta-analysis (2024 meta-GWAS), where the respective target dataset was always left out of the meta-analysis, we calculated PGS for all datasets with individual-level genotype information (Germany, Central Europe, Spain 1, Norway 1, and Norway 2 as well as the two replication cohorts (Spain 2 & All Of Us; total N_cases_=685, total N_controls_=107,750; Supplementary Tables S1&S2)). Similarly, we used the results of the prior BPD GWAS (2017 GWAS) to calculate PGS for the datasets not included in that analysis (Central Europe, Spain 1, Norway 1, Norway 2, and Spain 2, All Of Us).

As an effect measure of the association between BPD-PGS and case-control status, *Nagelkerke-pseudo-R2* (*NkR2*) was calculated comparing the *R2* of the full model including PGS and covariates (ancestry principal components) as predictors to the reduced (null) model including the covariates only. The resulting *NkR*^*2*^ was then converted to the liability scale ^102^ of the population assuming a lifetime disease risk of 1.5%^3^. Additionally, the area under the receiver operator characteristic curve (AUC) was calculated for each target dataset. Average *NkR2* and AUC were calculated across the five target datasets and weighted by the effective sample size of the respective target samples.

To calculate the odds ratio of the deciles of the PGS distribution, PRS-CS were normalized to have a mean of 0 and unit variance before combining them into two sets of scores for comparison: one based on Witt 2017 (*N*_controls_=3,496; *N*_cases_=1,758) and the other on BPD-meta 2024 (*N*_controls_=5,035; *N*_cases_=2,751). This part of the PGS analysis excluded the All of Us dataset, as its individual-level data could not be merged due to data protection regulations. Using the normalized PRS-CS, we categorized the data into ten deciles through quantile binning, assigning each observation a decile number ranging from 1 (lowest PRS-CS) to 10 (highest PRS-CS). We then created dummy variables by coding observations within each decile as cases, while those outside that decile were coded as controls. The dummy variable ranged from deciles 2 (Q2) to 10 (Q10), with decile 1 (Q1) serving as the reference category.

To assess the association between the deciles based on PRS-CS and the actual case and control status, we conducted logistic regression analyses using the decile-based dummy variables for Q2 to Q10. The odds ratios (OR) for each decile were calculated by exponentiating the coefficients from the logistic regression model, controlling for PCs (1, 2, 3, 4, 5, 6, 8, 10, 12, 14, 15, and 20). For the reference comparison (Q1/Q1), an odds ratio of 1 was assigned, indicating no effect. The odds ratios for each of the remaining deciles (Q2 to Q10) were computed accordingly OR_**Q1/Qj**_=exp(*βj*) for j=2,3,…,10. Additionally, 95%-CI for the odds ratios were calculated using the standard errors of the coefficients (CI_lower_=exp (*βj*–1.96×SEj); CI_upper_=exp *(βj*+1.96×SEj)). As an additional measure of effect, the highest decile (Q10) was compared to the middle 10% of the distribution as reference, and OR and CI were calculated as described above.

### Genetic correlations

Genetic correlations between subsets and with other disorders and traits were calculated using LD score regression^20^. Calculations were carried out with a free intercept and the 1000 Genomes dataset (EUR) as a reference LD structure panel ^103^.

In a targeted approach, genetic correlations were calculated with 50 GWAS of a range of other disorders and traits relevant to BPD, selected based on reported phenotypic and genetic associations in the literature, theoretical consideration, and data availability. Those included GWAS of mental disorders, suicide, self-harm, trauma, substance use, physical health, pain, sleep, personality traits (Big Five GWAS including data from 23andMe, Inc.), and cognition (Supplementary Table S22). A Bonferroni p-value threshold, corrected for the number of respective tests was applied (α=0.05/50=1×10^−3^).

### Phenome-wide association studies (PheWAS)

The targeted genetic correlation analyses were complemented by PheWAS, systematically exploring associations of the polygenic predisposition for BPD with over 1,000 diagnosis-based phenotypes across the diagnostic spectrum.

#### PheWAS in BioVU (EHR data)

Data from the Vanderbilt University Medical Center Biobank (BioVU) ^32^ were used to test the association of BPD-PGS with medical phenotypes based on two or more International Classification of Disease (ICD) diagnoses (“phecodes”; Phecode Map 1.2) documented in the electronic health records (EHR). For each phecode, a case was defined as having ≥1 ICD code in the phecode group on ≥2 distinct dates, and control was defined as having 0 codes in that phecode group. Individuals with only 1 single instance of a code in that phecode group were excluded from the case/control definition for that phecode. PGS were calculated in 66,325 unrelated subjects of European genetic ancestry (214 individuals with phecode 301.20 (*“antisocial/borderline personality disorder”*) based on summary statistics excluding the BioVU samples from the discovery meta-analysis (12,024 cases; 1,038,567 controls). PGS were computed using a continuous shrinkage prior (CS) to SNP effect sizes using PRS-CS^31^ and standardized to have a mean of 0 and a standard deviation of 1. Phecodes were tested for association with BPD-PGS when at least 100 cases were available using logistic regression models with sex (defined as sex reported at birth from the EHR), age, and the first 10 genetic PCs as covariates. 1,431 phecodes were included, and a Bonferroni-corrected threshold of α=0.05/1,431=3.49×10^−5^ tests was applied.

#### PheWAS in UKB

An additional PheWAS analysis was carried out using data from the UK Biobank (UKB). Here, PGS for BPD were calculated using PRS-CS based on summary statistics excluding the UKB samples from the discovery meta-analysis (12,157 cases; 1,039,897 controls) generated by excluding the UKB. The PheWAS was calculated using phecodes (Phecode Map 1.2) based on diagnoses recorded in Hospital Episode Statistics (HES) in a maximum sample of 316,635 subjects (211 individuals with phecode 301.20). For each phecode, a case was defined as having ≥1 ICD code in the phecode group, and control was defined as having 0 codes in that phecode group. A total of 1,250 phecodes with at least 100 cases were tested for association using a Bonferroni-corrected threshold of α=0.05/1,250=4.00×10^−5^ tests.

## Data availability statement

Individual-level data are not publicly available due to ethical restrictions. The results of the meta-analysis data will be publicly available for download upon publication of the manuscript via (via figshare.com; https://pgc.unc.edu/for-researchers/download-results/).

GWAS summary statistics of other traits and disorders used for analyses in this study are publicly available (sources listed in Supplementary Table S22). There exception are the GWAS summary statistics for self-harm^104^ which were provided by the respective corresponding authors and the GWAS used for the Big Five personality traits (except Neuroticism) which include data from 23andMe, and can be made available to qualified investigators, if they enter into an agreement with 23andMe that protects participant confidentiality. This study used data from the All of Us Research Program’s Controlled Tier Dataset v7, available to authorized users on the Researcher Workbench.

## Code availability statement

Quality control and imputation were carried out using RICOPILI (https://sites.google.com/a/broadinstitute.org/ricopili/; https://github.com/Ripkelab/ricopili). GWAS in datasets with individual data were carried out within each dataset using PLINK 1.9 (http://pngu.mgh.harvard.edu/purcell/plink/). Prephasing/imputation was carried out using EAGLE 2.4.1 (https://alkesgroup.broadinstitute.org/Eagle/) and MINIMAC3 (http://genome.sph.umich.edu/wiki/minimac3). Meta-analyses of GWAS were performed using METAL: (http://www.sph.umich.edu/csg/abecasis/metal/). Genetic correlations and heritability estimates were calculated using LDSC (https://github.com/bulik/ldsc).

FUMA 1.5.2 was used for downstream analyses including gene-based and gene-set analyses (https://fuma.ctglab.nl/), based on MAGMA 1.07 (http://ctglab.nl/software/magma).

Additionally, genes were prioritized using Polygenic Priority Score (PoPS) v0.2: (https://github.com/FinucaneLab/pops).

Enrichment for drug targets was carried out using GREP (https://github.com/saorisakaue/GREP), and DRUGSETS (https://github.com/nybell/drugsets). Weights for polygenic scores were generated using PRS-CS v.1.1.0 (https://github.com/getian107/PRScs).

## Acknowledgments

We thank all research participants and all researchers and clinicians who collected, generated, or processed the data used in this study. We thank all research participants and employees of 23andMe for making this work possible. We gratefully acknowledge *All of Us* participants for their contributions, without whom this research would not have been possible. We also thank the National Institutes of Health’s *All of Us* Research Program for making available the participant data examined in this study. We thank FinnGen for providing access to the meta-analysis results. The Genotype-Tissue Expression (GTEx) Project was supported by the Common Fund of the Office of the Director of the National Institutes of Health, and by NCI, NHGRI, NHLBI, NIDA, NIMH, and NINDS.

Fabian Streit is supported by a 2023 NARSAD Young Investigator Grant (#31537) from the Brain & Behavior Research Foundation with support from the Families for Borderline Personality Disorder Research. This research was supported by the Hector foundation II. Björn H Schott receives funding from the German Research Foundation (DFG CRC 1436, SP A05). Alice Braun was supported by the European Union’s Horizon program grant 101057454 - “PsychSTRATA”, by the German Research Foundation grant 402170461 - “TRR265”, and the Berlin Institute of Health at Charité. Elizabeth C Corfield was supported by the RCN (#274611) and the South-Eastern Norway Regional Health Authority (Helse Sør-Øst; #2021045). Ted Reichborn-Kjennerud was supported by the RCN (#274611). AH is supported by the Research Council of Norway (#336085), the South-Eastern Norway Regional Health Authority (#2020022, #2922083, #2022039, #2019097), and the European Union’s Horizon Europe Research and Innovation programme (FAMILY #101057529; HOMME #101142786; Marie Skłodowska-Curie grant ESSGN #101073237). Urs Heilbronner was supported by the European Union’s Horizon 2020 Research and Innovation Program (PSY-PGx, grant agreement No 945151) and the DFG (project number 514201724). Søren Brunak received funding from the Novo Nordisk Foundation (grants NNF17OC0027594 and NNF14CC0001). Ole Kristian Drange was supported by funding from the Research Council of Norway (RCN #223273, #324499). Kelli Lehto was supported by the Estonian Research Council (grant PSG615), Estonian Centre of Excellence for Well-Being Sciences, funded by the Estonian Ministry of Education and Research (grant TK218) and the European Union’s Horizon program grant 847776. Jean Gonzalez received funding from the National Institutes of Health (grant T32GM139790).

VN was supported by the DFG (DFG, NI 1332/16-1), and the German Federal Ministry of Education and Research (BMBF) through support of the German Center for Mental Health (DZPG, 01EE2306B)] This research was supported by the NHMRC grants 1172917 and 2025674.

## Conflicts of Interest

OAA is a consultant to Cortechs.ai and Precision Health and has received speaker’s honoraria from Lundbeck, Janssen, Sunovion, and Otsuka.

SB has ownerships in Hoba Therapeutics Aps, Novo Nordisk A/S, Lundbeck A/S, and Eli Lilly and Co.

JLK is a Scientific Advisory Board member of Myriad Neuroscience.

RM has received financial research support from Böhringer-Ingelheim and Otsuka Pharmaceuticals. He has received speakers’ honoraria from Otsuka Pharmaceuticals and Lundbeck and is a member of the advisory board of Böhringer-Ingelheim.

FP is a member of Scientific Advisory Boards of Brainsway Inc., Jerusalem, Israel, and of Sooma, Helsinki, Finland. He has received speaker’s honoraria from Mag&More GmbH, and the neuroCare Group, Munich, Germany.

JARQ was on the speakers’ bureau and/or acted as a consultant for Biogen, Idorsia, Casen-Recordati, Janssen-Cilag, Novartis, Takeda, Bial, Sincrolab, Neuraxpharm, Novartis, BMS, Medice, Rubió, Uriach, Technofarma and Raffo in the last 3 years. He also received travel awards (air tickets + hotel) for taking part in psychiatric meetings from Idorsia, Janssen-Cilag, Rubió, Takeda, Bial, and Medice. The Department of Psychiatry chaired by him received unrestricted educational and research support from the following companies in the last 3 years: Exeltis, Idorsia, Janssen-Cilag, Neuraxpharm, Oryzon, Roche, Probitas, and Rubió.

CS received lecturer fees from Idorsia Pharmaceuticals GmbH (2024).

MMN has received fees for membership in an advisory board from HMG Systems Engineering GmbH (Fürth, Germany), for membership in the Medical-Scientific Editorial Office of the Deutsches Ärzteblatt, for review activities from the European Research Council (ERC), and for serving as a consultant for EVERIS Belgique SPRL in a project of the European Commission (REFORM/SC2020/029). MMN receives salary payments from Life & Brain GmbH and holds shares in Life & Brain GmbH.

The other authors declare no competing interests.

## Extended Data Figures

**Extended Data Fig. 1:**
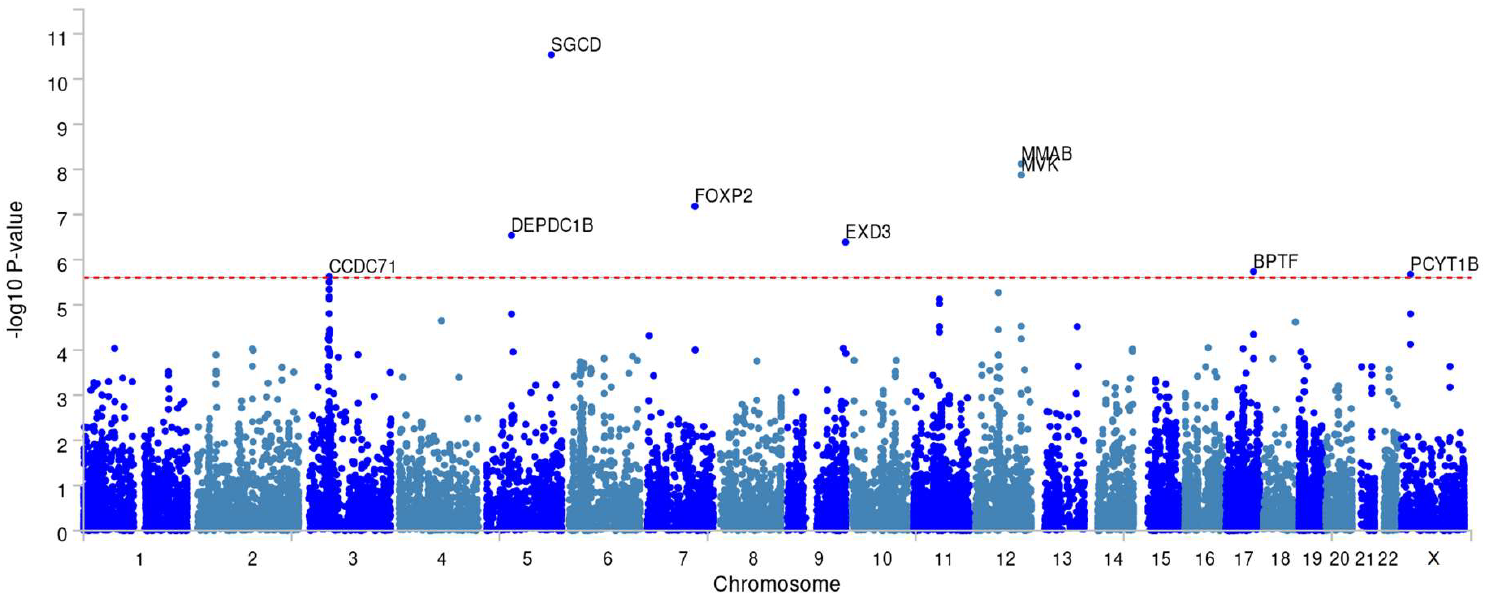
Manhattan plot of the gene-based analysis GWAS meta-analysis of BPD (N_cases_ = 12,339, N_controls_ = 1,041,717). The –log10 *p-value* for each gene is indicated on the y-axis (chromosomal position shown on the x-axis). The red line indicates significance after correction for multiple testing (*p*<0.05/19,843 = 2.5×10^−6^). Gene names are given for significant genes.

**Extended Data Fig. 2:**
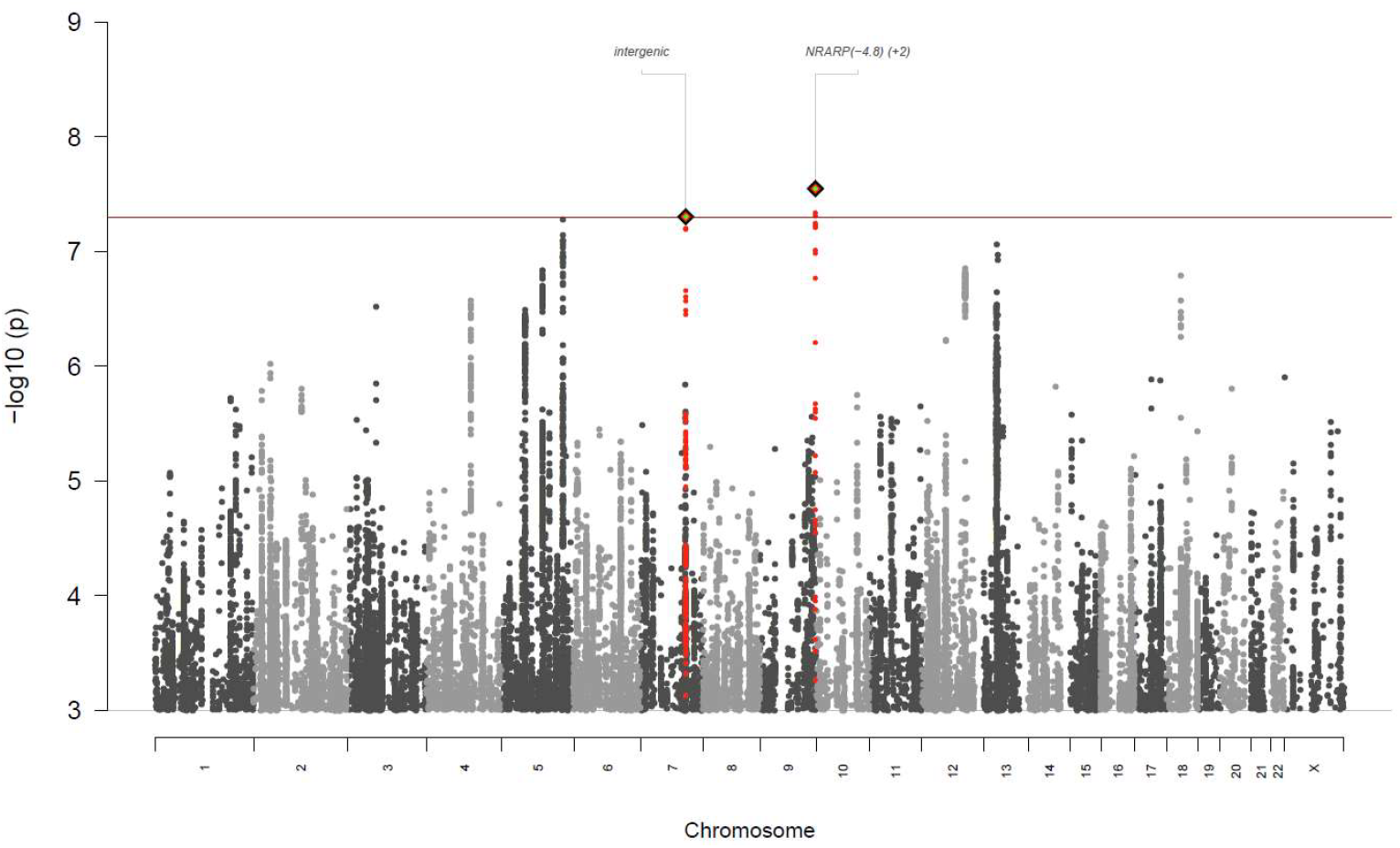
Female subset - Manhattan plot of the GWAS meta-analysis of BPD (N_cases_ = 10,025, N_controls_ = 547,333). The –log10 *p-value* for each single-nucleotide polymorphism (SNP) is indicated on the y-axis (chromosomal position shown on the x-axis). The red line indicates genome-wide significance (*p*<5×10^−8^). Index SNPs representing independent genome-wide significant associations are highlighted in diamonds, and SNPs in linkage disequilibrium with the index SNPs are highlighted in green.

**Extended Data Fig. 3:**
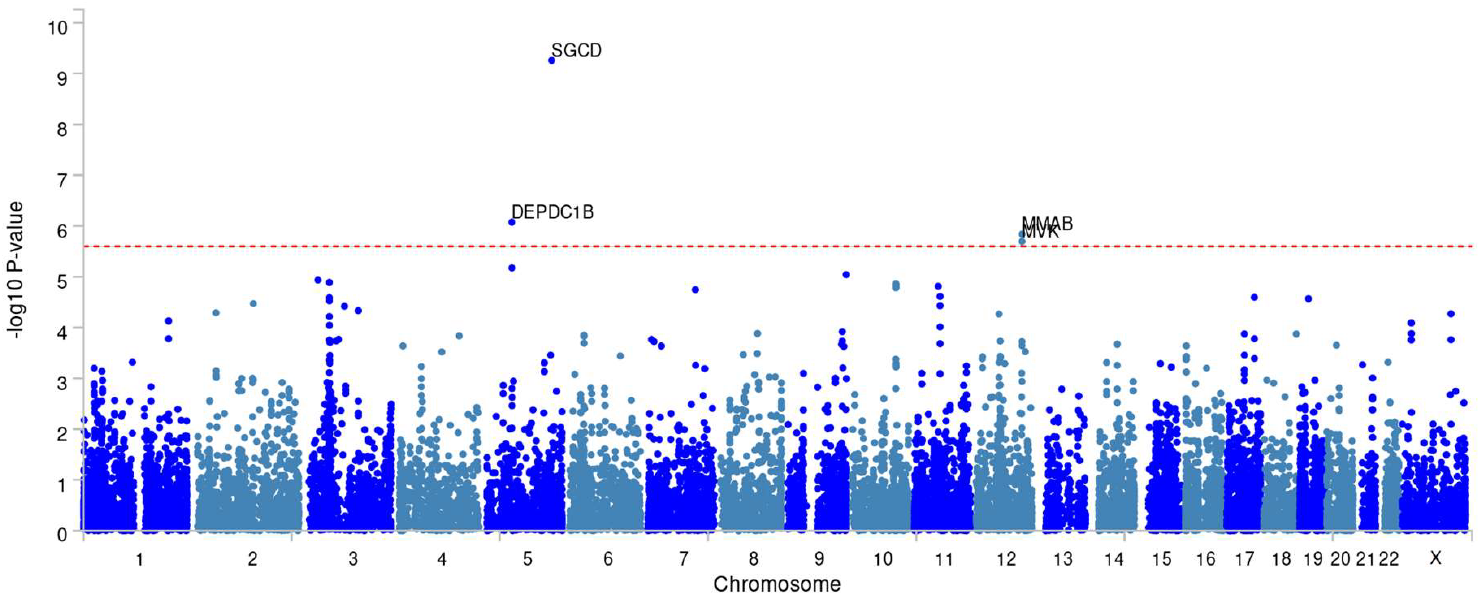
Female subset - Manhattan plot of the gene-based analysis female GWAS meta-analysis of BPD (N_cases_ = 10,025, N_controls_ = 547,333). The –log10 *p-value* for each gene is indicated on the y-axis (chromosomal position shown on the x-axis). The red line indicates significance after correction for multiple testing (*p*<0.05/19,844=2.5×10^−6^). Gene names are given for significant genes.

**Extended Data Fig. 4:**
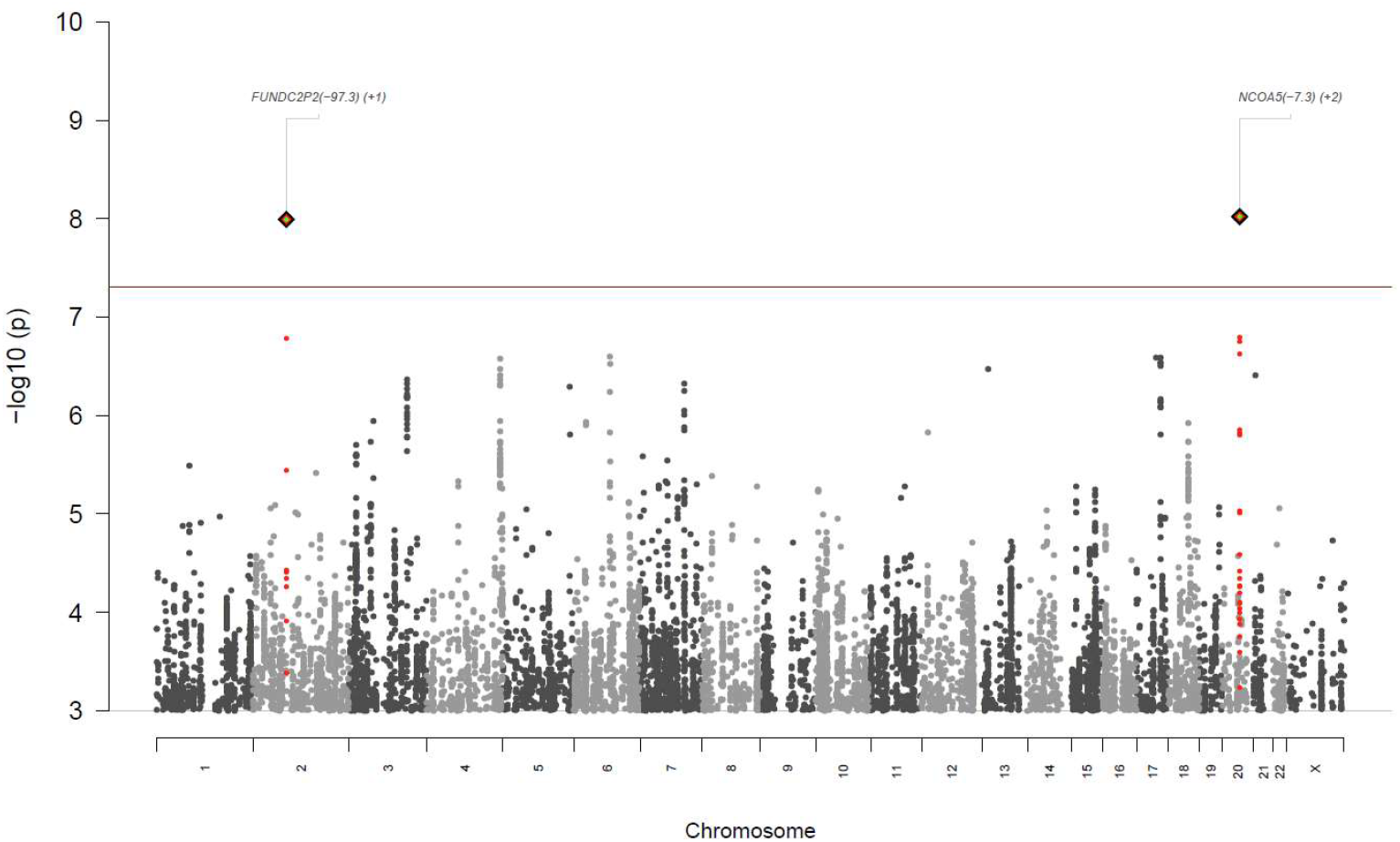
Male subset - Manhattan plot of the GWAS meta-analysis of BPD (N_cases_ = 2260, N_controls_ = 485,444). The –log10 *P-value* for each single-nucleotide polymorphism (SNP) is indicated on the y-axis (chromosomal position shown on the x-axis). The red line indicates genome-wide significance (*p*<5×10^−8^). Index SNPs representing independent genome-wide significant associations are highlighted in diamonds, and SNPs in linkage disequilibrium with the index SNPs are highlighted in red.

**Extended Data Fig. 5:**
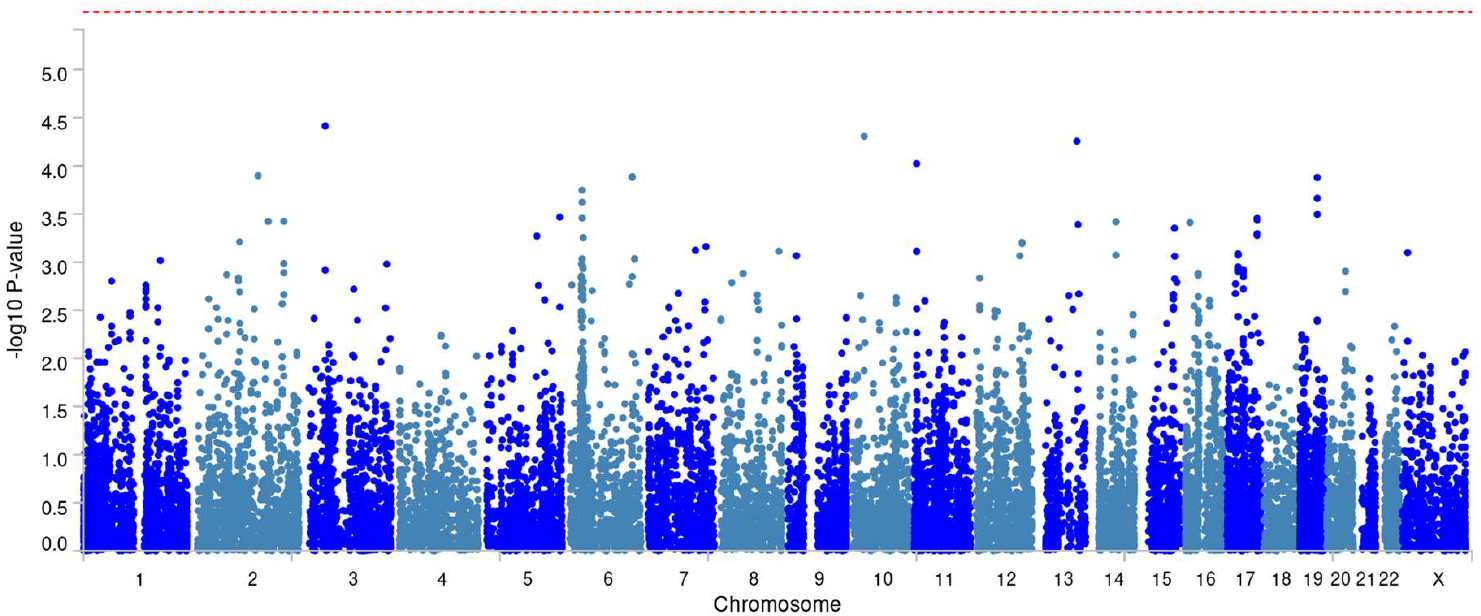
Male subset - Manhattan plot of the gene-based analysis (N_cases_ = 2260, N_controls_ = 485,444). The –log10 *p-value* for each gene is indicated on the y-axis (chromosomal position shown on the x-axis). The red line indicates significance after correction for multiple testing (*p*<0.05/19,856=2.5×10^−6^). Gene names are given for significant genes.

**Extended Data Fig. 6:**
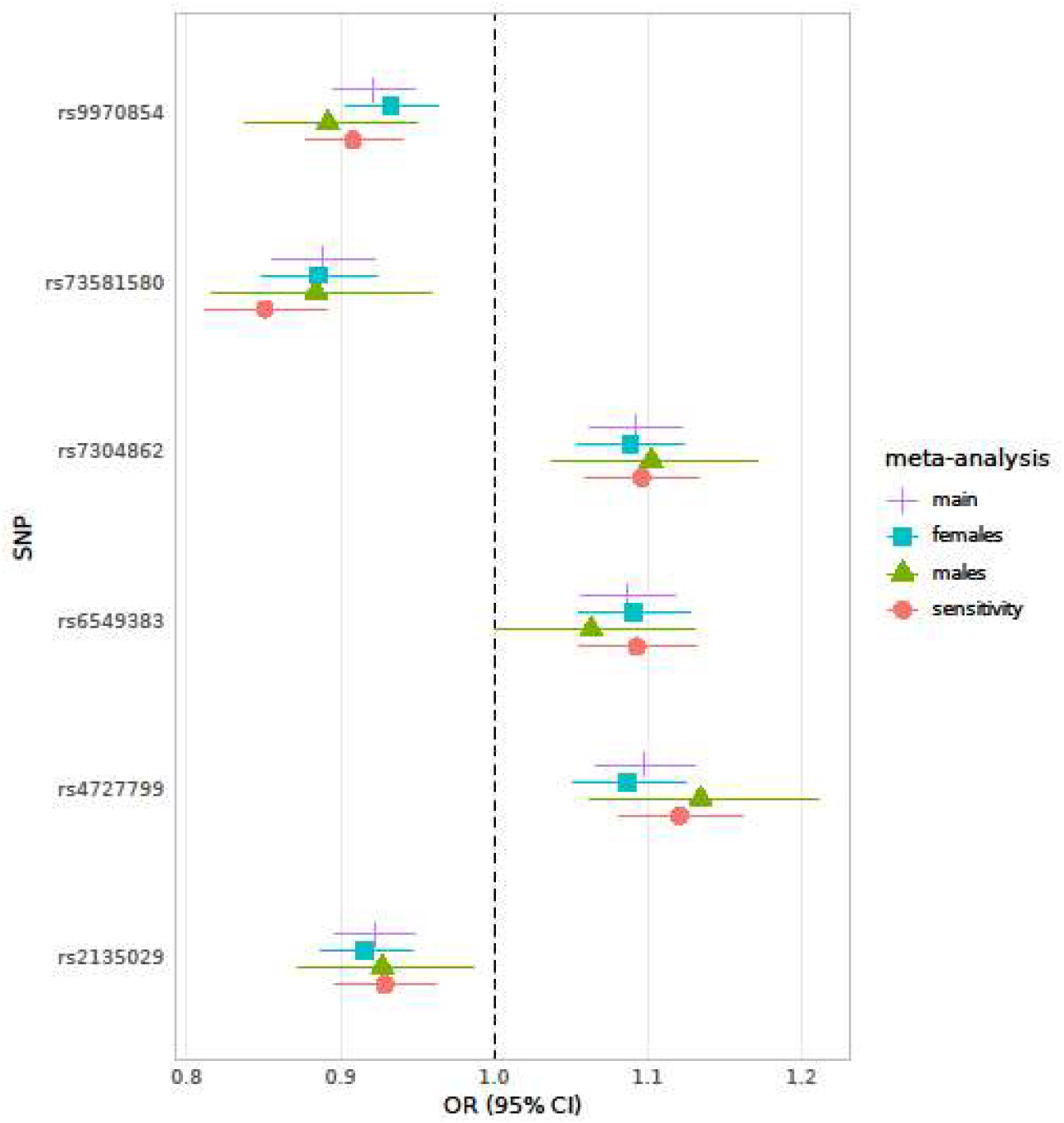
Comparison of the effect sizes of the six GWAS lead SNPs. Odds ratios (OR) are plotted for the main meta-analysis, the sex-stratified analyses, and the sensitivity analysis excluding cases with comorbidity of schizophrenia or bipolar disorder from the cohorts providing summary statistics. OR = odds ratio; 95% CI = 95% confidence interval; SNP = single nucleotide polymorphism

**Extended Data Fig. 7:**
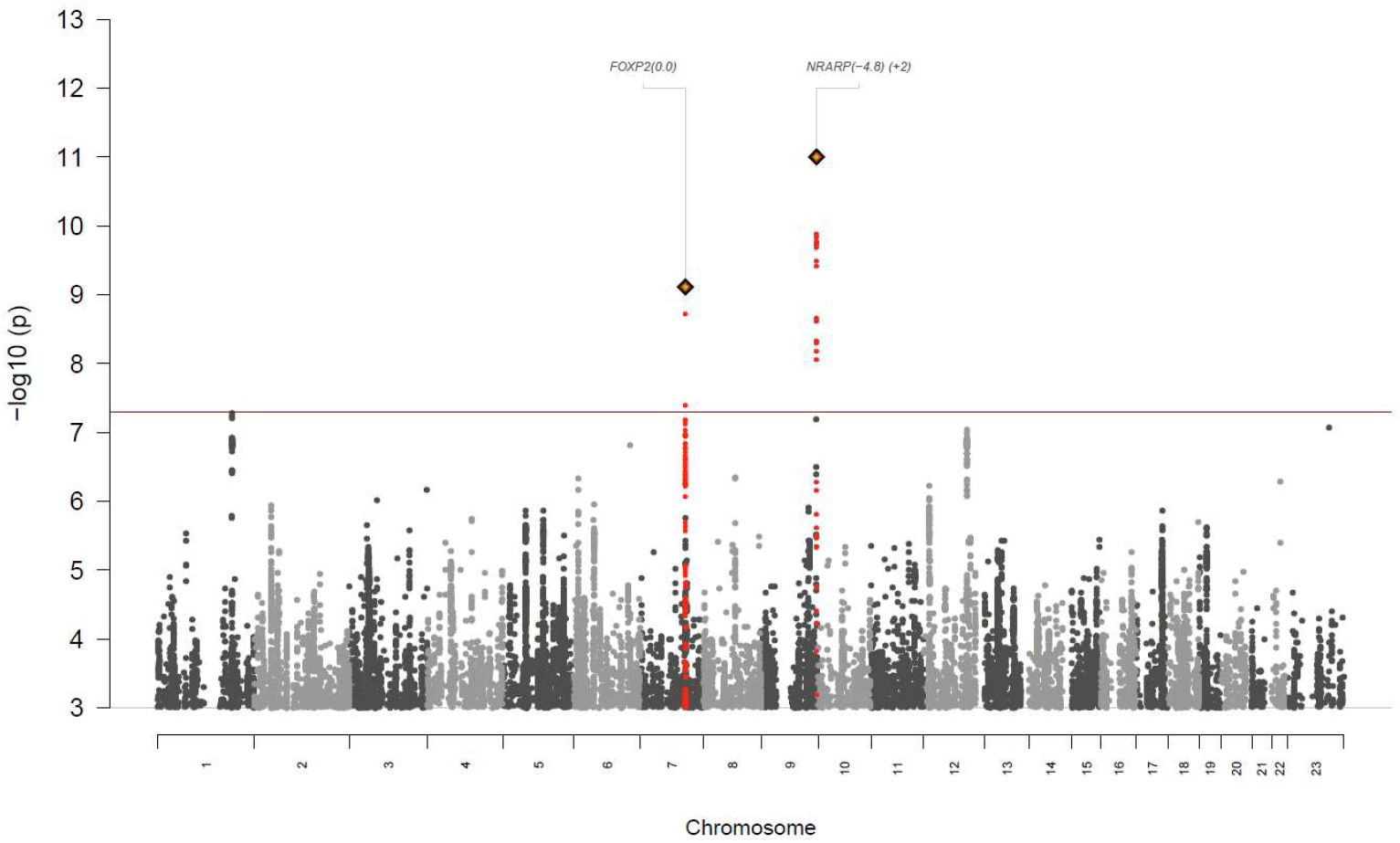
Sensitivity analysis - Manhattan plot of the GWAS meta-analysis of BPD excluding subjects with schizophrenia or bipolar disorder (N_cases_ = 8,618, N_controls_ = 1,027,690). The –log10 *p-value* for each single-nucleotide polymorphism (SNP) is indicated on the y-axis (chromosomal position shown on the x-axis). The red line indicates genome-wide significance (*p*<5×10^−8^). Index SNPs representing independent genome-wide significant associations are highlighted in diamonds, and SNPs in linkage disequilibrium with the index SNPs are highlighted in red.

**Extended Data Fig. 8:**
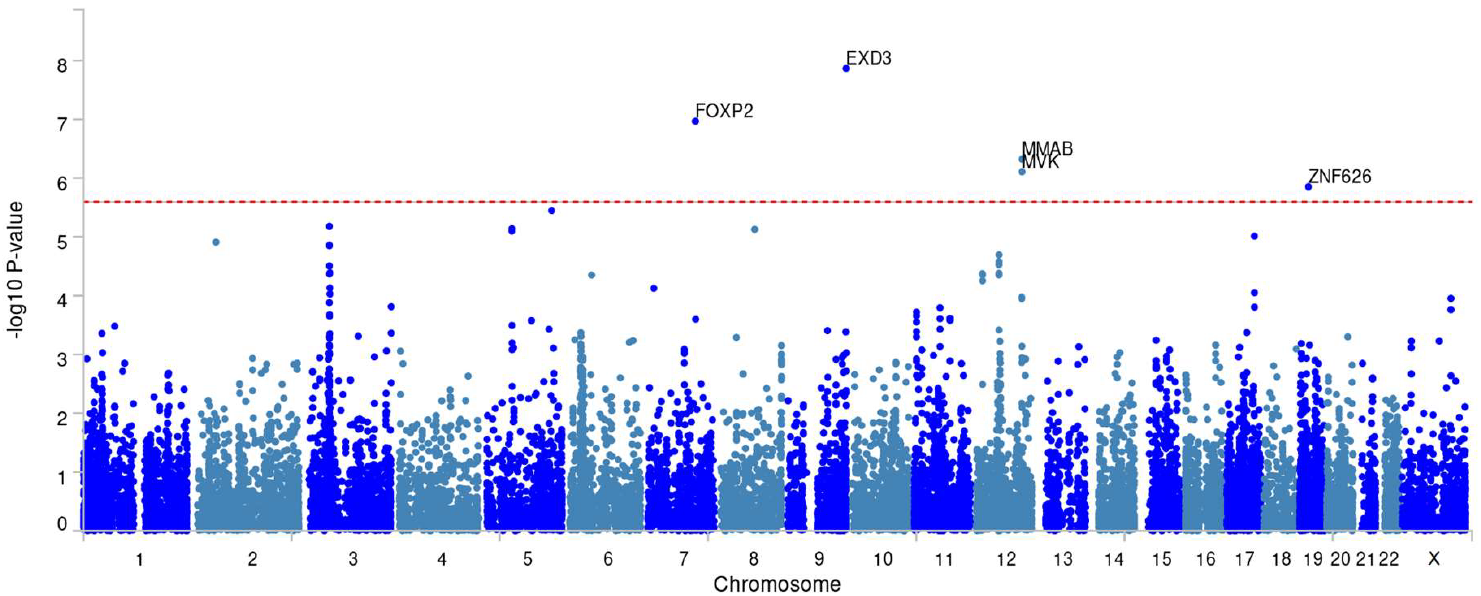
Sensitivity analysis - Manhattan plot of the gene-based analysis in the subset excluding subjects with schizophrenia or bipolar disorder - (N_cases_ = 8,618, N_controls_ = 1,027,690). The –log10 *p-value* for each gene is indicated on the y-axis (chromosomal position shown on the x-axis). The red line indicates significance after correction for multiple testing (*p*<0.05/19,842=2.5×10^−6^). Gene names are given for significant genes.

**Extended Data Fig. 9:**
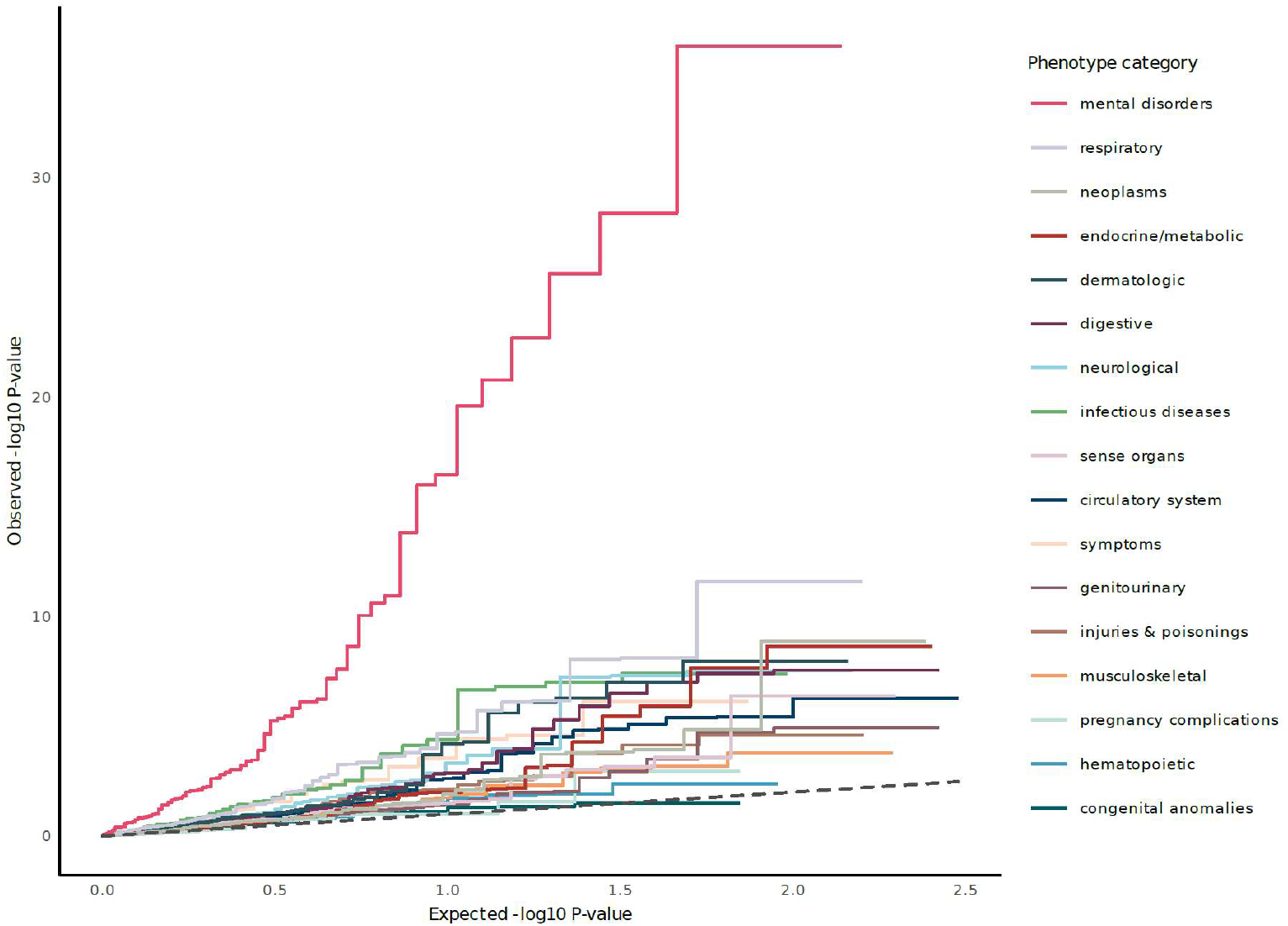
Quantile–Quantile plot of the PheWAS analysis in BioVU, stratified by phecode category (1,431 tested phecodes). Observed –log10 *p-values* are shown on the y-axis, and expected –log10 *p-values* are shown on the x-axis.

**Extended Data Fig. 10:**
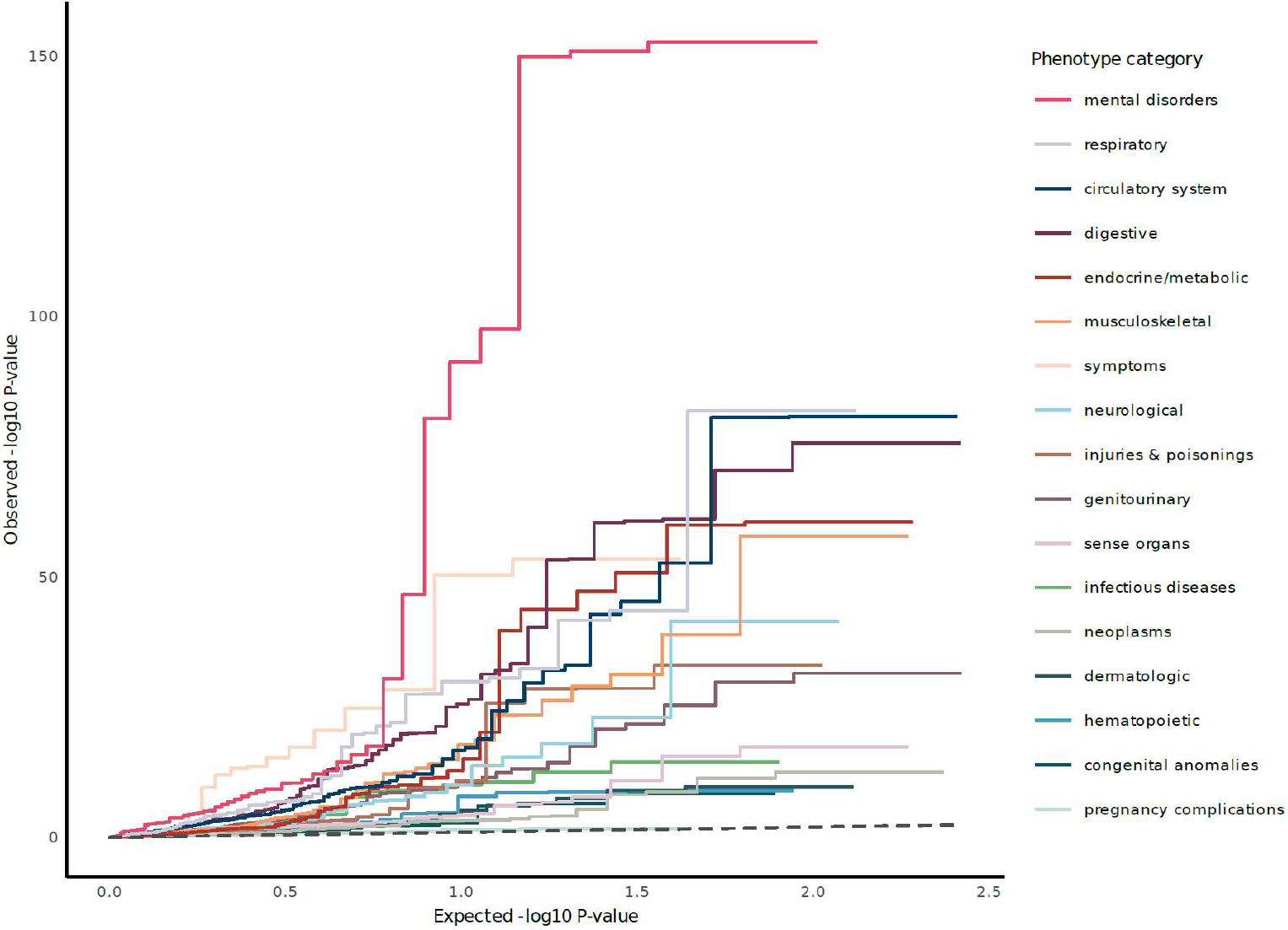
Quantile–Quantile plot of the PheWAS analysis in UKB, stratified by phecode category (1,250 tested phecodes). Observed –log10 *p-values* are shown on the y-axis, and expected –log10 *p-values* are shown on the x-axis.

