## Supplementary Methods for "Genome-wide association study of borderline personality disorder identifies 11 loci and highlights shared risk with mental and somatic disorders"

#### Table of Contents

**Supplementary text** 3
**Borderline personality disorder symptoms** 3
**Genotyping, quality control, and imputation** 3
**Description of studies providing individual-level data** 4
Witt2017 4
KFO256 6
BerlinCharité 7
Tuebingen 8
Basel 10
Geneva 11
MultiRCT 12
Hope 14
FASTER 15
SleepCIMH 17
Barcelona 18
SpainControls 19
POSEIDON 20
Munich 21
Oslo 22
Montreal 23
**Description of studies providing summary statistics** 25
BioVU 25
Copenhagen Hospital Biobank (CHB) and the Danish Blood Donor Study (DBDS) 26
deCODE 28
Estonian Biobank 30
HUNT 32
MoBa (Norwegian Mother, Father and Child Cohort Study) 35
UK Biobank 37
Mayo Clinic Biobank 38
GLAD 39
FinnGen 43
**Description of replication studies** 45
IPM-Reus (Spain 2 replication) 45
All of Us 46
**References** 47
**Members of contributing groups** 51

HUNT All-In Psychiatry 51
Estonian Biobank Research Team 52
The GLAD Study Group Authors 52
DBDS Genomic Consortium 54
**Supplementary Figures 57**
Figure S1: Power analysis of the discovery GWAS analysis 57
Figure S2: Quantile–Quantile plot of the GWAS meta-analysis of BPD (Ncases = 12,339, Ncontrols = 1,041,717) 58
Figure S3: region plot of rs9970854 59
Figure S4: forest plot of rs9970854 60
Figure S5: region plot of rs6549383 61
Figure S6: forest plot of rs6549383 62
Figure S7: region plot of rs2135029 63
Figure S8: forest plot of rs2135029 64
Figure S9: region plot of rs4727799 65
Figure S10: forest plot of rs4727799 66
Figure S11: region plot of rs73581580 67
Figure S12: forest plot of rs73581580 68
Figure S13: region plot of rs7304862 69
Figure S14: forest plot of rs7304862 70
Figure S15: region plot of rs2143288 71
Figure S16: forest plot of rs2143288 72
Figure S17: region plot of rs12466671 73
Figure S18: forest plot of rs12466671 74
Figure S19: region plot of rs10953781 75
Figure S20: forest plot of rs10953781 76
Figure S21: region plot of rs7210027 77
Figure S22: forest plot of rs7210027 78
Figure S23: region plot of rs5944622 79
Figure S24: forest plot of rs5944622 80
Figure S25: Quantile–Quantile plot of the gene-based analysis (Ncases = 12,339, Ncontrols = 1,041,717) 81
Figure S26: Female subset - Quantile–Quantile plot of the GWAS meta-analysis of BPD (Ncases = 10,025, Ncontrols = 547,333) 82
Figure S27: Female subset - region plot of rs73581580 83
Figure S28: Female subset - forest plot of rs73581580 84
Figure S29: Female subset - region plot of rs10227454 85
Figure S30: Female subset - forest plot of rs10227454 86
Figure S31: Female subset - Quantile–Quantile plot of the gene-based analysis (Ncases = 10,025, Ncontrols = 547,333) 87
Figure S32: Male subset - Quantile–Quantile plot of the GWAS meta-analysis of BPD (Ncases = 2260, Ncontrols = 485,444) 88
Figure S33: Male subset - region plot of rs6032676 89
Figure S34: Male subset - forest plot of rs6032676 90
Figure S35: Male subset - region plot of rs17757829 91

Figure S36: Male subset - forest plot of rs17757829 92
Figure S37: Male subset - Quantile–Quantile plot of the gene-based analysis (Ncases = 2260, Ncontrols = 485,444) 93
Figure S38: Sensitivity analysis - Quantile–Quantile plot of the GWAS meta-analysis of BPD excluding subjects with schizophrenia or bipolar disorder (Ncases = 8,618, Ncontrols = 1,027,690) 94
Figure S39: Sensitivity analysis - region plot of rs4727799 94
Figure S40: Sensitivity analysis - forest plot of rs4727799 95
Figure S41: Sensitivity analysis - region plot of rs73581580 96
Figure S42: Sensitivity analysis - forest plot of rs73581580 97
Figure S43: Sensitivity analysis - Quantile–Quantile plot of the gene-based analysis of subset excluding subjects with schizophrenia or bipolar disorder (Ncases = 8,618, Ncontrols = 1,027,690) 98
Figure S44: GTEx tissue enrichment expression of genes associated with BPD as implemented in FUMA 99
Figure S45: BrainSpan age groups: enrichment of expression of genes associated with BPD as implemented in FUMA 99
Figure S46: BrainSpan developmental stages: enrichment of expression of genes associated with BPD as implemented in FUMA 100
Figure S47: SNP-h² enrichment for BPD on the supercluster-level based on Human Brain Atlas single-nucleus RNA sequencing data 101
Figure S48: Drug target analysis 102
Figure S49: Genetic correlations of BPD with other phenotypes 103

### Supplementary text

#### Borderline personality disorder symptoms

The Diagnostic and Statistical Manual of Mental Disorders (DSM-5) requires the presence of five of nine possible criteria for the diagnosis of BPD: 1) frantic efforts to avoid real or imagined abandonment, 2) a pattern of unstable and intense interpersonal relationships, 3) identity disturbance, 4) impulsivity in at least two areas that are potentially self-damaging (e.g., spending, sex, substance abuse, reckless driving, binge eating), 5) recurrent suicidal or non-suicidal self-injuring behavior, 6) affective instability, 7) chronic feelings of emptiness, 8) inappropriate, intense anger or difficulty controlling anger, and 9) transient, stress-related paranoid ideation or severe dissociative symptoms (Association and American Psychiatric Association 2013; Leichsenring et al. 2023).

#### Genotyping, quality control, and imputation

Samples providing individual-level genotype data were grouped into batches based on array and ancestry, resulting in five batches (see Supplementary Tables S1&S2). Quality control and imputation were carried out using the RICOPILI GWAS pipeline (Lam et al. 2019).

Briefly, the exclusion criteria for SNPs and subjects in the first round of quality control were: genotyping call rate for given SNPs or individuals <98%, difference in SNP genotyping call rate between cases and controls >2%, deviation of autosomal heterozygosity from the mean (|Fhet|>0.2), or a deviation from Hardy-Weinberg equilibrium (HWE; $p<1\times10^{-10}$ in cases; $p<1\times10^{-6}$ in controls). Imputation was conducted using a publicly available reference panel consisting of 54,330 phased haplotypes with 36,678,882 variants from the haplotype reference consortium (EGAD00001002729) and the prephasing/imputation stepwise approach in EAGLE/MINIMAC3 (default parameters and a variable chunk size of 132 genomic chunks) (Das et al. 2016; Loh, Palamara, and Price 2016). We applied an additional round of the above QC to the X-chromosomal genotypes, analyzing male and female subgroups separately. SNPs deviating from HWE ($p<1\times10^{-6}$), those with a missing genotype rate <98%, and those with a minor allele frequency <5% were excluded.

Relatedness testing and population structure analysis were performed using a subset of 55,001 SNPs that fulfilled strict quality criteria after imputation (INFO>0.8, missingness<1%, minor allele frequency>0.05), and which had been subjected to LD pruning ($r^2>0.02$) in the second round of quality control. In the case of cryptically related subjects with pi-hat >0.2, one member of each pair was removed at random following the preferential retention of cases over controls. Twenty genetic principal components (PCs) were estimated from the quality-controlled genotypic data, and their association was tested using logistic regression. PCs 1, 2, 3, 4, 5, 6, 8, 10, 12, 14, 15, and 20 were ultimately employed in the downstream analysis to control for population stratification. The effect of individual PCs on genome-wide test statistics was also assessed using genomic inflation factor (λ).

#### Description of studies providing individual-level data

##### Witt2017

**PI**
Witt et al., 2017

**PMID**
28632202

**Analysis Code**
boma3

**Sample Description**

This sample has been reported as the first wave of the Borderline Personality Disorder GWAS (Witt et al. 2017). Patients were recruited from the following German academic institutions: Department of Psychosomatic Medicine, Central Institute of Mental Health, Mannheim; Department of Psychiatry and Psychotherapy, University Medical Centre Mainz; and Department of Psychiatry, Charité, Campus Benjamin Franklin, Berlin. The control sample consisted of unscreened blood donors from Mannheim and controls recruited from the University Medical Centre Mainz.

**Case Ascertainment**

Cases were included if they fulfilled the criteria for a lifetime diagnosis of borderline personality disorder according to DSM-IV criteria, were 16–65 years old, and had Central European ancestry. The diagnosis was on the basis of a structured clinical interview (German version of the IPDE or SCID-II). Diagnostic interviews were conducted by trained and experienced raters. Cases with a comorbid diagnosis of bipolar disorder or schizophrenia assessed with the SCID-I were excluded.

**Control Ascertainment**

Controls were included if they were at least 18 years or older. Controls from Mainz were screened for a list of psychiatric disorders (panic disorder, agoraphobia, social phobia, specific phobia, generalized anxiety disorder, PTSD, obsessive-compulsive disorder, major depression, dysthymia, mania, hypochondriacal disorder, somatoform disorder, pain, conversion disorders, anorexia nervosa, bulimia nervosa, harmful alcohol use, alcoholism, harmful drug use, drug addiction, schizophrenia, and schizotypal disorders). Controls from Mannheim were blood donors who did not meet the exclusion criteria for blood donation: <50 kg, concurrent general medical conditions, and an elevated risk for infection. Subjects were handed a questionnaire assessing demographic information and information about their mental and somatic health at the blood donation event which they then mailed back to the study center. However, the screening answers were not used for control ascertainment.

**Genotyping**

Automated genomic DNA extraction was performed using the chemagic Magnetic Separation Module I (Chemagen Biopolymer-Technologie, Baesweiler, Germany). Genotyping was performed using the Infinium PsychArray-24 Bead Chip (Illumina, San Diego, CA, USA).

**Funding**

The study was supported by the German Federal Ministry of Education and Research (BMBF) through the Integrated Network IntegraMent (Integrated Understanding of Causes and Mechanisms in Mental Disorders), under the auspices of the e:Med Programme (grant 01ZX1314A to MMN and SC; grant 01ZX1314G to MR). The study was supported by the German Research Foundation (DFG; grant FOR2107; RI908/11-1 to MR; WI3429/3-1 to SHW; NO246/10-1 to MMN; DA1151/5-1 to UD; KFO 256 BO 1487/12-1 to MB;.SFB 779 TP A08 to BHS).

**Ethics Statement**

All participants provided written information consent before participation. The study was approved by the local ethics committees.

#### KFO256

**PI**

Martin Bohus, Christian Schmahl

**PMID**

26401296

**Analysis Code**

borge

**Sample Description**

Recruitment was carried out by the central project of the KFO256, which is a clinical research unit funded by the German Research Foundation (DFG) dedicated to investigating mechanisms of disturbed emotion processing in BPD (Schmahl et al. 2014). Assessment included a range of instruments (SCID; IPDE-BPS; ZAN-BPS; SVV-Checkliste; GAF; ADHS-SB; AQ; BDI; BIS; BSL; CAARS; CTQ; DERS; FDS; HEXACO; SCL-90-R; STAI; STAXI; WURS-k; Persönlichkeitsinventar für DSM-5; SPF).

**Case Ascertainment**

The diagnosis according to DSM-IV was made by trained clinical psychologist using the International Personality Disorder Examination (IPDE), a semi-structured clinical interview assessing personality disorders. Patients met at least five diagnostic criteria for BPD as defined by DSM-IV. Exclusion criteria were a lifetime history of bipolar affective disorder or psychotic disorder; mental deficiency and developmental disorders; pregnancy; significant somatic diseases; psychotropic medication within last 2 weeks; current substance abuse disorder (last 6 months); current suicidal crisis

**Control Ascertainment**

Exclusion criteria for controls were a lifetime history of mental disorders and/or significant somatic disorders or pregnancy.

**Genotyping**

DNA was extracted from peripheral blood samples using automated DNA extraction with the chemagic Magnetic Separation Module I (Chemagen Biopolymer-Technologie, Baesweiler, Germany). All samples were genotyped using Illumina InfiniumGlobal Screening Arrays (Illumina, San Diego, CA, USA).

**Funding**

We thank all participants involved in the study. This project was funded the German Research Foundation (KFO256).

**Ethics Statement**

Ethics Committee of the Medical Faculty Mannheim of the University of Heidelberg (reference: 2011-22N-MA)

#### BerlinCharité

**PI**
Stefan Röpke

**PMID**
22686225

**Analysis Code**
borge

**Sample Description**

The sample consists of hospitalised BPD patients recruited at Charité – Universitätsmedizin Berlin, corporate member of Freie Universität Berlin and Humboldt-Universität zu Berlin, Campus Benjamin Franklin,Department of Psychiatry and Neurosciences, Berlin, Germany.

**Case Ascertainment**

BPD patients (18 years and older) were diagnosed using the SCID-IV interview and met at least five diagnostic criteria for BPD according to the DSM-IV definition. The diagnostic interviews were conducted by trained and experienced assessors. All study participants were further characterised using the Symptom Checklist 90 (SCL-90-R) (Franke and Derogatis 2002) and the Borderline Symptom List 23 (BSL-23) (Wolf et al. 2009). Additional questionnaires recorded socio-demographic information.

**Control Ascertainment**
Not applicable

**Genotyping**
DNA was extracted from whole blood. Genome-wide genotyping was performed using Global Screening Array (Illumina, Inc., San Diego, CA, USA) at the Life&Brain facilities, Bonn, Germany.

**Funding**
None

**Ethics Statement**

The study was approved by the Ethics Committee of Charité - Universitätsmedizin Berlin, Berlin, Germany, and was conducted in accordance with the Declaration of Helsinki. Written informed consent was obtained from all study participants.

#### Tuebingen

### PI

Vanessa Nieratschker

##### PMID

31587837, 29274998, 30134995

##### Analysis Code

borge

##### Sample Description

The sample consists of hospitalized BPD patients and healthy control individuals without any history of psychiatric disorders recruited at the Department of Psychiatry and Psychotherapy, Tuebingen. All subjects were of Caucasian origin and groups were matched for age and sex.

##### Case Ascertainment

BPD patients (18 years and older) were diagnosed according to the International Personality Disorder Examination (IPDE) and met at least five diagnostic criteria for BPD as defined in DSM-IV. Diagnostic interviews were conducted by trained and experienced raters. All study participants were further characterized using the Symptom Checklist 90 (SCL90R) (Franke and Derogatis 2002), the Borderline Symptom List 23 (BSL23) (Wolf et al. 2009) and the Childhood Trauma Questionnaire (Bernstein et al. 2003). Additional questionnaires assessed demographic information along with information about nicotine and alcohol consumption (Audit (Saunders et al. 1993) and Fagerstrom-Test (Fagerstrom and Schneider 1989)). GSI (global severity index = average rating given to all items) and PST scores (positive symptom total = number of symptoms/items rated higher than zero) were calculated from the SCL90R.

##### Control Ascertainment

Controls were included if they were at least 18 years or older. Controls were screened for psychiatric disorders and only included if they did not present a psychiatric history. All study participants were further characterized using the Symptom Checklist 90 (SCL90R) (Franke and Derogatis 2002), the Borderline Symptom List 23 (BSL23) (Wolf et al. 2009) and the Childhood Trauma Questionnaire (Bernstein et al. 2003). Additional questionnaires assessed demographic information along with information about nicotine and alcohol consumption (Audit (Saunders et al. 1993) and Fagerstrom-Test (Fagerstrom and Schneider 1989)). GSI (global severity index = average rating given to all items) and PST scores (positive symptom total = number of symptoms/items rated higher than zero) were calculated from the SCL90R.

##### Genotyping

Venous blood was drawn from all subjects, collected in Ethylenediaminetetraacetic acid (EDTA) tubes and stored at -80°C until further analysis. DNA extraction was performed using the QIAamp DNA Blood Maxi-Kit (Qiagen, Hilden, Germany).

##### Funding

This work was funded by a NASARD Young Investigator grant (23494) from the Brain & Behavior Research Foundation to VN, a grant from the German Research Foundation (DFG) to VN (NI1332/16-1), and by an IZKF grant (PK2015-1-11) to VN and NK.

**Ethics Statement**

The study was approved by the ethics committee of the University of Tuebingen and was conducted in accordance with the Declaration of Helsinki. Written informed consent was obtained from all study participants.

#### Basel

**PI**
Sebastian Euler, Sven Cichon

**PMID**
33619789

**Analysis Code**
borge

**Sample Description**
The sample is a subsample of a larger study investigating clinical and neurobiological aspects of inpatients with BPD. Patients were included after admission to a disorder-specific treatment. Before treatment, all patients had two initial outpatient diagnostic assessments. Exclusion criteria were substance abuse 1 week before admission, psychotic symptoms, intellectual disability and age below 18.

**Case Ascertainment**
Diagnosis was confirmed according to SCID-II. Additional questionnaires assessed demographic information along with clinical information including SCID I, STAXI, BDI, RSQ, BIS-11, BSL-23, PNI-54, and IPO.

**Control Ascertainment**
none

**Genotyping**
Automated genomic DNA extraction was performed using the chemagic Magnetic Separation Module I (Chemagen Biopolymer-Technologie, Baesweiler, Germany). Genotyping was performed using the Infinium Global Screening Array (GSAMD24v2-0_20024620_A1, Illumina, San Diego, CA, USA).

**Funding**
The study was funded by the research funding pool of the Psychiatric University Hospital Basel.

**Ethics Statement**
The study was approved by the ethics committee of north western and central Switzerland and was conducted in accordance with the declaration of Helsinki. Written informed consent was obtained from all study participants.

#### Geneva

**PI**
Nader Perroud

**PMID**
22832351, 26350166

**Analysis Code**
borge

**Sample Description**
BPD were recruited in an outpatient unit specialized in the assessment and treatment of borderline personality disorder (BPD) in the University Hospitals of Geneva, in Switzerland. Patients are usually referred by their general practitioner, psychiatrist, psychologist, or other mental health care professional for suicidal or deliberate self-harm behaviors. All subjects were of Caucasian origin. The study protocol was approved by the Ethic Committee of the Republic and Canton of Geneva and all subjects gave their informed written consent.

**Case Ascertainment**
BPD patients (18 years and older) were diagnosed using the Screening Interview for Axis II Disorders BPD part and met at least five diagnostic criteria for BPD as defined in DSM-IV. Diagnostic interviews were conducted by trained and experienced raters. In addition, all participants were assessed for Axis I psychiatric diagnoses using the French version of the Diagnostic Interview for Genetic Studies (DIGS). Additional questionnaires assessed demographic information along with information about current level of depression using the Beck Depression Inventory II and the Childhood Trauma Questionnaire (CTQ) to assess history of childhood trauma. Other questionnaires assessing anger (STAXI) and impulsivity (BIS-10) were also used.

**Control Ascertainment**
-

**Genotyping**
Venous blood was drawn from all subjects, collected in Ethylenediaminetetraacetic acid (EDTA) tubes and stored at -80°C until further analysis. DNA was extracted from blood white cells (Nucleon extraction kit, Amersham Life Science, Dubendorf, Switzerland).

**Funding**
Swiss National Center of Competences in Research “Synapsis Basis of Mental Diseases” N° 51NF40-158776.

**Ethics Statement**
The study was approved by the ethics committee of the Republic and Canton of Geneva and was conducted in accordance with the Declaration of Helsinki. Written informed consent was obtained from all study participants.

#### MultiRCT

**PI**

Arnoud Arntz

**PMID**

25407009, 35234828

**Analysis Code**

borge

**Sample Description**

BPD participants were recruited for an RCT into group schema therapy in outpatient mental health institutes in Australia, Germany, Greece, the Netherlands, and the UK. Controls were recruited in Germany and the Netherlands.

**Case Ascertainment**

Cases were included if they had a primary diagnosis of BPD (DSM-IV), age of 18 to 65 years, a BPD-severity score assessed with the BPDSI-IV > 20, and expressed willingness & practical ability to participate in 2 years of research and treatment.

The diagnosis was made on the basis of a structured clinical interview (SCID-I and SCID-II). Diagnostic interviews were conducted by trained and experienced raters.

The following exclusion criteria were used: inability to speak, read, and/or understand the study site's language, IQ less than 80, a psychotic disorder (except reactive episodes; BPD criterion 9 per the DSM-5), bipolar disorder 1, dissociative identity disorder, untreated attention deficit/hyperactivity disorder, addiction needing clinical detoxification (inclusion after detoxification was allowed), full or subthreshold narcissistic or antisocial PD, a serious and/or unstable medical illness, and having received ST for more than 3 months during the previous 3 years.

**Control Ascertainment**

Controls were women, age of 18 to 65, who reported to have never had a mental disorder (lifte-time) and according to SCID-I and SCID-II did not meet any diagnosis. They were matched on (estimated) IQ to the cases. They were excluded when unable to speak, read, and/or understand the study site's language, or had IQ < 80.

**Genotyping**

DNA was either collected with DNA Genotek Oragene Kits and extracted with the Oragene DNA purifier OG-L2P, in accordance with the Oragene DNA protocol (DNA Genotek Inc.), or collected with a mouthwash solution, and extracted using an automated system (QIAsymphony, Qiagen) according to the manufacturer's protocol. All samples were genotyped on the Illumina InfiniumGlobal Screening Array (Illumina, San Diego, CA, USA).

**Funding**

Funding/Support: The sites in the Netherlands were supported by ZonMW grant 80-82310-97-12142 from the Netherlands Organization for Health Research and Development (Dr Arntz) and grant 2008 6350 from the Netherlands Foundation for Mental

Health (Dr Arntz). The sites in Germany were supported by Else Kröner-Fresenius-Stiftung (Dr Jacob). The sites in Australia were supported by Australian Rotary Health (Dr Lee). The site in Greece was supported by the Greek Society of Schema Therapy, the First Department of Psychiatry of the Medical School of the University of Athens, and the Institut für Verhaltenstherapie Ausbildung Hamburg. The site in London, UK, was supported by the South London and Maudsley NHS Foundation Trust and by Research Center Experimental Psychopathology, Maastricht University. The site in Bradford, UK, was supported by the Bradford District Care NHS Foundation Trust.
Role of the Funder/Sponsor: The funders had no role in the design and conduct of the study; collection, management, analysis, and interpretation of the data; preparation, review, or approval of the manuscript; and decision to submit the manuscript for publication.

**Ethics Statement**

All participants provided written information consent before participation. The study was approved by the local ethics committees: Medical Ethical Committee Academic Hospital Maastricht & Maastricht University, the Netherlands (reference METC 09-3-044); Murdoch University Human Research Ethics Committee, Australia (reference 2010/217); NRES Committee London - Camberwell St Giles, UK (reference 13/LO/1631); Ethik-Kommission Albert-Lüdwigs-Universität Freiburg, Germany (reference 303/09); Ethik-Kommission Universität zu Lübeck, Germany (reference 09-094); Ethics Committee and Committee for the Human Rights of Eginition Hospital Athens, Greece (reference 185, 20/3/2014)

#### Hope

**PIs**
James Kennedy; Shelley McMain

**PMIDs**
19755574; 22581157

**Analysis Code**
borge

**Sample Description**
Genetic samples were obtained from individuals that consented to participate in a single-blind, randomized controlled trial originally designed to compare the clinical effectiveness of 1-year dialectical behaviour therapy (DBT) to general psychiatric management. Patients were recruited from the Centre for Addiction and Mental Health (CAMH) and St. Michael's Hospital in Toronto, Ontario, Canada, both affiliated with the University of Toronto, between July 2003 and April 2006.

**Case Ascertainment**
To be included in the study, patients had to meet DSM-IV criteria for BPD, be 18-60 years of age, and have had at least two episodes of suicidal or non-suicidal self-injurious behaviour in the past five years, at least one of which occurred in the three months preceding enrollment. Diagnostic interviews were conducted by experienced, doctoral-level clinicians or board-certified psychiatrists who used the SCID-I and IPDE.

Individuals were excluded if they had a DSM-IV diagnosis of a psychotic disorder, bipolar I, delirium, dementia, intellectual disability, or substance use; had a medical condition that precluded psychiatric medications; lived outside a 40-mile radius of Toronto; had a serious medical condition likely to require hospitalization within the next year; or had plans to leave the province within two years.

**Control Ascertainment**
This cohort does not contain any controls.

**Genotyping**
DNA was extracted using a non-enzymatic, high-salt method. Genotyping was performed using the Global Screening Array (Illumina, Inc., San Diego, CA, USA).

**Funding**
The RCT was funded by the Canadian Institutes for Health Research Grant (200204MCT-101123).

**Ethics Statement**
Protocols were approved by the CAMH and St. Michael's Hospital Research Ethics Boards, and written informed consent was obtained prior to participant enrollment and sample collection.

#### FASTER

**PIs**
James Kennedy; Shelley McMain; Alexander Chapman

**PMIDs**
30016935; 35738244

**Analysis Code**
borge

**Sample Description**
Samples were obtained from individuals that consented to participate in a single-blind, randomized controlled trial designed to compare the clinical and cost effectiveness of 6 versus 12 months of dialectical behaviour therapy (DBT) for chronically suicidal individuals with BPD. Individuals were recruited from existing treatment or research wait-lists at the BPD Clinic at the Centre for Addiction and Mental Health (CAMH) in Toronto, Ontario and the DBT Centre at Simon Fraser University (SFU) in Vancouver, British Columbia, Canada, as well as through advertisements at hospitals, universities, health service centres, and word-of-mouth referrals between 2015 and 2018. In September of 2015, an amendment to the original study protocol was approved that incorporated a genetic component for the collection of saliva samples.

**Case Ascertainment**
Individuals aged 18-65 were eligible to participate if they met DSM-IV criteria for BPD based on the IPDE, exhibited recent and chronic self-injurious behaviours (at least two episodes of self-injury or suicide attempts in the past five years, including one in the past eight weeks), were proficient in English, had not received more than eight weeks of DBT in the previous year, and had health insurance coverage for one year or more. Following a telephone pre-screen, diagnostic interviews were conducted by experienced and trained study assessors.

Participants were excluded if they met criteria for a specific psychotic disorder, bipolar disorder I, or dementia according to DSM-IV criteria, had an estimated IQ less than or equal to 70, had a chronic or serious physical health problem expected to require hospitalization in the next year, or had plans to move out of the province within two years.

**Control Ascertainment**
This cohort does not contain controls.

**Genotyping**
Saliva samples were collected using Oragene DNA kits (DNA Genotek Inc.) and DNA was extracted using an automated, non-enzymatic, high-salt method. Genotyping was performed using the Global Screening Array (Illumina, Inc., San Diego, CA, USA).

**Funding**
This study was funded by the Canadian Institutes of Health Research Grant (CIHR FRN 133428) and the Larry and Judy Tanenbaum Foundation.

**Ethics Statement**
Ethics approval was granted by the research ethics board at CAMH on May 15, 2014 (#026/2014) and at SFU on August 28, 2015 (#2014 s0263). In September 2015, an

amendment was approved for recruitment from the existing study to participate in the genetics sub-project, which had a separate consent form and process.

#### SleepCIMH

**PI**
Claudia Schilling; Michael Schredl

**PMID**
29325115

**Analysis Code**
borge

**Sample Description**
This sample consists of healthy control subjects participating in the study „Activity of stress systems, metabolic sequelae and genetics of patients with sleep disorders and healthy controls” conducted at the Central Institute of Mental Health (CIMH) in Mannheim, Germany. For this purpose, subjects were screened for eligibility and underwent two nights of polysomnographic investigation and blood sampling. For the present study, we included subjects for which genetic material in the form of a blood sample was available.

**Case Ascertainment**
This sample does not contain any cases.

**Control Ascertainment**
Inclusion criteria: 18–80 years old, German-speaking, Exclusion criteria: Any sleep disorder, history of any somatic or psychiatric disorder or treatment interfering with sleep investigation.

**Genotyping**
DNA was extracted from whole blood. Genome-wide genotyping was performed using Global Screening Array 1.0 (Illumina, Inc., San Diego, CA, USA) at the Life&Brain facilities, Bonn, Germany.

**Funding**
None.

**Ethics Statement**
The study was approved by the Ethics Committee of the Medical Faculty Mannheim of the University of Heidelberg, and all subjects provided written consent (reference: 2011-315N-MA).

#### Barcelona

**PI**
Juan Carlos Pascual Mateos

**PMID**
33414392

**Analysis Code**
borsm

**Sample Description**
Patients were enlisted from the Borderline Personality Disorder (BPD) outpatient units of two hospitals (Hospital de la Santa Creu I Sant Pau and the Consorci Sanitari de l'Anoia, Barcelona). Healthy controls were recruited from the general population. Several clinical instruments were used for assessment: Structured clinical interview for DSM IV axis II personality disorders (SCID II), Revised Diagnostic Interview for Borderlines (DIB-R), McLean Screening Instrument for Borderline Personality Disorder (MSI-BPD).

**Case Ascertainment**
BPD patients (18–50 years) met at least five diagnostic criteria for BPD as defined in DSM-IV.

**Control Ascertainment**
Healthy control participants were mandated to have no record of mental health disorders, substance use, or treatment with psychotropic drugs.

**Genotyping**
DNA was extracted from whole blood. Genome-wide genotyping was performed using Global Screening Array 1.0 (Illumina, Inc., San Diego, CA, USA) at the Life&Brain facilities, Bonn, Germany.

**Funding**
This work was supported by the Centro de Investigación Biomédica en Red de Salud Mental (CIBERSAM), the Instituto de Salud Carlos III [PI21/00505] and co-financed by the European Regional Development Fund (ERDF).

**Ethics Statement**
The study was approved by the Ethical Board: Hospital de la Santa Creu I Sant Pau Clinical Research Ethics Committee.

#### SpainControls

### PI

Marta Ribases and Josep Antoni Ramos-Quiroga

##### PMID

32279069

##### Analysis Code

borsm

##### Sample Description and Control Ascertainment

A total of 678 controls from the Mental-Cat cohort (74.6% female) were evaluated and recruited prospectively from a restricted geographic area at the Hospital Universitari Vall d'Hebron of Barcelona (Spain) and consisted of unrelated healthy blood donors.

##### Genotyping

Genomic DNA was obtained from peripheral blood lymphocytes by the salting out procedure. DNA concentrations were determined using the Pico-Green dsDNA Quantitation Kit (Molecular Probes, Eugene, OR) and genotyped with the Ilumina Infinium™ Global Screening Array-24 v2.0 (GSA).

##### Funding

This work was supported by the Agència de Gestió d'Ajuts Universitaris i de Recerca (AGAUR, 2021SGR-00840), the Instituto de Salud Carlos III (PI20/00041), the Biomedical Network Research Centre on Mental Health (CIBERSAM) and the European Regional Development Fund (ERDF).

##### Ethics Statement

The study was approved by the Clinical Research Ethics Committee (CREC) of Hospital Universitari Vall d'Hebron, all methods were performed in accordance with the relevant guidelines and regulations and written informed consent was obtained from participants before inclusion into the study.

#### POSEIDON

**PI**
Michael Deuschle

**PMID**
30471571

**Analysis Code**
borge

**Sample Description**
This sample consists of parents of the families participating in the longitudinal study POSEIDON (Pre-, Peri-, and Postnatal Stress: Epigenetic impact on Depression) for whom genetic data was available. Pregnant women were recruited from hospitals in the Rhine-Neckar Region of Germany about 4–8 weeks prior to delivery. Data was collected during the third trimester of pregnancy (T1) and 45 months postpartum (T4). The last assessment (T4) took place between August 2014 and January 2017. At T4, dropouts were replaced by families with children of the same age as the original POSEIDON cohort. For this purpose, information about eligible new participants was obtained from the local registries of residents (birth year of the child, parents' names and address). The eligible new participants were informed about the study by mail. For the present study, we included parents for which genetic material in the form of a saliva sample was available.

**Case Ascertainment**
This sample does not contain any cases.

**Control Ascertainment**
Mothers were contacted during pregnancy at the hospital. The subjects replacing dropouts at T4 were contacted via local registries. Maternal inclusion criteria: 16–45 years old, German-speaking, and presumably the child's main caregiver. Maternal exclusion criteria: positive for hepatitis B, hepatitis C or human immunodeficiency virus, current psychiatric disorder requiring inpatient treatment, history or current diagnosis of schizophrenia or psychotic disorder, substance dependency other than nicotine during pregnancy. No paternal exclusion criteria were applied.

**Genotyping**
DNA was extracted from saliva using the Oragene DNA purifier OG-L2P, in accordance with the Oragene DNA protocol (DNA Genotek Inc.). Genotyping for the parents was performed with the Infinium Global Screening Array-24 Multi-Disease Bead Chip version 3.0 (Illumina, San Diego, CA, USA).

**Funding**
The POSEIDON study was supported by the German Federal Ministry of Education and Research (BMBF) through the Integrated Network IntegraMent, under the auspices of the e:Med Programme [01ZX1314G; 01ZX1614G] and through ERA-NET NEURON “*Impact of Early life MetaBolic and psychosocial strEss on susceptibility to mental Disorders; from converging epigenetic signatures to novel targets for therapeutic intervention*” [01EW1904], and by a grant of the Dietmar-Hopp Foundation.

**Ethics Statement**
The study was approved by the Ethics Committee of the Medical Faculty Mannheim of the University of Heidelberg, and all families provided written consent (reference: 2014-550N-MA).

#### Munich

**PI**

Richard Musil

**PMID**

n.a.

**Analysis Code**

borge

**Sample Description**

Patients were recruited on the specialized ward for Borderline Personality Disorder of the Psychiatric clinic of the University (LMU) of Munich. All patients took part in the Munich Mental Health Biobank. Some of the patients also took part in the DBT-CBASP-Study.

**Case Ascertainment**

Patients had to fulfill 5 out of 9 DSM criteria, ascertained via SCID-II or SCID-5-PD.

**Control Ascertainment**

This sample does not contain any controls.

**Genotyping**

DNA was extracted from whole blood. Genome-wide genotyping was performed using Global Screening Array 1.0 (Illumina, Inc., San Diego, CA, USA) at the Life&Brain facilities, Bonn, Germany.

**Funding**

The study had no specific funding.

**Ethics Statement**

The study was approved by the local Ethics Committee of the Medical Faculty of LMU University, Munich (DBT-CBASP-Study, PI Frank Padberg, IRB number 713-15, German Clinical Trials Register: DRKS00019821).

#### Oslo

**PI** Benjamin Hummelen, Ole Andreassen

**PMID**
36409693

**Analysis Code** boro1 (Oslo1); boro2 (Oslo2)

**Sample Description**
This sample consisted of both cases and control persons. Patients were recruited from the Norwegian Network for Personality Disorders, a clinical research network in Southern Norway consisting of clinical units specialized in the assessment and treatment of patients with different types of personality disorders (Pedersen et al. 2023). Four units participated in the Genetics project. Control subjects were provided by the Norwegian Thematic Organized Psychosis Research Study (TOP Study), consisting of randomly selected participants from statistical records from the same catchement area as the cases and of Caucasian origin.

**Case Ascertainment**
All cases were assessed by the Structured Clinical Interview for DSM-IV Axis Personality Disorders (SCID-II), administered by experienced clinicians. In order to be included in the current study, patients had to meet at least five BPD criteria. Symptom disorders were assessed using the Mini-International Neuropsychiatric Interview (MINI).

**Control Ascertainment**
Control subjects were evaluated with a clinical interview about severe mental disorder symptoms and the Primary Care Evaluation of Mental Disorders (PRIME-MD). The subjects were excluded if they or any of their close relatives had a lifetime history of a severe psychiatric, if they had an unstable medical condition known to interfere with brain function, neurological disorder, IQ<70 or history of severe head trauma as well as significant illicit drug use.

**Genotyping**
*Oslo1:* Cases and controls were genotyped on the Human OmniExpress chip (Illumina Inc., San Diego, 135 CA, USA) in accordance with the standard Illumina protocol conducted by deCODE Genetics (Reykjavik, Iceland).
*Oslo2:* Cases and controls were genotyped on the DeCODE Genetics V1, a custom version of the Global Screening Array (Illumina Inc., San Diego, CA, USA) in accordance with the standard Illumina protocol, conducted by deCODE Genetics (Reykjavik, Iceland).

**Funding**
Oslo University Hospital, Norwegian Research Council, University of Oslo.

**Ethics Statement**
The study was approved by the Regional Committees for Medical and Health Research Ethics (reference 12656). All participants provided written consent before participating in the study.

**Acknowledgements**
We wish to thank the patients, staff and research coordinators from the Norwegian Network for Personality Disorders for their contribution to this study. The collaboration includes the following units: Unit for Group Therapy, Øvre Romerike District Psychiatric Center, Akershus University Hospital, Jessheim, Norway; Group Therapy Unit, Follo District Psychiatric Center, Akershus University Hospital, Ski, Norway; Clinic for Personality disorders, Outpatient Clinic for Specialized Treatment of Personality Disorders, Section for Personality psychiatry and specialized treatments, Oslo University Hospital, Oslo, Norway; Group Therapy Unit, Lovisenberg District Psychiatric Center, Lovisenberg Hospital, Oslo.

#### Montreal

**PI**
Gustavo Turecki

**PMID**
17503981, 16263852

**Analysis Code**
boro2

**Sample Description**
This sample consists of postmortem human brain tissue obtained as part of the activities of the Douglas Bell-Canada Brain Bank (douglasbrainbank.ca). Collection of brain tissue is obtained in collaboration with Quebec's Coroner Office from individuals who died suddenly without agonal periods. Tissue is obtained following consent from next-of-kin, and subsequently, donors are clinically characterized following proxy-based, structured procedures adapted for best-informants and known as psychological autopsies.

**Case Ascertainment**
In this study we used samples from individuals meeting criteria for borderline personality disorder according to DSM-IV criteria as ascertained by SCID II interviews, adapted for proxy-based interviews (McGirr et al. 2007).

**Control Ascertainment**
Controls were individuals without major psychiatric disorders and not meeting criteria for borderline personality disorder according to DSM-IV criteria and based on SCID-I and SCID-II interviews.

**Genotyping**
DNA was extracted from brain tissue samples. Genotyping was performed with the Infinium Global Screening Array-24 (Illumina, San Diego, CA, USA).

**Funding**

The Douglas Bell-Canada Brain Bank is funded by the Fonds de Recherche en Santé du Québec (FRSQ), as well as by Brain Canada and Healthy Brain for Healthy Living (HBHL; Canada First Research Excellence Fund - CFREF).

**Ethics Statement**

The activities of the Douglas Bell-Canada Brain Bank are regulated through an authorization of the Douglas Institute Ethics Research Board for tissue banking.

### Description of studies providing summary statistics

#### BioVU

### PI

Lea K Davis

##### PMID

18500243

##### Analysis Code

bioVU

##### Sample Description

The Vanderbilt University Medical Center (VUMC) EHR was established in 1998 (Geissbühler et al. 1998) and houses the medical records of 3.2 million individuals. The Synthetic Derivative (SD) database is a de-identified mirror image of the VUMC EHR and includes data on billing codes from the International Classification of Diseases, 9th and 10th editions (ICD-9 and ICD-10), demographics, Current Procedural Terminology (CPT) codes, laboratory values, clinical documentation, and prescription medications. In 2007 VUMC launched a biobank (BioVU) that stores DNA from over 250,000 VUMC patients, and links genotype data to the SD (Roden et al. 2008).

##### Case Ascertainment

In this study we used samples from individuals meeting criteria for borderline personality disorder ICD9/10 codes (N=315 cases; see Supplementary tables for specific criteria).

##### Control Ascertainment

3,150 controls were matched to cases based on sex, race, ethnicity, and median age of record. The inclusion and exclusion criteria are described in the Supplementary tables.

##### Genotyping, Quality Control, Imputation, and Analysis

Genotype data for the BioVU population was generated using the Illumina Multi-Ethnic Genotype Array ($MEGA^{EX}$). The genotype data were imputed into the HRC reference panel using the Michigan imputation server. This set was restricted to the individuals who clustered with those of European ancestry based on principal component analysis of genotype data. For more information see (Dennis et al. 2021). GWAS was performed using SAIGE version 0.42.1, including the top 10 PCs as fixed effect covariates.

##### Funding

The dataset(s) used for the analyses described were obtained from Vanderbilt University Medical Center's BioVU which is supported by numerous sources: institutional funding, private agencies, and federal grants. These include the NIH funded Shared Instrumentation Grant S10RR025141; and CTSA grants UL1TR002243, UL1TR000445, and UL1RR024975. Genomic data are also supported by investigator-led projects that include U01HG004798, R01NS032830, RC2GM092618, P50GM115305, U01HG006378, U19HL065962, R01HD074711; and additional funding sources listed at https://victr.vumc.org/biovu-funding/. LKD was supported by R01 MH113362.

##### Ethics Statement

The project was approved by the Institutional Review Board (IRB) from VUMC (#160302, #172020, #190418).

#### Copenhagen Hospital Biobank (CHB) and the Danish Blood Donor Study (DBDS)

**PI**

CHB: Sisse Rye Ostrowski and Erik Sørensen

DBDS: Sisse Rye Ostrowski, Ole Birger Pedersen, Erik Sørensen and Christian Erikstrup

**PMID**

33169150 (CHB) and 36194120 (DBDS)

**Analysis Code**

danish

**Sample Description**

The Copenhagen Hospital Biobank (CHB) was established in 2009 as a biobank based on surplus material from diagnostic testing in the Danish Capital Region. Patients with a sample in the biobank have been informed about this at admission to hospital and also through a letter sent to them when entering the scientific study with an option to opt out. The present study is based on the Copenhagen Hospital Biobank Pain and Degenerative musculoskeletal disease Study (CHB-PDS) which include >250,000 individuals. The cohort has been linked to Danish health registers to identify diseases that the participants develop over time.

The Danish Blood Donor Study (DBDS) was established in 2010 as a prospective epidemiological research study and biobank using the existing Danish blood bank infrastructure. Upon visits to the blood donation centers, Danish blood donors are invited to participate in the study with informed consent. They are asked to fill in a questionnaire and provide a blood sample for research. Currently we have consent from >160,000 blood donors, have >400,000 questionnaire responses, and >2 million plasma samples on the participants. At the time of inclusion to the study, donors are required to be 18-75 years old, weigh >50 kg, have no risk behavior, and be in overall good health. Like all other Danes the participants develop disease that we can identify through linkage to Danish health registers.

The health register data includes e.g. in- and out-patient hospital visits with ICD-10 codes since 1978 and 1994, respectively, treatment and intervention codes since 1978, prescriptions since 1998, laboratory assay measurements performed at hospitals since 2006.

In total, the summary statistics provided to the meta-analysis were based on the genotypes of individuals with ICD10-F60.3 diagnosis (N=2,379) compared with controls without that diagnosis (N=334,766). The results were obtained by joint analysis of CHB and DBDS cases and controls according to the approval by the Danish National Committee on Health Research Ethics (NVK-1700407 and NVK-1803812).

**Case Ascertainment**

Cases included individuals diagnosed with emotionally unstable personality disorder (ICD-10 code F60.3) according to the National Patient Register (Landspatientregisteret; LPR).

**Control Ascertainment**

Controls included all individuals in the DBDS cohort without emotionally unstable personality disorder (ICD-10 code F60.3) and those individuals in the CHB cohort without emotionally unstable personality disorder available according to the ethical approval according to the National Patient Register (Landspatientregisteret; LPR).

**Genotyping, Quality Control, Imputation, and Analysis**

*Genotyping*: All samples were genotyped at deCODE genetics using the Global Screening Array by Illumina. Whole genome sequencing, chip-typing, quality control, long-range phasing, and imputation from which the data for this analysis were generated was performed at deCODE genetics. The samples were long-range phased together with over 200,000 genotyped samples from North-western Europe using Eagle(Loh et al. 2016). Samples and variants with less than 98% yield were excluded. A haplotype reference panel was prepared in the same manner as for the Icelandic samples (Kong et al. 2008; Gudbjartsson, Helgason, et al. 2015) by phasing whole-genome sequence genotypes of ~15,000 individuals from Scandinavia, the Netherlands, and Ireland using the phased chip data. Graphtyper (Eggertsson et al. 2017) was used to call the genotypes which were subsequently imputed into the phased chip data. Logistic regression was used to test for association between sequence variants and depression. Using the additive model, the expected allele counts were used as a covariate, and the analysis adjusted for sex, whether the individual had been chip-typed and/or sequenced, and the first 20 principal components. LD score regression was used to account for distribution inflation due to cryptic relatedness and population stratification (Bulik-Sullivan et al. 2015) and used the intercept as correction factor (CF = 1.03). The resulting summary statistics contained results for 7,737,239 markers (MAF>1%, imputation info > 0.8).

**Funding**

The DBDS and CHB research infrastructure is funded by the Danish Blood Banks and Bio- and Genome Bank Denmark (DBB). In addition, the studies have received grants from Novo Nordisk Foundation (NNF17OC0027594, NNF23OC0082015) and Rigshospitalet Research Council.

**Ethics Statement**

Both the CHB-PDS and the DBDS were approved by the National Committee on Health Research Ethics and the Capital Region Data Protection Agency (NVK-1803812, NVK-1700407 and P-2019-51, P-2019-99, respectively).

#### deCODE

### PI

Thorgeir Thorgeirsson / Kari Stefansson

##### PMID

25977816

##### Analysis Code

deCODE

##### Sample Description

deCODE genetics (www.decode.com) has, since the year 1996, gathered genotypic and medical data from more than 160,000 volunteer participants in Iceland, comprising well over half of the adult population. Recruitment has been through various projects aimed at specific common disorders, as well as through wider recruitment strategies aimed at studying overall health. Cases (N=294) and controls (N=259,612) used were all Icelandic and were recruited from all over Iceland.

##### Case Ascertainment

Most of the diagnoses were made by clinicians at the Landspitali University Hospital (1987-2019) and were obtained from electronic health records (ICD-10: F60.3). For sensitivity analyses subjects diagnosed with schizophrenia or bipolar disorder were removed from cases using diagnoses from electronic health records at Landspitali University Hospital (ICD9/ICD10).

##### Control Ascertainment

Controls were recruited as a part of various genetic programs at deCODE. The controls were not specifically screened for psychiatric conditions, but subjects ever receiving any ICD10-F6x diagnoses were excluded.

##### Genotyping, Quality Control, Imputation, and Analysis

The Icelandic chip-typed samples (N 166,000) were assayed with the Illumina HumanHap300, HumanCNV370, HumanHap610, HumanHap1M, HumanHap660, Omni-1, Omni 2.5 or Omni Express bead chips at deCODE genetics. Genotypes of single nucleotide polymorphisms (SNPs) and insertions/deletions (indels) were identified and called jointly by Graphtyper (Eggertsson et al. 2017). All samples with a call rate below 98% were excluded from the analysis, and individual SNPs were excluded if there was significant deviation from Hardy-Weinberg equilibrium in the controls, if they produced an excessive inheritance error rate, or if there was substantial difference in allele frequency between chip types. Over 60,000 Icelandic samples have been WGS using GAIIx, HiSeq,X, and NovaSeq Illumina technology. Sample preparation, sequencing methods, alignment and BAM file generation have been described (Gudbjartsson, Sulem, et al. 2015). Two types of imputations from sequence data were performed; into SNP-typed individuals after long-range phasing (Kong et al. 2008) using a method(Gudbjartsson, Sulem, et al. 2015) based on IMPUTE (Marchini et al. 2007), followed by a familial imputation estimating genotype probabilities for about 290,000 un-typed relatives of SNP-typed individuals (Gudbjartsson, Sulem, et al. 2015). The details of the association analysis approach have been described (Gudbjartsson, Sulem, et al. 2015), but logistic regression was used to test for association between SNPs and emotional instability disorder, treating case status as the response and expected genotype counts from imputation or allele counts from direct genotyping as predictor, adjusting for sex, age (or age at death), county of origin, chip-typed and/or sequence status, and an indicator function for the overlap of the lifetime of the individual with the time span of phenotype collection. Testing was performed using the likelihood ratio statistic. To account for confounding biases such as relatedness and stratification within the case and control sample sets we used LD score regression (Bulik-Sullivan et al. 2015) to estimate the correction factor for test statistic inflation due to these biases (CF = 1.06). *P* values were adjusted accordingly. Summary statistics, based on data for 294 cases and 259,612 controls, were filtered on MAF ≥ 1% and imputation information > 0.8, and contained information on ~6.48 million variants.

**Ethics Statement**

All Icelandic studies were approved by the Icelandic National Bioethics Committee and the Data Protection Authority. All subjects signed informed consents prior to donating samples for the genetic study. Personal identifiers of the patients and biological samples were encrypted by a third-party system overseen by the Icelandic Data Protection Authority. Approval for the study was granted by the Icelandic Data Protection Authority and the National Bioethics Committee of Iceland (Approval no: VSNb2016040011/03.01 and VSNb2021090024/03.01).

#### Estonian Biobank

**PI**
Kelli Lehto

**PMID**
38381979, 24518929

**Analysis Code**
EstBB

**Sample Description**
The Estonian Biobank (EstBB) is a population-based biobank cohort that currently comprises >212000 individuals, representing ~20% of the adult population in Estonia. At recruitment, participants sign an informed consent form allowing follow-up linkage of their electronic health records (EHR), thereby providing a longitudinal collection of their phenotypic information. Thus, EstBB is linked to and regularly updated with the national electronic health records, including the national Health Insurance Fund database (available from 2004), Tartu University Hospital (from 2008), and North Estonia Medical Center (from 2005), and various quality registries (causes of death, cancer, etc.). Since >95% Estonian residents are covered with publicly funded universal health care, EstBB holds complete information on diagnoses in ICD-10 coding (both primary and specialist care) as well as drug dispensing data, including drug ATC codes, prescription status and purchase date (if available) for almost all biobank participants.

**Case Ascertainment**
Two types of definitions were used. In Definition I, cases are individuals with two or more ICD-10 diagnoses of F60.3, F60.30, or F60.31 recorded in their electronic health records (EHR). Definition II, used for sensitivity analysis, is similar to Definition I but includes the additional criterion of excluding individuals with any diagnoses coded under ICD-10 F20*–F29* or F31*.

**Control Ascertainment**
Controls are age and gender-matched to cases at a ratio of 1:5. They are selected based on not having any ICD-10 F diagnoses in their EHR.

**Genotyping, Quality Control, Imputation, and Analysis**
Genotyping of DNA samples from the Estonian Biobank was done at the Core Genotyping Lab of the Institute of Genomics, University of Tartu using the Illumina Global Screening Arrays (GSAv1.0, GSAv2.0, and GSAv2.0_EST). Altogether 206,448 samples were genotyped and then PLINK format files were created using Illumina GenomeStudio v2.0.4. During the quality control all individuals with call-rate < 95% or mismatching sex between the genetic (defined based on the heterozygosity of X chromosome) and phenotypic data were excluded.. Variants were filtered by call-rate < 95% and HWE p-value < 1e-4 (autosomal variants only). Variant positions were updated to Genome Reference Consortium Human Build 37, and all variants were changed to the TOP strand based on reference data from the University of Oxford. (https://www.well.ox.ac.uk/~wrayner/strand/).

After QC, the dataset contained 202,910 samples for imputation. Before imputation, variants with MAF<1% and Indels were removed. Prephasing was done using the Eagle v2.3 software (Loh et al. 2016) (number of conditioning haplotypes Eagle2 uses when phasing each sample was set to: --Kpbwt=20000) and imputation was carried out using Beagle v.18May20.d20 (S. R. Browning and Browning 2007; B. L. Browning, Zhou, and Browning 2018) with an effective population size ne=20,000. As a reference, Estonian population specific imputation reference of 2297 WGS samples was used (Mitt et al. 2017). Further, EstBB samples were combined with the 1000 genomes phase 3 dataset for ancestry analysis. Genetic principal components were calculated using a subset of quality controlled and pruned genotyped SNPs. This was further used to identify and remove samples that deviated from the main cluster.

Genome-wide association analysis was performed with SAIGE version 0.43.1.(LOCO parameter == TRUE, --minMAF=0.0001 --minMAC=1) on genome build 37/hg19. Covariates included 10 principal components.

**Funding**
This research was funded by European Union through the European Regional Development Fund Project No. 2014–2020.4.01.15–0012 GENTRANSMED and by the Estonian Research Council grant number PSG615.

**Ethics Statement**
Individual-level data analysis in the EstBB was carried out under the ethical approval 1.1–12/624 from the Estonian Committee on Bioethics and Human Research (Estonian Ministry of Social Affairs).

#### HUNT

### PI

Bendik Winsvold

##### PMID

36777998

##### Analysis Code

hunt

##### Sample Description

The Trøndelag Health Study (HUNT) is a population based cohort study. Inhabitants from the general population in the county of Trøndelag in Norway have been recruited to the HUNT study in four waves (HUNT1 to HUNT4) since the 1980s. For the present study we included genotyped participants 20 years or older when included in the HUNT2 (1995–1997) or HUNT3 (2006–2008) surveys. Participants completed questionnaires, interviews, and measurements to the HUNT databank (https://hunt-db.medisin.ntnu.no/hunt-db/), as well as biological samples to the HUNT biobank (https://www.ntnu.edu/hunt/hunt-biobank). DNA was extracted from whole blood. The Norwegian Identification Number was used to link data from HUNT with local and national registries. We obtained data from local hospital registries on ICD-10 codes from all inpatient and outpatient contacts from 1997 through 2017.

The health care system in Norway is publicly funded. The psychiatric departments at the local hospitals in Nord-Trøndelag, where the HUNT2 and HUNT3 surveys were conducted, had catchment area responsibilities for the whole county. The diagnostic procedure used at most local hospital outpatient clinics the last decades was based on an interview to collect biographic information, mental status examination, and standardized assessment. The standardized assessment implied the Mini International Neuropsychiatric Interview (M.I.N.I.) for symptom diagnoses (Sheehan et al. 1994). Trained therapists conducted the interviews. Furthermore, the Iowa Personality Disorder Screen (IPDS) was used for screening for any personality disorder. The IPDS is a 11-item self-report questionnaire derived from the Structured Interview for the DSM III Personality disorder (SIPD-R) (Stangl et al. 1985). Each item is scored dichotomously (yes/no), giving IPDS scores ranging from 0 to 11 (Langbehn et al. 1999; Olssøn, Sørebø, and Dahl 2011). If there was a suspicion of personality disorder, a Structured Clinical Interview for DSM-IV Axis II Personality Disorders (SCID-II) was completed (First et al. 1995). The SCID-II is a semi-structured interview covering the ten personality disorders as defined in DSM-IV TR (Bell 1994), as well as the entity PD-NOS (not otherwise specified). Within every specific personality disorder, all the criteria were scored separately. All clinicians were trained in performing the SCID-II through local courses. At the end of the assessment all information collected was presented to a team with at least one psychiatrist and one psychologist present to decide diagnoses according to the ICD-10.

##### Case Ascertainment

Cases were defined by at least one contact at local hospitals due to ICD-10 F60.3 emotionally unstable personality disorder.

##### Control Ascertainment

Controls were defined by 1) age >40 years by 2017, 2) no ICD-9 or ICD-10 diagnoses of psychiatric disorders in local hospital registries, 3) no ICPC-2 diagnoses of psychiatric disorders in primary care doctor registries, 4) no self-reported psychiatric disorders in the

HUNT questionnaires, 5) no self-reported daily use of antidepressants, relaxants, or sleeping medications in the HUNT questionnaires, and 6) HADS-A and HADS-D <=11.

**Genotyping, Quality Control, Imputation, and Analysis**

A total of 71,860 participants from the HUNT2 and HUNT3 surveys have been genotyped with Illumina HumanCoreExome arrays (HumanCoreExome12 v1.0, HumanCoreExome12 v1.1, or UM HUNT Biobank v1.0). The genotyping was performed at the Genomics-Core Facility at the Norwegian University of Science and Technology.

We excluded participants whose genotypes had 1) call rates <99%, 2) contamination >2.5%, 3) large chromosomal copy number variants, 4) lowest call rate among technical duplicate pair or twins, 5) uncommon sex chromosomal constellations (i.e. others than XX or XY), or 6) discrepancies with reported sex. The remaining genotypes were analyzed in a second round of genotype calling following the Genome Studio quality control protocol (Guo et al. 2014) [https://www.illumina.com/techniques/microarrays/array-data-analysis-experimental-design/genomestudio.html]. We used BLAT (James Kent 2002) to determine genomic position, strand orientation and reference allele of all genotyped variants, using the Genome Reference Consortium Human genome build 37 [http://genome.ucsc.edu] and the revised Cambridge Reference Sequence of the Human Mitochondrial DNA [http://mitomap.org] as reference. Variants were excluded if they had 1) call rates <99%, 2) higher call rates genotyped in another assay, 3) probe sequences not mapping to the reference genome, 4) cluster separation <0.3, 5) gentrain score < 0.15, or 6) Hardy Weinberg equilibrium deviation from unrelated samples of European ancestry with p<0.0001.

To harmonize the three arrays, we removed variants with frequency differences >15% between the datasets or were monomorphic in one dataset and had MAF > 1% in one of the others. We inferred ancestry, using PLINK v1.90 (Chang et al. 2015), to project the genotype samples into the space of the principal components of the Human Genome Diversity Project reference panel (Li et al. 2008; Wang et al. 2014)[http://csg.sph.umich.edu/chaolong/LASER]. Only individuals of European ancestry were included. Eagle2 v2.3 (Loh et al. 2016) were then used to phase the data. A total of 69,716 samples passed the quality control.

Imputation was performed using the Minimac3 v2.0.1 software (Das et al. 2016) [http://genome.sph.umich.edu/wiki/Minimac3]. For the autosomal variants, we used a customized merged reference panel of 1) 2,201 low-coverage whole-genome sequenced samples from the HUNT study and 2) the Haplotype Reference consortium release 1.1 (HRC v1.1), excluding 1,023 samples from the HUNT study. For the X-chromosome, we used the HRC v1.1 alone. Variants with estimated squared correlations between imputed and true genotypes (Rsq) <0.3 were excluded, resulting in a total of ~25 millions well-imputed variants.

We used the Scalable and Accurate Implementation of GEneralized mixed model (SAIGE) version 0.35.8.3 (Zhou et al. 2018) for testing of associations between binary traits and common genetic variants. The SAIGE method is tailored for GWAS analyses of psychiatric traits in population-based data from a restricted geographic area as it controls for case-control imbalance and relatedness. Sex, batch, and the first 5 principal components were included as covariates. We assumed an additive allelic effect, and included only variants with MAC ≥ 3 in the analysis.

**Funding and acknowledgements**

The Trøndelag Health Study (HUNT) is a collaboration between HUNT Research Centre (Faculty of Medicine and Health Sciences, Norwegian University of Science and Technology NTNU), Trøndelag County Council, Central Norway Regional Health Authority, and the Norwegian Institute of Public Health. The genotyping was financed by the National Institute of health (NIH), University of Michigan, The Norwegian Research council, and Central Norway Regional Health Authority and the Faculty of Medicine and Health Sciences, Norwegian University of Science and Technology (NTNU). The genotype quality control and imputation has been conducted by the K.G. Jebsen center for genetic epidemiology, Department of public health and nursing, Faculty of medicine and health sciences, Norwegian University of Science and Technology (NTNU).

**Ethics Statement**

The GWAS and sharing of summary statistics with international consortia was approved by the Regional Committee for Medical and Health Research Ethics (ref. 2015/575).

#### MoBa (Norwegian Mother, Father and Child Cohort Study)

**PI**

Ted Reichborn-Kjennerud

**PMID**

27063603

**Analysis Code**

moba

**Sample Description**

The Norwegian Mother, Father and Child Cohort Study (MoBa) is a population-based pregnancy cohort study conducted by the Norwegian Institute of Public Health (Magnus et al. 2016). Participants were recruited from all over Norway, at ultrasound examinations around week 17 of pregnancy, from 1999-2008. The women consented to participation in 41% of the pregnancies. The children were born between 1999 and 2009. The cohort includes approximately 114,500 children, 95,200 mothers, and 75,200 fathers. Blood samples were obtained from both parents during pregnancy and from mothers and children (umbilical cord) at birth (Paltiel et al. 2014).

**Case Ascertainment**

Cases were defined as MoBa parents with genotyping data and at least one occurrence of an ICD-10 F60.3 code registered in the Norwegian Patient registry.

**Control Ascertainment**

Controls were defined as MoBa parents matched by sex and genotype batch to cases without any occurrences of ICD-10 F codes in the Norwegian Patient registry. To account for case-control imbalance matched controls were randomly selected so cases represented approximately 5% of the sample size.

**Genotyping, Quality Control, Imputation, and Analysis**

Full details about MoBa genotyping, pre-imputation quality control (QC), phasing, imputation, and post-imputation QC have been previously described (Corfield et al. 2022). Logistic regression modeling was used to conduct case-control genome-wide association analyses using PLINK version 1.90b6.2 (Chang et al. 2015). The first five principal components and genotyping batch were included as covariates.

**Funding**

The Norwegian Mother, Father and Child Cohort Study is supported by the Norwegian Ministry of Health and Care Services and the Ministry of Education and Research. We are grateful to all the participating families in Norway who take part in this on-going cohort study. We thank the Norwegian Institute of Public Health (NIPH) for generating high-quality genomic data. This research is part of the HARVEST collaboration, supported by the Research Council of Norway (#229624). We also thank the NORMENT Centre for providing genotype data, funded by the Research Council of Norway (#223273), South East Norway Health Authorities and Stiftelsen Kristian Gerhard Jebsen.

We further thank the Center for Diabetes Research, the University of Bergen for providing genotype data and performing quality control and imputation of the data funded by the ERC AdG project SELECTionPREDISPOSED, Stiftelsen Kristian Gerhard Jebsen, Trond Mohn Foundation, the Research Council of Norway, the Novo Nordisk Foundation, the University of Bergen, and the Western Norway Health Authorities. This work was performed on the Tjeneste for Sensitive Data (TSD) facilities, owned by the University of Oslo, operated and developed by the TSD service group at the University of Oslo, IT-Department (USIT), using resources provided by Sigma2—the National Infrastructure for High Performance Computing and Data Storage in Norway (UNINETT).

**Ethics Statement**

The current study is based on version 11 of the quality-assured data files released for research in 2019. The establishment of MoBa and initial data collection was based on a license from the Norwegian Data Protection Agency and approval from The Regional Committees for Medical and Health Research Ethics. The MoBa cohort is currently regulated by the Norwegian Health Registry Act. The current study was approved by The Regional Committees for Medical and Health Research Ethics (14140 and 2016/1226). In accordance with REK regulations, individuals who withdraw consent are excluded.

**Disclaimer**

Data from the Norwegian Patient Registry has been used in this publication. The interpretation and reporting of these data are the sole responsibility of the authors, and no endorsement by the Norwegian Patient Registry is intended nor should be inferred.

#### UK Biobank

**PI**

Stephan Ripke

**PMID**

30305743

**Analysis Code**

UKbb

**Sample Description**

The UK Biobank is a large database for biomedical research that includes genomic, lifestyle, environmental and medical data. The UK Biobank database ~500,000 individuals aged between 40–69 years, which were recruited from the UK population between 2006–2010. Cases were identified using ICD-10 summary codes F60.3 (data field: 41270). Only European individuals were retained for the analysis. Cases were matched to controls by age, sex, and the first four principal components (PCs) in a 1:10 ratio resulting in a final sample set of 182 cases and 1820 controls.

**Case Ascertainment**

Hospital inpatient records (ICD-10 summary diagnoses [data field: 41270]: F60.3).

**Control Ascertainment**

Hospital inpatient records (ICD-10 summary diagnoses [data field: 41270]: no history of F60.3).

**Genotyping, Quality Control, Imputation, and Analysis**

UK Biobank participants were assayed on the UK BiLEVE Axiom Array and UK Biobank Axiom Array by Affymetrix. Basic QC was conducted, followed by a genotype imputation to Haplotype Reference Consortium (HRC) and UK10K + 1000 Genomes reference panels. Case-control Genome-Wide Association (GWAS) study was then conducted using general linear regression models implemented in PLINK 2.0 (Chang et al. 2015) on SNPs with a MAF > 1% and INFO scores > 0.6 and array version and PC 1-20 as covariates.

**Funding**

Core funding was obtained from the Wellcome Trust medical charity, Medical Research Council, Department of Health, Scottish Government and the Northwest Regional Development Agency. A full list of funders can be retrieved from: https://www.ukbiobank.ac.uk/learn-more-about-uk-biobank/about-us/our-funding.

**Ethics Statement**

Ethical approval was granted by the North West Multi-centre Research Ethics Committee (MREC). A detailed ethics statement has been released by the UK Biobank under the following link: https://www.ukbiobank.ac.uk/learn-more-about-uk-biobank/about-us/ethics.

PheWAS analysis in the UK Biobank

The PheWAS analysis in the UK Biobank was performed under application number 96802.

#### Mayo Clinic Biobank

**PI**
Brandon Coombes; Joanna Biernacka

**PMID**
31699749

**Analysis Code**
MCbb

**Sample Description**
MCB active enrollment spanned from April 2009 to March 2016 and totaled nearly 60,000 participants aged 18 years and older, selected largely through medical visits to primary care departments at the clinic. Cases were identified using ICD-9/10 diagnosis codes.

**Case Ascertainment**
One or more instances of ICD9: 301.83* and ICD10: F60.3* in the Mayo Clinic electronic health record.

**Control Ascertainment**
There was no screening of controls and thus anyone without a diagnosis of BPD was used as a control.

**Genotyping, Quality Control, Imputation, and Analysis**
Cases and controls were ascertained using EHR data from 57,001 patients from the Mayo Clinic Biobank. Samples were sequenced at the Regeneron Genetics Center (RGC) using a custom design that additionally augments the exome capture with “backbone” regions intended to measure common tagging variation for purposes of GWAS. These backbone regions are targeted at lower depth and undergo substantial post-processing that can boost genotyping quality based on shared information via linkage disequilibrium and population allele frequencies. This is referred to by RGC as genotyping-by-sequencing (GxS).

The resulting GxS data was run through the Mayo Clinic Genotype QC pipeline. In this QC pipeline, SNPs were excluded using filters for call rate (<95%), minor allele frequency (<0.5%), and Hardy-Weinberg Equilibrium ($p < 1e\text{-}6$). Individuals were excluded for excessive missing data (>5%), sex errors, abnormal heterozygosity (< 70% on multiple chromosomes). Analysis of genetic ancestry was performed on a subset of 4874 high-quality HapMap3 SNPs. Principal components analysis (PCA) was first performed on the HapMap3 samples and then MCB samples were projected onto these PCs. Kernel density estimators were trained for each of the individual HapMap3 populations and mapped to one of five main ancestral super-populations (AFR=African; AMR=Admixed American; EAS=East Asian; EUR=European; SAS=South Asian) based on specific likelihood criteria for the individual populations such that the likelihood for a given ancestry group was greater than 0.3, the sample was assigned to that ancestry group. When two ancestry groups had a likelihood of greater than 0.3, RGC assigned AFR over EUR, AMR over EUR, AMR over EAS, SAS over EUR, and AMR over AFR.

Cryptic relatedness analysis was performed in an iterative process using PLINK and PRIMUS to estimate IBD sharing. Highly related samples were removed from the sample if they had >100 closely related samples (PI_HAT>0.1875) or >25000 related samples (PI_HAT>0.08); the relatedness analysis was performed iteratively until no such samples remained. For each pair with an estimated 2nd degree or higher relatedness, we removed the individual with a shorter length of EHR record.

Genotype imputation using the TOPMed Imputation Server was performed after the initial quality control and converted to best-guess genotypes for all markers with high-quality imputation (dosage-R 2 > 0.8).

The GWAS was run using PLINK2 using Firth Regression since the case-control ratio is very unbalanced. The analysis was adjusted for sex and the first principal component of ancestry.

**Funding**
This study was supported by the National Institute of Mental Health (NIMH) grant R01MH121924.

**Ethics Statement**
The study was reviewed and approved by Mayo Clinic's Institutional Review Board (Mayo Clinic IRB approval 19-006227).

#### GLAD

**PI**

Gerome Breen & Thalia Eley

**PMID**

31715324; 37495971

**Analysis Code**

GLAD

**Sample Description**

The Genetic Links to Anxiety and Depression (GLAD) sample is part of the National Institute for Health and Care Research (NIHR) BioResource. The NIHR BioResource is a databank and recontactable resource of volunteers who have provided medical, clinical, and biological data. Volunteers were recruited through a variety of approaches, including National Health Service (NHS) blood transfusion services and various disease/disorder focussed research efforts.

Throughout the pandemic, NIHR BioResource participants were given the opportunity to join the COPING study, which launched in April 2020. The COPING study contained questionnaires from the sign-up surveys of the Genetic Links to Anxiety and Depression (GLAD) Study (Davies et al. 2019) and the Eating Disorders Genetics Initiative (EDGI UK), as well as additional questionnaires to assess COVID-related variables, that is, experiences related to the COVID-19 pandemic. Participants first completed a baseline survey and then follow-up surveys, initially every two weeks but then monthly from August 2020. The first COPING study baseline survey was distributed to existing NIHR BioResource participants on the 30th April 2020, at which point stay-at-home orders had been in place for approximately one month (i.e., since 26th March 2020). All invites for existing participants were sent by 11th May 2020. New GLAD Study and EDGI UK survey participants were sent invitations throughout the pandemic. The last round of invites was sent on 19th January 2021. Details are described elsewhere (Davies et al. 2022). Our study includes all COPING study data collected between April 2020 and July 2021.

COPING study participants come from multiple sub-cohorts of the NIHR BioResource, including: the GLAD Study; EDGI UK; the Inflammatory Bowel Disease BioResource; NHS blood and transplant studies, including INTERVAL , COMPARE, and STRategies to Improve Donor Experiences; and the Research Tissue Bank-Generic. Saliva samples were provided at home and sent in for genotyping.

**Case Ascertainment**

The sign-up process for enrolment in GLAD consisted of collecting personal information and phenotypic data entirely online through the GLAD Study website (www.gladstudy.org.uk). Participants register on the website with their name, email address, phone number, date of birth, sex, and gender. They are then able to read the information sheet and provide consent. As part of the consenting process participants agree to long-term storage of their sample, requests to complete follow-up questionnaires, anonymised data sharing, recontact for future research studies based on their phenotype/genotype information, and access to their full medical and health related records. Following consent, participants complete the sign-up questionnaire to assess their eligibility. To facilitate future meta-analyses with other cohorts, measures were selected when possible to align with the UK Biobank Mental Health Questionnaire (MHQ) (Davis et al. 2020).

The participants were asked the following question: Have you ever been diagnosed with one or more of the following disorders by a professional, even if you don't have it currently? By professional we mean: any doctor, nurse or person with specialist training. Please include disorders even if you did not need treatment for them or if you did not agree with the diagnosis.

This included the following psychiatric disorders: Major depressive disorder, perinatal depression, premenstrual dysphoric disorder, bipolar disorder, generalised anxiety disorder, social anxiety, specific phobia, agoraphobia, panic disorder, panic attacks, posttraumatic stress disorder, obsessive-compulsive disorder (OCD), body dysmorphic disorder, other ocd-like syndromes, anorexia nervosa, bulimia nervosa, binge-eating disorder, atypical anorexia nervosa, schizophrenia, schizoaffective disorder, psychosis, personality disorder, autism spectrum disorder, attention deficit hyperactivity disorder.

**Control Ascertainment**

Study participants who did not endorse any of these disorders: Major depressive disorder, perinatal depression, premenstrual dysphoric disorder, bipolar disorder, generalised anxiety disorder, social anxiety, specific phobia, agoraphobia, panic disorder, panic attacks, posttraumatic stress disorder, obsessive-compulsive disorder (OCD), body dysmorphic disorder, other ocd-like syndromes, anorexia nervosa, bulimia nervosa, binge-eating disorder, atypical anorexia nervosa, schizophrenia, schizoaffective disorder, psychosis, personality disorder, autism spectrum disorder, attention deficit hyperactivity disorder.

**Genotyping, Quality Control, Imputation, and Analysis**

All data from GLAD and COPING NBR were genotyped by ThermoFisher on the UK Biobank Axiom Array v1 and v2 across numerous genotyping batches. Genetic data for GLAD and COPING NBR cohorts were merged, and restricted to individuals from European ancestries (749,044 SNPs before quality control). Ancestry was determined using GenoPred (https://opain.github.io/GenoPred/index.html), by projecting GLAD and COPING individuals on genomic principal components from the 1000 Genomes reference data, and assigning individuals a genetic ancestry if they lay < 3 SD from the mean of individuals from that ancestry superpopulation in 1000 Genomes. Quality control was conducted, excluding variants with MAF < 0.01, call rate < 0.95, or which were deviant from Hardy-Weinberg equilibrium (p < 10^-10).

Individuals were excluded if they had withdrawn from the study following genotyping, if they were a duplicate of a higher-quality sample (not including known identical twins), if they were known to be mislabelled, if their genotypic sex (males Fx > 0.8, females Fx < 0.5) did not match their sex assigned at birth, if they were outliers on genome-wide heterozygosity (absolute(Fhat) > 0.2), or if they had an excess of relatives (average pi-hat > 3 SD from the mean). Following quality control, 33,635 individuals and 484,182 variants were available for imputation. Imputation was carried out to TopMED Freeze 8, using the dedicated imputation server (https://imputation.biodatacatalyst.nhlbi.nih.gov/#!). Following imputation, data was further restricted to data with MAF >= 0.01 and R2 >= 0.3.

**Funding**

This work was supported by the National Institute of Health Research (NIHR) BioResource, NIHR Biomedical Research Centre [IS-BRC-1215-20018], HSC R&D Division, Public Health Agency [COM/5516/18], MRC Mental Health Data Pathfinder Award (MC_PC_17,217), and the National Centre for Mental Health funding through Health and Care Research Wales.

**Ethics Statement**

The London-Fulham Research Ethics Committee approved the GLAD Study on 21st August 2018 (REC reference: 18/LO/1218) and EDGI UK on 29th July 2019 (REC reference: 19/LO/1254). The NIHR BioResource has been approved as a Research Tissue Bank by the East of England-Cambridge Central Committee (REC reference: 17/EE/0025). The COVID-19 Psychiatry and Neurological Genetics study was approved by the South West-Central Bristol Research Ethics Committee on 27th April 2020 (REC reference: 20/SW/0078). The RAMP Study was approved by the Psychiatry, Nursing, and Midwifery Research Ethics Committee at King's College London on 27th March 2020 (HR-19/20–18157).

#### FinnGen

**PI**

Jaakko Kaprio

**PMID**

36829046

**Analysis Code**

FinGen

**Sample Description**

FinnGen is a large public-private partnership aiming to collect and analyse genome and health data from 500,000 Finnish biobank participants. FinnGen aims on one hand to provide novel medically and therapeutically relevant insights but also construct a world-class resource that can be applied for future studies. The project aims to produce close to complete genome variant data using GWAS genotyping.

The GWAS data are combined with phenotype data produced from several national health registries, such as hospital discharge records, prescription records and cause-of death data.. The collected samples consist of two entities: 1) legacy samples from multiple existing cohort studies, most of which collected by the THL (National Institute for Health and Welfare) and 2) about 300 000 prospective samples which have mainly been collected by hospital biobanks.

**Case Ascertainment**

Hospital discharge registry: ICD-10: F60.3, ICD-9: 3018D, ICD-8: 3013.

**Control Ascertainment**

Hospital discharge registry: excluded any of the following diagnoses: Disorders of adult personality and behaviour (ICD-10: F60, F62, F63, F64, F65, F66, F68, F69 & ICD-9: 312, 3023, 3025, 3026, 3021, 3022, 3024, 3028, 3029, 3020 & ICD-8: 302, 3023)

**Genotyping, Quality Control, Imputation, and Analysis**

Chip genotype data processing and QC Samples were genotyped with Illumina (Illumina Inc., San Diego, CA, USA) and Affymetrix arrays (Thermo Fisher Scientific, Santa Clara, CA, USA). Genotype calls were made with GenCall and zCall algorithms for Illumina and AxiomGT1 algorithm for Affymetrix data.

Chip genotyping data produced with previous chip platforms and reference genome builds were lifted over to build version 38 (GRCh38/hg38) following the protocol described here: dx.doi.org/10.17504/protocols.io.xbhfij6. In sample-wise quality control steps, individuals with ambiguous gender, high genotype missingness (>5%), excess heterozygosity (+-4SD) and non-Finnish ancestry were excluded. In variant-wise quality control steps, variants with high missingness (>2%), low HWE P-value (<1e-6) and low minor allele count (MAC<3) were excluded. Before imputation, chip-genotyped samples were pre-phased with Eagle 2.3.5 using the default parameters, except the number of conditioning haplotypes, which was set to 20,000. Genotype imputation was carried out by using the population-specific SISu v4.0 imputation reference panel with Beagle 4.1 (version 08Jun17.d8b) as described in the following protocol: dx.doi.org/10.17504/protocols.io.xbgfijw. Genome-wide association analyses were run using Firth-logistic regression on PLINK 2.0 on SNPs with imputation quality >0.6 and MAC>5, having age at end of assessment or death, gender and 10 PCs as covariates.

**Funding**

The FinnGen project is funded by two grants from Business Finland (HUS 4685/31/2016 and UH 4386/31/2016) and the following industry partners: AbbVie Inc., AstraZeneca UK Ltd, Biogen MA Inc., Bristol Myers Squibb (and Celgene Corporation & Celgene International II Sàrl), Genentech Inc., Merck Sharp & Dohme LCC, Pfizer Inc., GlaxoSmithKline Intellectual Property Development Ltd., Sanofi US Services Inc., Maze Therapeutics Inc., Janssen Biotech Inc, Novartis Pharma AG, and Boehringer Ingelheim International GmbH.

**Ethics Statement**

Patients and control subjects in FinnGen provided informed consent for biobank research, based on the Finnish Biobank Act. Alternatively, separate research cohorts, collected prior the Finnish Biobank Act came into effect (in September 2013) and start of FinnGen (August 2017), were collected based on study-specific consents and later transferred to the Finnish biobanks after approval by Fimea (Finnish Medicines Agency), the National Supervisory Authority for Welfare and Health. Recruitment protocols followed the biobank protocols approved by Fimea. The Coordinating Ethics Committee of the Hospital District of Helsinki and Uusimaa (HUS) statement number for the FinnGen study is Nr HUS/990/2017.

The FinnGen study is approved by Finnish Institute for Health and Welfare (permit numbers: THL/2031/6.02.00/2017, THL/1101/5.05.00/2017, THL/341/6.02.00/2018, THL/2222/6.02.00/2018, THL/283/6.02.00/2019, THL/1721/5.05.00/2019 and THL/1524/5.05.00/2020), Digital and population data service agency (permit numbers: VRK43431/2017-3, VRK/6909/2018-3, VRK/4415/2019-3), the Social Insurance Institution (permit numbers: KELA 58/522/2017, KELA 131/522/2018, KELA 70/522/2019, KELA 98/522/2019, KELA 134/522/2019, KELA 138/522/2019, KELA 2/522/2020, KELA 16/522/2020), Findata permit numbers THL/2364/14.02/2020, THL/4055/14.06.00/2020,,THL/3433/14.06.00/2020, THL/4432/14.06/2020, THL/5189/14.06/2020, THL/5894/14.06.00/2020, THL/6619/14.06.00/2020, THL/209/14.06.00/2021, THL/688/14.06.00/2021, THL/1284/14.06.00/2021, THL/1965/14.06.00/2021, THL/5546/14.02.00/2020, THL/2658/14.06.00/2021, THL/4235/14.06.00/2021 and Statistics Finland (permit numbers: TK-53-1041-17 and TK/143/07.03.00/2020 (earlier TK-53-90-20) TK/1735/07.03.00/2021).

The Biobank Access Decisions for FinnGen samples and data utilized in FinnGen Data Freeze 8 include: THL Biobank BB2017_55, BB2017_111, BB2018_19, BB_2018_34, BB_2018_67, BB2018_71, BB2019_7, BB2019_8, BB2019_26, BB2020_1, Finnish Red Cross Blood Service Biobank 7.12.2017, Helsinki Biobank HUS/359/2017, Auria Biobank AB17-5154 and amendment #1 (August 17 2020), AB20-5926 and amendment #1 (April 23 2020), Biobank Borealis of Northern Finland_2017_1013, Biobank of Eastern Finland 1186/2018 and amendment 22 § /2020, Finnish Clinical Biobank Tampere MH0004 and amendments (21.02.2020 & 06.10.2020), Central Finland Biobank 1-2017, and Terveystalo Biobank STB 2018001.

### Description of replication studies

#### IPM-Reus (Spain 2 replication)

**PI**
Lourdes Martorell and Elisabet Vilella

**PMID**
35396580

**Analysis Code**
borls.rep

**Sample Description**
This sample consisted of both cases and controls who participated in research studies on the genetics of psychiatric disorders and provided a blood sample for this purpose.
Patients were selected from admissions to the Hospital Universitari Institut Pere Mata (Spain). Inclusion criteria for patients were an age between 18 and 65 years and a ICD-9 diagnosis code of borderline personality disorder. Exclusion criteria were non-Caucausian origin and intellectual disability.
Controls were drawn from two population-based samples from the same geographic region as the cases. Inclusion criteria were age between 18 and 75 years old. Exclusion criteria were: personal history of psychiatric disorder and non-Caucasian origin.

**Case Ascertainment**
In the clinical health record, one or more instances of ICD-9: 301.83.

**Control Ascertainment**
Individuals from the general population were interviewed by a trained researcher to rule out a personal history of psychiatric disorder.

**Genotyping**
Total DNA was extracted from peripheral blood samples using Gentra Puregene® reagents (Qiagen, Germany) according to established procedures. Genotyping was performed using the Infinium PsychArray-24 Bead Chip (Illumina, San Diego, CA, USA).

**Funding**
This work was supported by the Agència de Gestió d'Ajuts Universitaris i de Recerca (AGAUR, 2021SGR-001065)

**Ethics Statement**
Participants (cases and the two population-based samples) were included in three different research projects approved by the Ethics Committee of the Hospital Universitari Sant Joan de Reus (06/2000 for case samples; 06/1998 and 10/2000 for control samples). All participants provided written consent before participating in the study.

#### All of Us

**PI**

The All of Us Research Program Investigators

**PMID**

31412182; 38374255

**Analysis Code**

AllOfUs

**Sample Description**

The All of Us Research Program aims to enroll one million individuals across the United States, with a focus on fostering diversity to support progress in biomedical research and enhance public health. Enrollment began in May 2018 and is open to adults aged 18 and older through a network of over 340 recruitment centers. The program gathers information through health surveys, electronic health records (EHRs), physical assessments, digital health tools, and the collection of biological samples.

As of June 2025, more than 862,000 participants have enrolled in All of Us. Additionally, EHR data from over 470,000 individuals have been compiled. We used data release version 7 of the All of Us cohort's short-read whole-genome sequence data to conduct replication and polygenic scoring analyses. The data were accessed via the Controlled Tier of the All of Us Researcher Workbench.

https://support.researchallofus.org/hc/en-us/articles/14769699298324-Curated-Data-Repository-CDR-version-7-Release-Notes

**Case Ascertainment**

Borderline personality disorder cases were defined using phecode MB_296.4, aggregated via PhecodeX (https://github.com/PheWAS/PhecodeX), and mapped to the ICD-10 code F60.3. We included cases that were enrolled in the All of Us study, had samples subjected to short-read whole-genome sequencing, and were included in the All of Us Controlled Tier Dataset v7.

**Control Ascertainment**

We included unscreened controls who were enrolled in the All of Us study, had samples subjected to short-read whole-genome sequencing, and were included in the All of Us Controlled Tier Dataset v7.

**Genotyping, Quality Control, Imputation, and Analysis**

A detailed report on sample handling, processing, and quality control is available on the All of Us Researcher Hub website:

https://support.researchallofus.org/hc/en-us/articles/29390274413716-All-of-Us-Genomic-Quality-Report For the presented analyses, we included individuals of European ancestry, as the sample sizes for other ancestry groups were too small for robust analysis. Ancestry was predicted as described in the All of Us Research Program Investigators' 2024 publication (https://doi.org/10.1038/s41586-023-06957-x). We excluded any related or flagged samples, as indicated in the Auxiliary Data available in the All of Us Workbench. We filtered down the "ACAF" callset (population-specific allele count > 100 in any computed ancestry subpopulation or population-specific allele frequency > 1%) derived from the short-read whole-genome sequencing data to retain only variants present in the PRS-CS output file. Polygenic scoring was performed using PLINK 1.9, with PRS-CS-derived posterior SNP effect sizes, and the first 10 principal components included in the logistic regression model to control for population stratification. Output measures were calculated based on the following analysis script: https://github.com/Ripkelab/ricopili/blob/master/rp_bin/danscore_result_3

**Funding**

All of Us is supported by grants through the National Institutes of Health Office of the Director: Regional Medical Centers: 1 OT2 OD026549; 1 OT2 OD026554; 1 OT2 OD026557; 1 OT2 OD026556; 1 OT2 OD026550; 1 OT2 OD 026552; 1 OT2 OD026553; 1 OT2 OD026548; 1 OT2 OD026551; 1 OT2 OD026555; IAA #: AOD 16037; Federally Qualified Health Centers: HHSN 263201600085U; Data and Research Center: 5 U2C OD023196; Biobank: 1 U24 OD023121; The Participant Center: U24 OD023176; Participant Technology Systems Center: 1 U24 OD023163; Communications and Engagement: 3 OT2 OD023205; 3 OT2 OD023206; Community Partners: 1 OT2 OD025277; 3 OT2 OD025315; 1 OT2 OD025337; 1 OT2 OD025276; and the All of Us Pilot: 1 OT2 OD023132.

**Ethics Statement**

Analyses were conducted in accordance with the All of Us Code of Conduct

**Acknowledgment**

The All of Us Research Program is supported by the National Institutes of Health, Office of the Director: Regional Medical Centers: 1 OT2 OD026549; 1 OT2 OD026554; 1 OT2 OD026557; 1 OT2 OD026556; 1 OT2 OD026550; 1 OT2 OD 026552; 1 OT2 OD026553; 1 OT2 OD026548; 1 OT2 OD026551; 1 OT2 OD026555; IAA #: AOD 16037; Federally Qualified Health Centers: HHSN 263201600085U; Data and Research Center: 5 U2C OD023196; Biobank: 1 U24 OD023121; The Participant Center: U24 OD023176; Participant Technology Systems Center: 1 U24 OD023163; Communications and Engagement: 3 OT2 OD023205; 3 OT2 OD023206; and Community Partners: 1 OT2 OD025277; 3 OT2 OD025315; 1 OT2 OD025337; 1 OT2 OD025276. In addition, the All of Us Research Program would not be possible without the partnership of its participants.

Chang, Christopher C., Carson C. Chow, Laurent Cam Tellier, Shashaank Vattikuti, Shaun M. Purcell, and James J. Lee. 2015. "Second-Generation PLINK: Rising to the Challenge of Larger and Richer Datasets." *GigaScience* 4 (February):7.

Corfield, Elizabeth C., Oleksandr Frei, Alexey A. Shadrin, Zillur Rahman, Aihua Lin, Lavinia Athanasiu, Bayram Cevdet Akdeniz, et al. 2022. "The Norwegian Mother, Father, and Child Cohort Study (MoBa) Genotyping Data Resource: MoBaPsychGen Pipeline v.1." *bioRxiv*. https://doi.org/10.1101/2022.06.23.496289.

Das, Sayantan, Lukas Forer, Sebastian Schönherr, Carlo Sidore, Adam E. Locke, Alan Kwong, Scott I. Vrieze, et al. 2016. "Next-Generation Genotype Imputation Service and Methods." *Nature Genetics* 48 (10): 1284–87.

Davies, Molly R., Joshua E. J. Buckman, Brett N. Adey, Chérie Armour, John R. Bradley, Susannah C. B. Curzons, Helena L. Davies, et al. 2022. "Comparison of Symptom-Based versus Self-Reported Diagnostic Measures of Anxiety and Depression Disorders in the GLAD and COPING Cohorts." *Journal of Anxiety Disorders* 85 (January):102491.

Davies, Molly R., Gursharan Kalsi, Chérie Armour, Ian R. Jones, Andrew M. McIntosh, Daniel J. Smith, James T. R. Walters, et al. 2019. "The Genetic Links to Anxiety and Depression (GLAD) Study: Online Recruitment into the Largest Recontactable Study of Depression and Anxiety." *Behaviour Research and Therapy* 123 (December):103503.

Davis, Katrina A. S., Jonathan R. I. Coleman, Mark Adams, Naomi Allen, Gerome Breen, Breda Cullen, Chris Dickens, et al. 2020. "Mental Health in UK Biobank--Development, Implementation and Results from an Online Questionnaire Completed by 157 366 Participants: A Reanalysis." *BJPsych Open* 6 (2): e18.

Dennis, Jessica, Julia Sealock, Rebecca T. Levinson, Eric Farber-Eger, Jacob Franco, Sarah Fong, Peter Straub, et al. 2021. "Genetic Risk for Major Depressive Disorder and Loneliness in Sex-Specific Associations with Coronary Artery Disease." *Molecular Psychiatry* 26 (8): 4254–64.

Eggertsson, Hannes P., Hakon Jonsson, Snaedis Kristmundsdottir, Eirikur Hjartarson, Birte Kehr, Gisli Masson, Florian Zink, et al. 2017. "Graphtyper Enables Population-Scale Genotyping Using Pangenome Graphs." *Nature Genetics* 49 (11): 1654–60.

Fagerstrom, K. O., and N. G. Schneider. 1989. "Measuring Nicotine Dependence: A Review of the Fagerstrom Tolerance Questionnaire." *Journal of Behavioral Medicine* 12 (2):

159–82.

First, Michael B., Robert L. Spitzer, Miriam Gibbon, and Janet B. W. Williams. 1995. “The Structured Clinical Interview for DSM-III-R Personality Disorders (SCID-II). Part I: Description.” *Journal of Personality Disorders* 9 (2): 83–91.

Franke, Gabriele Helga, and Leonard R. Derogatis. 2002. *Symptom-Checkliste von LR Derogatis: SCL-90-R; Deutsche Version*. Beltz Test.

Geissbühler, Antoine, Dario Giuse, Jonathan Grande, Randolph A. Miller, and William W. Stead. 1998. “The Vanderbilt Patient-Care Information System.” *Proceedings: A Conference of the American Medical Informatics Association / ... AMIA Annual Fall Symposium. AMIA Fall Symposium*, 1121.

Gudbjartsson, Daniel F., Hannes Helgason, Sigurjon A. Gudjonsson, Florian Zink, Asmundur Oddson, Arnaldur Gylfason, Soren Besenbacher, et al. 2015. “Large-Scale Whole-Genome Sequencing of the Icelandic Population.” *Nature Genetics* 47 (5): 435–44.

Gudbjartsson, Daniel F., Patrick Sulem, Hannes Helgason, Arnaldur Gylfason, Sigurjon A. Gudjonsson, Florian Zink, Asmundur Oddson, et al. 2015. “Sequence Variants from Whole Genome Sequencing a Large Group of Icelanders.” *Scientific Data* 2 (March):150011.

Guo, Yan, Jing He, Shilin Zhao, Hui Wu, Xue Zhong, Quanhu Sheng, David C. Samuels, Yu Shyr, and Jirong Long. 2014. “Illumina Human Exome Genotyping Array Clustering and Quality Control.” *Nature Protocols* 9 (11): 2643–62.

James Kent, W. 2002. “BLAT—The BLAST-Like Alignment Tool.” *Genome Research* 12 (4): 656–64.

Kong, Augustine, Gisli Masson, Michael L. Frigge, Arnaldur Gylfason, Pasha Zusmanovich, Gudmar Thorleifsson, Pall I. Olason, et al. 2008. “Detection of Sharing by Descent, Long-Range Phasing and Haplotype Imputation.” *Nature Genetics* 40 (9): 1068–75.

Lam, Max, Swapnil Awasthi, Hunna J. Watson, Jackie Goldstein, Georgia Panagiotaropoulou, Vassily Trubetskoy, Robert Karlsson, et al. 2019. “RICOPILI: Rapid Imputation for COnsortias PIpeLIne.” *Bioinformatics* , August. https://doi.org/10.1093/bioinformatics/btz633.

Langbehn, D. R., B. M. Pfohl, S. Reynolds, L. A. Clark, M. Battaglia, L. Bellodi, R. Cadoret, W. Grove, P. Pilkonis, and P. Links. 1999. “The Iowa Personality Disorder Screen: Development and Preliminary Validation of a Brief Screening Interview.” *Journal of Personality Disorders* 13 (1): 75–89.

Leichsenring, Falk, Nikolas Heim, Frank Leweke, Carsten Spitzer, Christiane Steinert, and Otto F. Kernberg. 2023. “Borderline Personality Disorder: A Review.” *JAMA: The Journal of the American Medical Association* 329 (8): 670–79.

Li, Jun Z., Devin M. Absher, Hua Tang, Audrey M. Southwick, Amanda M. Casto, Sohini Ramachandran, Howard M. Cann, et al. 2008. “Worldwide Human Relationships Inferred from Genome-Wide Patterns of Variation.” *Science* 319 (5866): 1100–1104.

Loh, Po-Ru, Petr Danecek, Pier Francesco Palamara, Christian Fuchsberger, Yakir A Reshef, Hilary K Finucane, Sebastian Schoenherr, et al. 2016. “Reference-Based Phasing Using the Haplotype Reference Consortium Panel.” *Nature Genetics* 48 (11): 1443–48.

Loh, Po-Ru, Pier Francesco Palamara, and Alkes L. Price. 2016. “Fast and Accurate Long-Range Phasing in a UK Biobank Cohort.” *Nature Genetics* 48 (7): 811–16.

Magnus, Per, Charlotte Birke, Kristine Vejrup, Anita Haugan, Elin Alsaker, Anne Kjersti Daltveit, Marte Handal, et al. 2016. “Cohort Profile Update: The Norwegian Mother and Child Cohort Study (MoBa).” *International Journal of Epidemiology* 45 (2): 382–88.

Marchini, Jonathan, Bryan Howie, Simon Myers, Gil McVean, and Peter Donnelly. 2007. “A New Multipoint Method for Genome-Wide Association Studies by Imputation of Genotypes.” *Nature Genetics* 39 (7): 906–13.

McGirr, Alexander, Joel Paris, Alain Lesage, Johanne Renaud, and Gustavo Turecki. 2007. “Risk Factors for Suicide Completion in Borderline Personality Disorder: A Case-Control Study of Cluster B Comorbidity and Impulsive Aggression.” *The Journal of Clinical*

*Psychiatry* 68 (5): 721–29.
Mitt, Mario, Mart Kals, Kalle Pärn, Stacey B. Gabriel, Eric S. Lander, Aarno Palotie, Samuli Ripatti, et al. 2017. "Improved Imputation Accuracy of Rare and Low-Frequency Variants Using Population-Specific High-Coverage WGS-Based Imputation Reference Panel." *European Journal of Human Genetics: EJHG* 25 (7): 869–76.
Olssøn, Ingrid, Øystein Sørebø, and Alv A. Dahl. 2011. "A Cross-Sectional Testing of The Iowa Personality Disorder Screen in a Psychiatric Outpatient Setting." *BMC Psychiatry* 11 (June):105.
Paltiel, Liv, Haugan Anita, Trine Skjerden, Kari Harbak, Siri Bækken, Stensrud Nina Kristin, Gun Peggy Knudsen, and Per Magnus. 2014. "The Biobank of the Norwegian Mother and Child Cohort Study – Present Status." *Norsk Epidemiologi* 24 (1-2). https://doi.org/10.5324/nje.v24i1-2.1755.
Pedersen, Geir, Theresa Wilberg, Benjamin Hummelen, and Elfrida Hartveit Kvarstein. 2023. "The Norwegian Network for Personality Disorders – Development, Contributions and Challenges through 30 Years." *Nordic Journal of Psychiatry* 77 (5): 512–20.
Roden, D. M., J. M. Pulley, M. A. Basford, G. R. Bernard, E. W. Clayton, J. R. Balser, and D. R. Masys. 2008. "Development of a Large-Scale de-Identified DNA Biobank to Enable Personalized Medicine." *Clinical Pharmacology and Therapeutics* 84 (3): 362–69.
Saunders, J. B., O. G. Aasland, T. F. Babor, J. R. de la Fuente, and M. Grant. 1993. "Development of the Alcohol Use Disorders Identification Test (AUDIT): WHO Collaborative Project on Early Detection of Persons with Harmful Alcohol Consumption--II." *Addiction* 88 (6): 791–804.
Schmahl, Christian, Sabine C. Herpertz, Katja Bertsch, Gabriele Ende, Herta Flor, Peter Kirsch, Stefanie Lis, et al. 2014. "Mechanisms of Disturbed Emotion Processing and Social Interaction in Borderline Personality Disorder: State of Knowledge and Research Agenda of the German Clinical Research Unit." *Borderline Personality Disorder and Emotion Dysregulation* 1 (September):12.
Sheehan, D., J. Janavs, R. Baker, K. Harnett-Sheehan, E. Knapp, and M. Sheehan. 1994. "Mini International Neuropsychiatric Interview." *Tampa: University of South Florida*.
Stangl, D., B. Pfohl, M. Zimmerman, W. Bowers, and C. Corenthal. 1985. "A Structured Interview for the DSM-III Personality Disorders. A Preliminary Report." *Archives of General Psychiatry* 42 (6): 591–96.
Wang, Chaolong, Xiaowei Zhan, Jennifer Bragg-Gresham, Hyun Min Kang, Dwight Stambolian, Emily Y. Chew, Kari E. Branham, et al. 2014. "Ancestry Estimation and Control of Population Stratification for Sequence-Based Association Studies." *Nature Genetics* 46 (4): 409–15.
Witt, S. H., F. Streit, M. Jungkunz, J. Frank, S. Awasthi, C. S. Reinbold, J. Treutlein, et al. 2017. "Genome-Wide Association Study of Borderline Personality Disorder Reveals Genetic Overlap with Bipolar Disorder, Major Depression and Schizophrenia." *Translational Psychiatry* 7 (6): e1155.
Wolf, Martina, Matthias F. Limberger, Nikolaus Kleindienst, Rolf-Dieter Stieglitz, Melanie Domsalla, Alexandra Philipsen, Regina Steil, and Martin Bohus. 2009. "Kurzversion der Borderline-Symptom-Liste (BSL-23): Entwicklung und Überprüfung der psychometrischen Eigenschaften." *Psychotherapie, Psychosomatik, medizinische Psychologie* 59 (8): 321–24.
Zhou, Wei, Jonas B. Nielsen, Lars G. Fritsche, Rounak Dey, Maiken E. Gabrielsen, Brooke N. Wolford, Jonathon LeFaive, et al. 2018. "Efficiently Controlling for Case-Control Imbalance and Sample Relatedness in Large-Scale Genetic Association Studies." *Nature Genetics* 50 (9): 1335–41.

### Members of contributing groups

#### HUNT All-In Psychiatry

Anne Heidi Skogholt PhD[1], Børge Sivertsen PhD[2,3,4], Eystein Stordal MD, PhD[3,5], Grete Dyb MD, PhD[6], Gunnar Morken MD, PhD[3,7], Håvard Kallestad MD[3,7], Ingrid Heuch MD, PhD[8], Jonas B Nielsen PhD[1,9], Katrine K Fjukstad MD, PhD[10,11], Lars G Fritsche PhD[12], Laurent F Thomas PhD[1,13,14,15], Linda M Pedersen PhD[8], Maiken E Gabrielsen PhD[1], Marit S Indredavik MD, PhD[16], Marit Skrove MD, PhD[17], Ole A Andreassen MD, PhD[18], Ottar Bjerkeset MD, PhD[3,19], Sigrid Børte MD, PhD[20,1,21], Synne Ø Stensland MD, PhD[21,6], Torunn S Nøvik MD, PhD[22], Wei Zhou PhD[23,24]

1 K. G. Jebsen Center for Genetic Epidemiology, Department of Public Health and Nursing, Faculty of Medicine and Health Sciences, Norwegian University of Science and Technology (NTNU), Trondheim, Norway.
2 Department of Health Promotion, Norwegian Institute of Public Health, Bergen, Norway.
3 Department of Mental Health, Faculty of Medicine and Health Sciences, Norwegian University of Science and Technology (NTNU), Trondheim, Norway.
4 Department of Research and Innovation, Helse-Fonna HF, Haugesund, Norway.
5 Department of Psychiatry, Hospital Namsos, Nord-Trøndelag Health Trust, Namsos, Norway.
6 Norwegian Centre for Violence and Traumatic Stress Studies, Oslo, Norway.
7 Division of Mental Health Care, St. Olavs Hospital, Trondheim University Hospital, Trondheim, Norway.
8 Department of Research and Innovation, Division of Clinical Neuroscience, Oslo University Hospital, Oslo, Norway.
9 Department of Internal Medicine, Division of Cardiovascular Medicine, University of Michigan, Ann Arbor, MI, 48109, USA.
10 Department of Psychiatry, Nord-Trøndelag Hospital Trust, Levanger Hospital, Levanger, Norway.
11 Department of Laboratory Medicine, Children’s and Women’s Health, Norwegian University of Science and Technology (NTNU), Trondheim, Norway.
12 Center for Statistical Genetics, Department of Biostatistics, University of Michigan, Ann Arbor, MI, 48109, USA.
13 Department of Clinical and Molecular Medicine, Norwegian University of Science and Technology (NTNU), Trondheim, Norway.
14 BioCore - Bioinformatics Core Facility, Norwegian University of Science and Technology (NTNU), Trondheim, Norway.
15 Clinic of Laboratory Medicine, St. Olavs Hospital, Trondheim University Hospital, Trondheim, Norway.

16 Department of Clinical and Molecular Medicine, Faculty of Medicine and Health Sciences, Norwegian University of Science and Technology (NTNU), Trondheim, Norway.
17 Regional Centre for Child and Youth Mental Health and Child Welfare, Department of Mental Health, Faculty of Medicine and Health Sciences, Norwegian University of Science and Technology (NTNU), Trondheim, Norway.
18 Division of Mental Health and Addiction, Oslo University Hospital, Oslo, Norway.
19 Faculty of Nursing and Health Sciences, NORD University, Levanger, Norway.
20 Institute of Clinical Medicine, Faculty of Medicine, University of Oslo, Oslo, Norway.
21 Research and Communication Unit for Musculoskeletal Health (FORMI), Department of Research and Innovation, Division of Clinical Neuroscience, Oslo University Hospital, Oslo, Norway.
22 Department of Child and Adolescent Psychiatry, St. Olavs Hospital, Trondheim University Hospital, Trondheim, Norway.
23 Department of Computational Medicine and Bioinformatics, University of Michigan, Ann Arbor, MI, 48109, USA.
24 Analytic and Translational Genetics Unit, Massachusetts General Hospital, Boston, MA, USA.

#### Estonian Biobank Research Team

Andres Metspalu

Lili Milani

Tõnu Esko

Reedik Mägi

Mari Nelis

Georgi Hudjashov

#### The GLAD Study Group Authors

Gursharan Kalsi[1,2], Saakshi Kakar[1,2], Christopher Hübel[1,2,3,4], Ian Marsh[1,2], Laura H Meldrum[1,2], Iona Smith[1,2], Jahnavi Arora[1,2], Henry C. Rogers[1,2,5], Brett N. Adey[1,2], Zain Ahmad[1], Shannon Bristow[1,2], Charles J. Curtis[1,2], Susannah C. B. Curzons[1,2], Helena L. Davies[1,6,7], Molly R. Davies[8], Abigail R. ter Kuile[1,2], Sang Hyuck Lee[1,2], Yuhao Lin[1,2], Jared G. Maina[1,2], Monika McAtarsney-Kovacs[1,2], Dina Monssen[1,2], Jessica Mundy[1,2],

Alish B. Palmos[1,2], Alicia J. Peel[1,2], Kirstin Purves[1,2], Christopher Rayner[1,2], Megan Skelton[1,2], Katherine N. Thompson[1,9], Rujia Wang[1,2], Johan Zvrskovec[1,2], Joshua E. J. Buckman[10], Ewan Carr[11], Antony J. Cleare[12,13], Katrina A. S. Davis[8], Kimberly A. Goldsmith[11], Colette R. Hirsch[14,15], Georgina Krebs[1,16], Donald M. Lyall[17], Katharine A. Rimes[12], Evangelos Vassos[1,2], David Veale[12,13], Janet Wingrove[18], Allan H. Young[19], Roland Zahn[19], Le Roy Dowey[20], Victor Gault[20], Chérie Armour[21], John R. Bradley[22], Ian R. Jones[23], Nathalie Kingston[22], Andrew M. McIntosh[24], Daniel J. Smith[25], James T. R. Walters[26], NIHR BioResource consortium, Jonathan R. I. Coleman[1,2], Matthew Hotopf[2,8], Thalia C. Eley[1,2], Gerome Breen[1,2]

*1. Social, Genetic, and Developmental Psychiatry Centre; Institute of Psychiatry, Psychology and Neuroscience; King's College London, London, UK*

*2. UK National Institute for Health Research (NIHR) Biomedical Research Centre, South London and Maudsley Hospital and King's College London, London, UK*

*3. National Centre for Register-based Research, Aarhus University, Aarhus, Denmark*

*4. Department of Pediatric Neurology, Charité - Universitätsmedizin Berlin, Berlin, Germany*

*5. Department of Psychiatry, Mount Sinai Health System, New York, USA*

*6. Mental Health Center Ballerup, Copenhagen University Hospital – Mental Health Services CPH, Center for Eating and feeding Disorders Research, Copenhagen, Denmark*

*7. Institute of Biological Psychiatry, Mental Health Center Sct. Hans, Mental Health Services Copenhagen, Roskilde, Denmark*

*8. Department of Psychological Medicine, Institute of Psychiatry, Psychology & Neuroscience, King's College London, London, UK*

*9. Department of Sociology, College of Liberal Arts, Purdue University, West Lafayette, IN, USA*

*10. CORE Data Lab, Centre for Outcomes Research and Effectiveness (CORE), Research Department of Clinical, Educational, and Health Psychology, UCL, London, UK*

*11. Department of Biostatistics and Health Informatics, Institute of Psychiatry, Psychology and Neuroscience, King's College London, London, UK*

*12. The Institute of Psychiatry, Psychology and Neuroscience, King's College London, London, UK*
*13. South London and Maudsley NHS Foundation Trust, Maudsley Hospital, London, UK*
*14. Department of Psychology, Institute of Psychiatry, Psychology and Neuroscience, King's College London, Denmark Hill, Camberwell, London, UK*

*15. Centre for Anxiety Disorders and Trauma, South London and Maudsley Hospital, London, UK*
*16. Research Department of Clinical, Educational and Health Psychology, University College London, London, UK*
*17. Institute of Health and Wellbeing, University of Glasgow, Glasgow, UK*

*18. Talking Therapies Southwark, South London and Maudsley NHS Foundation Trust, London, UK*

*19. South London and Maudsley NHS Foundation Trust and Centre for Affective Disorders, Department of Psychological Medicine, Institute of Psychiatry, Psychology, and Neuroscience, King's College London, London, UK*
*20. School of Biomedical Sciences, Ulster University, Coleraine, Northern Ireland, UK*

*21. Research Centre for Stress Trauma and Related Conditions (STARC), School of Psychology, Queen's University Belfast, Belfast, UK*

*22. University of Cambridge, Cambridge, UK*

*23. National Centre for Mental Health, Cardiff University, Cardiff, UK*

*24. Division of Psychiatry, Centre for Clinical Brain Sciences, University of Edinburgh, Edinburgh, UK*

*25. Division of Psychiatry, Centre for Clinical Brain Sciences, University of Edinburgh, Royal Edinburgh Hospital, Edinburgh, UK*
*26. Centre for Neuropsychiatric Genetics and Genomics, Division of Psychological Medicine and Clinical Neurosciences, School of Medicine, Cardiff University, Cardiff, UK*

#### DBDS Genomic Consortium

Karina Banasik; Novo Nordisk Foundation Center for Protein Research, Faculty of Health and Medical Sciences, University of Copenhagen, Copenhagen, Denmark
Jakob Bay; Department of Clinical Immunology, Zealand University Hospital, Køge, Denmark
Jens Kjærgaard Boldsen; Department of Clinical Immunology, Aarhus University Hospital, Aarhus, Denmark
Thorsten Brodersen; Department of Clinical Immunology, Zealand University Hospital, Køge, Denmark
Søren Brunak; Novo Nordisk Foundation Center for Protein Research, Faculty of Health and Medical Sciences, University of Copenhagen, Copenhagen, Denmark
Alfonso Buil Demur; Institute of Biological Psychiatry, Mental Health Centre, Sct. Hans, Copenhagen University Hospital, Roskilde, Denmark
Lea Arregui Nordahl Christoffersen; Department of Clinical Immunology, Zealand University Hospital, Køge, Denmark

Maria Didriksen; Department of Clinical Immunology, Copenhagen University Hospital, Rigshospitalet, Copenhagen, Denmark
Khoa Manh Dinh; Department of Clinical Immunology, Aarhus University Hospital, Aarhus, Denmark
Joseph Dowsett; Department of Clinical Immunology, Copenhagen University Hospital, Rigshospitalet, Copenhagen, Denmark
Christian Erikstrup; Department of Clinical Immunology, Aarhus University Hospital, Aarhus, Denmark; Department of Clinical Medicine, Health, Aarhus University, Aarhus, Denmark
Bjarke Feenstra; Department of Clinical Immunology, Copenhagen University Hospital, Rigshospitalet, Copenhagen, Denmark; Department of Epidemiology Research, Statens Serum Institut, Copenhagen, Denmark
Frank Geller; Department of Clinical Immunology, Copenhagen University Hospital, Rigshospitalet, Copenhagen, Denmark; Department of Epidemiology Research, Statens Serum Institut, Copenhagen, Denmark
Daniel Gudbjartsson; deCODE Genetics, Reykjavik, Iceland
Thomas Folkmann Hansen; Danish Headache Center, Department of Neurology, Copenhagen University Hospital, Rigshospitalet-Glostrup, Copenhagen, Denmark
Dorte Helenius Mikkelsen; Institute of Biological Psychiatry, Mental Health Centre, Sct. Hans, Copenhagen University Hospital, Roskilde, Denmark
Lotte Hindhede; Department of Clinical Immunology, Aarhus University Hospital, Aarhus, Denmark
Henrik Hjalgrim; Danish Cancer Society Research Center, Copenhagen, Denmark; Department of Epidemiology Research, Statens Serum Institut, Copenhagen, Denmark
Jakob Hjorth von Stemann; Department of Clinical Immunology, Copenhagen University Hospital, Rigshospitalet, Copenhagen, Denmark
Bitten Aagaard Jensen; Department of Clinical Immunology, Aalborg University Hospital, Aalborg, Denmark
Andrew Joseph Schork; Institute of Biological Psychiatry, Mental Health Centre, Sct. Hans, Copenhagen University Hospital, Roskilde, Denmark
Kathrine Kaspersen; Department of Clinical Immunology, Aarhus University Hospital, Aarhus, Denmark
Bertram Dalskov Kjerulff; Department of Clinical Immunology, Aarhus University Hospital, Aarhus, Denmark
Mette Kongstad; Department of Clinical Immunology, Copenhagen University Hospital, Rigshospitalet, Copenhagen, Denmark
Susan Mikkelsen; Department of Clinical Immunology, Aarhus University Hospital, Aarhus, Denmark
Christina Mikkelsen; Department of Clinical Immunology, Copenhagen University Hospital, Rigshospitalet, Copenhagen, Denmark
Janna Nissen; Department of Clinical Immunology, Copenhagen University Hospital, Rigshospitalet, Copenhagen, Denmark
Mette Nyegaard; Department of Health Science and Technology, Faculty of Medicine, Aalborg Univeristy, Aalborg, Denmark
Sisse Rye Ostrowski; Department of Clinical Immunology, Copenhagen University Hospital, Rigshospitalet, Copenhagen, Denmark ; Department of Clinical Medicine, Faculty of Health and Medical Sciences, University of Copenhagen, Copenhagen, Denmark
Ole Birger Pedersen; Department of Clinical Immunology, Zealand University Hospital, Køge, Denmark; Department of Clinical Medicine, Faculty of Health and Medical Sciences,

University of Copenhagen, Copenhagen, Denmark
Liam James Elgaard Quinn; Department of Clinical Immunology, Zealand University Hospital, Køge, Denmark
Þórunn Rafnar; deCODE Genetics, Reykjavik, Iceland
Palle Duun Rohde; Department of Health Science and Technology, Faculty of Medicine, Aalborg Univeristy, Aalborg, Denmark
Klaus Rostgaard; Danish Cancer Society Research Center, Copenhagen, Denmark; Department of Epidemiology Research, Statens Serum Institut, Copenhagen, Denmark
Michael Schwinn; Department of Clinical Immunology, Copenhagen University Hospital, Rigshospitalet, Copenhagen, Denmark
Erik Sørensen; Department of Clinical Immunology, Copenhagen University Hospital, Rigshospitalet, Copenhagen, Denmark
Kari Stefansson; deCODE Genetics, Reykjavik, Iceland
Hreinn Stefánsson; deCODE Genetics, Reykjavik, Iceland
Lise Wegner Thørner; Department of Clinical Immunology, Copenhagen University Hospital, Rigshospitalet, Copenhagen, Denmark
Unnur Þorsteinsdóttir; deCODE Genetics, Reykjavik, Iceland
Mie Topholm Bruun; Department of Clinical Immunology, Odense University Hospital, Odense, Denmark
Henrik Ullum; Statens Serum Institut, Copenhagen, Denmark;
Thomas Werge; Institute of Biological Psychiatry, Mental Health Centre, Sct. Hans, Copenhagen University Hospital, Roskilde, Denmark; Department of Clinical Medicine, Faculty of Health and Medical Sciences, University of Copenhagen, Copenhagen, Denmark
David Westergaard; Novo Nordisk Foundation Center for Protein Research, Faculty of Health and Medical Sciences, University of Copenhagen, Copenhagen, Denmark

### Supplementary Figures

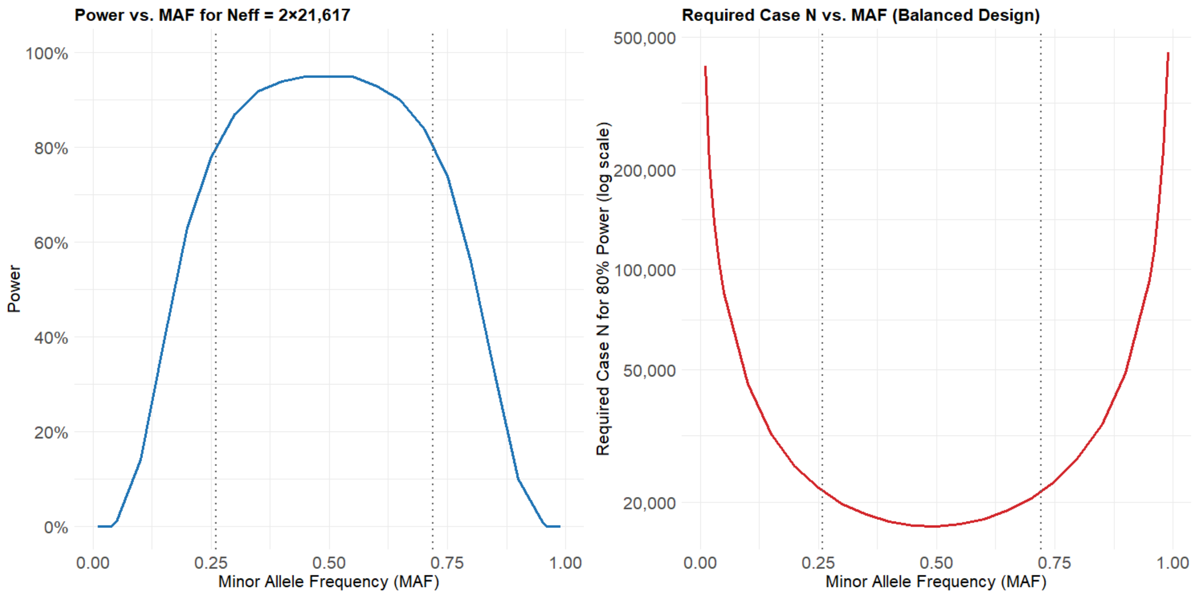


#### Figure S1: Power analysis of the discovery GWAS analysis

The left panel indicates the statistical power to detect effects of 1.1 for the effective N (Neff) of 2x21,617, and in dependence on the effect allele frequency. The right panel indicates the required case N in a balanced design to detect an effect of 1.1 with a power of 80% in depending on the effect allele frequency. Power and the required N were calculated using the Genetic Power Calculator (Purcell, Cherny, and Sham 2003)

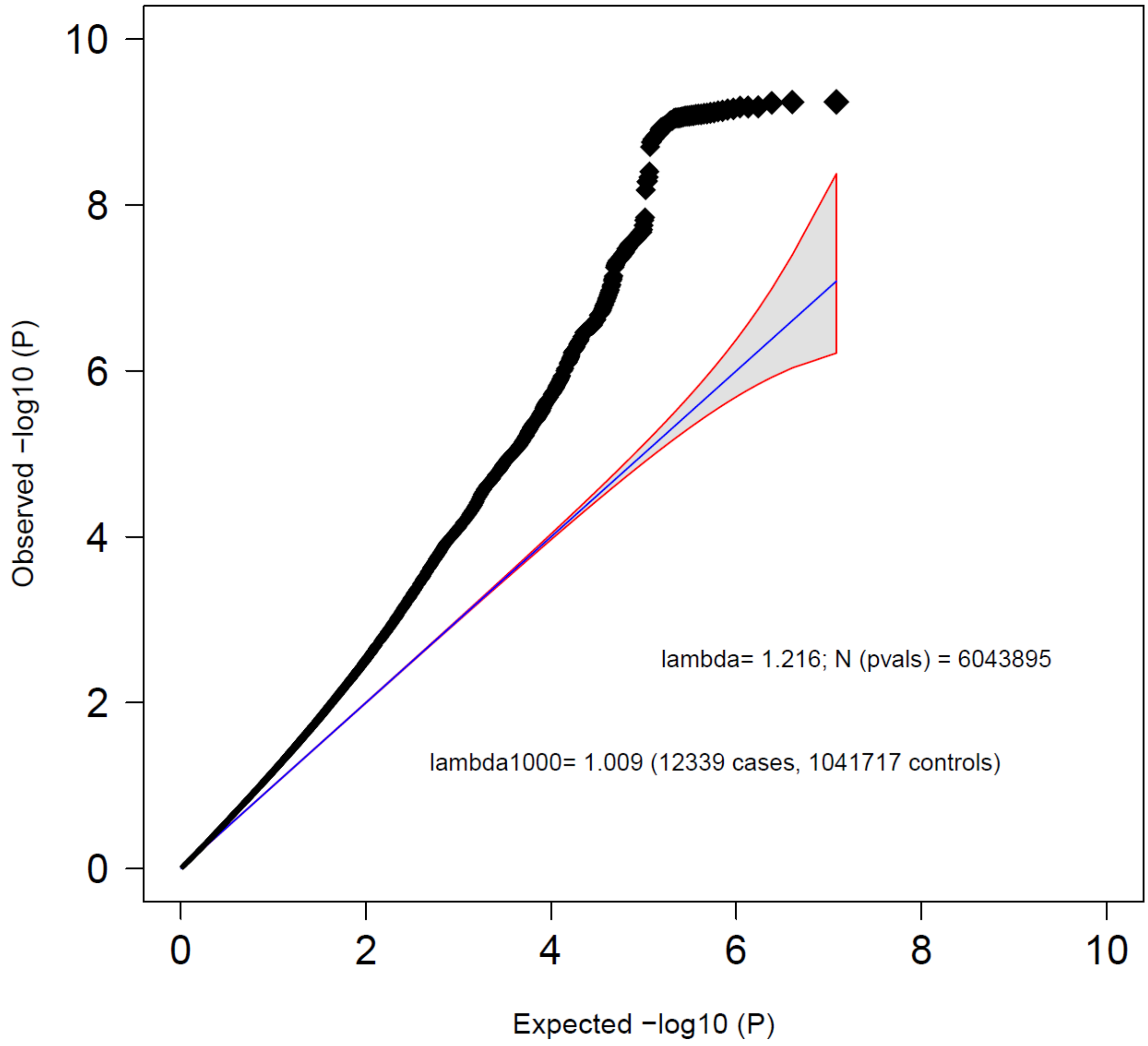


**Figure S2: Quantile–Quantile plot of the GWAS meta-analysis of BPD (Ncases = 12,339, Ncontrols = 1,041,717)**

Observed –log10 *P-values* are shown on the y-axis, and expected –log10 *P-values* are shown on the x-axis. Lambda values were calculated based on the associations of the autosomal variants. The 95% confidence interval of expected P-values under the null hypothesis is indicated by the shaded region.

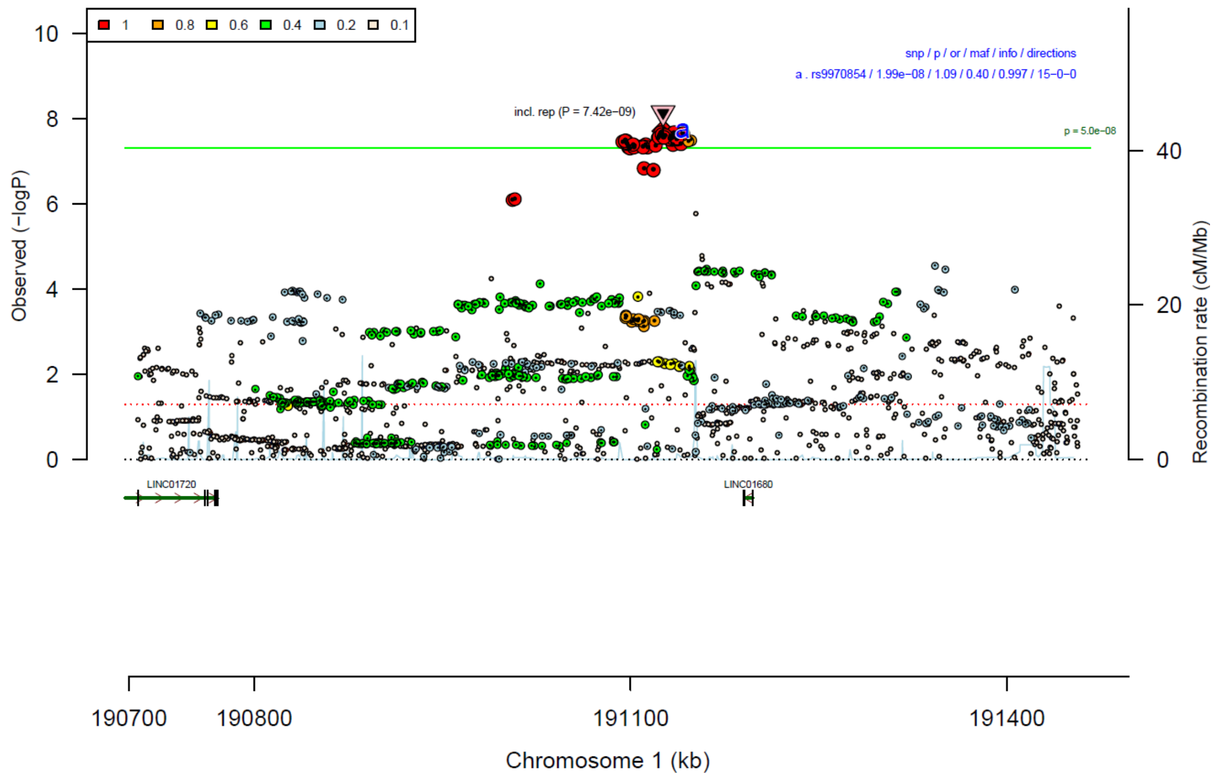


**Figure S3: region plot of rs9970854**

The lead SNP is indicated in diamond shape, and the linkage disequilibrium of the other SNPs ($r^2$) with the index SNP is represented by the colors. The downward-pointing triangle indicated the significance in the combined meta-analysis (discovery & replication). The green line indicates genome-wide significance ($P < 5*10^{-8}$).

| rs9970854 | G/A | | 1:191125935 | | |
|---|---|---|---|---|---|
| | +++++++++++++++ | het_P: | 0.822 | het_I: | 0.0 |
| | **p_value** | **f_ca(n)** | **f_co(n)** | **ln(OR)** | **STDerr** |
| BioVU | 0.84 | 0.595(315) | 0.600(3150) | −0.017 | 0.087 |
| Danish | 0.101 | 0.598(2379) | 0.598(334766) | −0.05 | 0.03 |
| Decode | 0.0471 | 0.577(294) | 0.577(259612) | −0.192 | 0.097 |
| EstBb | 0.0637 | 0.603(351) | 0.640(1755) | −0.166 | 0.09 |
| FinGen | 0.000195 | 0.592(3812) | 0.611(332850) | −0.088 | 0.024 |
| GLAD | 0.0718 | 0.604(1327) | 0.604(6703) | −0.098 | 0.054 |
| HUNT | 0.00381 | 0.604(295) | 0.604(42535) | −0.245 | 0.085 |
| McBb | 0.348 | 0.574(293) | 0.595(46196) | −0.078 | 0.084 |
| MoBa | 0.495 | 0.601(386) | 0.605(7730) | −0.052 | 0.0767 |
| UKBb | 0.51 | 0.567(182) | 0.590(1820) | −0.075 | 0.114 |
| borge | 0.266 | 0.407(1285) | 0.388(1332) | −0.066 | 0.0596 |
| boro1 | 0.184 | 0.465(57) | 0.400(700) | −0.266 | 0.2002 |
| boro2 | 0.8 | 0.421(64) | 0.396(270) | −0.052 | 0.2077 |
| borsm | 0.68 | 0.407(306) | 0.397(759) | −0.042 | 0.1013 |
| boma3 | 0.457 | 0.398(993) | 0.391(1539) | −0.049 | 0.066 |
| **META** | **1.99e−08** | **0.595(12339)** | **0.597(1041717)** | **−0.082** | **0.0145** |
| Replication (AllofUs + borls) | 0.183 | 0.604(685) | 0.599(107750) | −0.073 | 0.0549 |
| **META (including replication)** | **7.42e−09** | **0.595(13024)** | **0.597(1149467)** | **−0.081** | **0.014** |

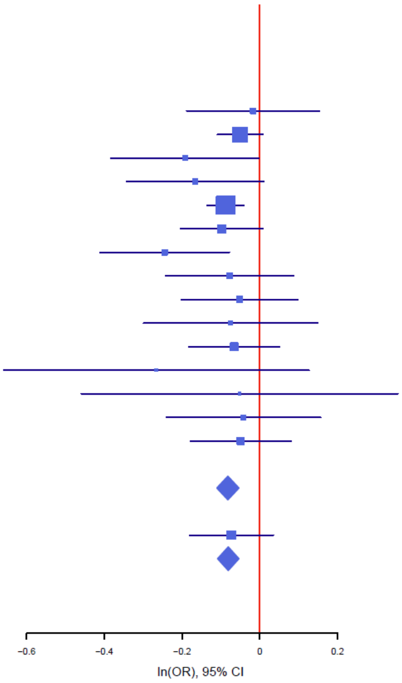


#### Figure S4: forest plot of rs9970854

The plot shows the size and direction of the SNP's effect in each analyzed cohort. Additionally, cohort-specific information and meta-analytic information on the SNP (imputed vs. genotyped SNP (ngt), info score (info), p value, frequency in cases and controls (f_ca(n), f_co(n)) effect size (ln(OR)) and standard error (STDerr)) are given in the table.

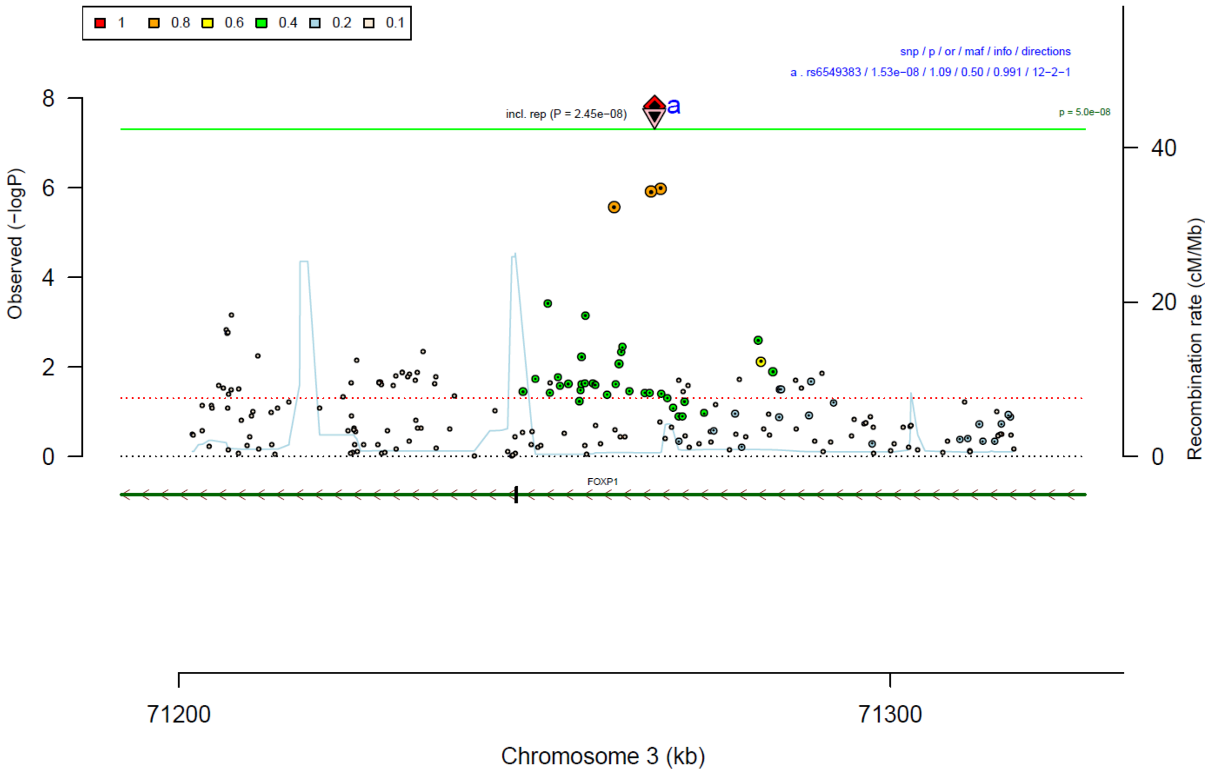


#### Figure S5: region plot of rs6549383

The lead SNP is indicated in diamond shape, and the linkage disequilibrium of the other SNPs ($r^2$) with the index SNP is represented by the colors. The downward-pointing triangle indicated the significance in the combined meta-analysis (discovery & replication). The green line indicates genome-wide significance ($P < 5*10^{-8}$).

| rs6549383 | T/C | | 3:71266885 | | |
|---|---|---|---|---|---|
| | ++−++?++++++−++ | het_P: | 0.2063 | het_I: | 22.8 |
| | **p_value** | **f_ca(n)** | **f_co(n)** | **ln(OR)** | **STDerr** |
| BioVU | 0.93 | 0.510(315) | 0.510(3150) | 0.007 | 0.084 |
| Danish | 0.0157 | 0.498(2379) | 0.498(334766) | 0.073 | 0.03 |
| Decode | 0.695 | 0.542(294) | 0.542(259612) | −0.039 | 0.098 |
| EstBb | 0.534 | 0.456(351) | 0.445(1755) | 0.054 | 0.087 |
| FinGen | 0.00218 | 0.498(3812) | 0.480(332850) | 0.07 | 0.023 |
| HUNT | 0.0985 | 0.495(295) | 0.495(42535) | 0.136 | 0.083 |
| McBb | 0.105 | 0.538(293) | 0.502(46196) | 0.135 | 0.083 |
| MoBa | 0.0331 | 0.555(386) | 0.513(7730) | 0.158 | 0.0742 |
| UKBb | 0.672 | 0.528(182) | 0.509(1820) | 0.048 | 0.112 |
| borge | 0.0248 | 0.506(1285) | 0.469(1332) | 0.132 | 0.0587 |
| boro1 | 0.366 | 0.553(57) | 0.507(700) | 0.193 | 0.2128 |
| boro2 | 0.741 | 0.524(64) | 0.524(270) | 0.069 | 0.2083 |
| borsm | 0.244 | 0.487(306) | 0.523(759) | −0.112 | 0.0961 |
| boma3 | 7.87e−05 | 0.505(993) | 0.466(1539) | 0.251 | 0.0635 |
| **META** | **1.53e−08** | **0.503(11012)** | **0.503(1035014)** | **0.083** | **0.0147** |
| Replication (AllofUs + borls) | 0.664 | 0.499(685) | 0.498(107750) | 0.024 | 0.0543 |
| **META (including replication)** | **2.45e−08** | **0.503(11697)** | **0.503(1142764)** | **0.079** | **0.0142** |

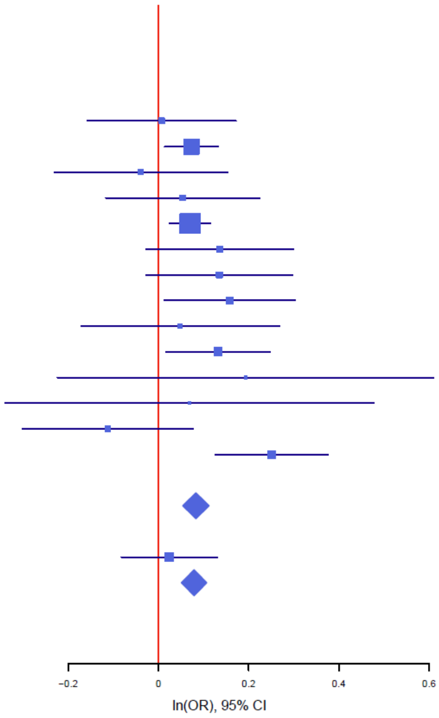


#### Figure S6: forest plot of rs6549383

The plot shows the size and direction of the SNP’s effect in each analyzed cohort. Additionally, cohort-specific information and meta-analytic information on the SNP (imputed vs. genotyped SNP (ngt), info score (info), p value, frequency in cases and controls (f_ca(n), f_co(n)) effect size (ln(OR)) and standard error (STDerr)) are given in the table.

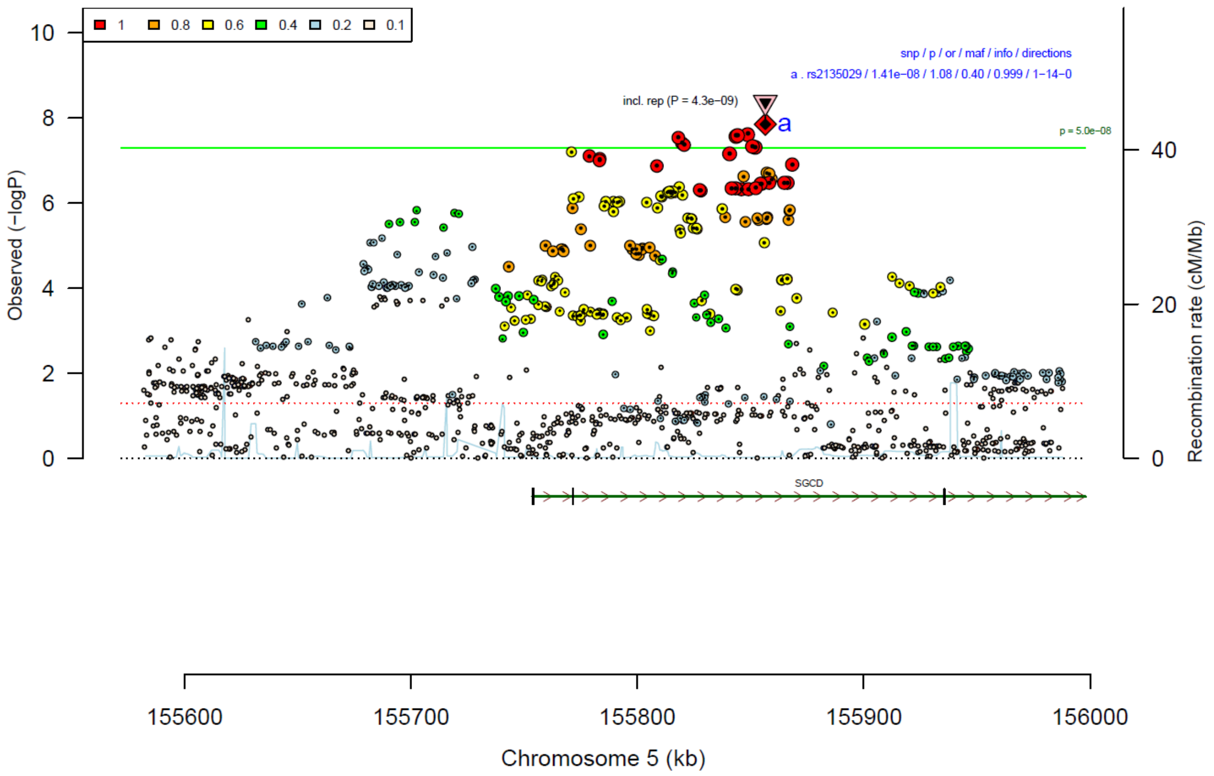


#### Figure S7: region plot of rs2135029

The lead SNP is indicated in diamond shape, and the linkage disequilibrium of the other SNPs ($r^2$) with the index SNP is represented by the colors. The downward-pointing triangle indicated the significance in the combined meta-analysis (discovery & replication). The green line indicates genome-wide significance ($P < 5*10^{-8}$).

| rs2135029 | A/G | | 5:155856538 | | |
|---|---|---|---|---|---|
|  | -----------+---- | het_P: | 0.9158 | het_I: | 0.0 |
| | **p_value** | **f_ca(n)** | **f_co(n)** | **ln(OR)** | **STDerr** |
| BioVU | 0.857 | 0.611(315) | 0.616(3150) | −0.015 | 0.086 |
| Danish | 0.00695 | 0.598(2379) | 0.598(334766) | −0.082 | 0.03 |
| Decode | 0.287 | 0.603(294) | 0.603(259612) | −0.102 | 0.096 |
| EstBb | 0.079 | 0.521(351) | 0.552(1755) | −0.152 | 0.086 |
| FinGen | 0.000287 | 0.577(3812) | 0.595(332850) | −0.086 | 0.023 |
| GLAD | 0.437 | 0.616(1327) | 0.616(6703) | −0.044 | 0.056 |
| HUNT | 0.302 | 0.581(295) | 0.581(42535) | −0.087 | 0.084 |
| McBb | 0.542 | 0.607(293) | 0.617(46196) | −0.051 | 0.085 |
| MoBa | 0.0435 | 0.559(386) | 0.602(7730) | −0.15 | 0.0743 |
| UKBb | 0.13 | 0.577(182) | 0.622(1820) | −0.174 | 0.115 |
| borge | 0.51 | 0.381(1285) | 0.374(1332) | −0.04 | 0.0608 |
| boro1 | 0.203 | 0.377(57) | 0.414(700) | 0.27 | 0.2123 |
| boro2 | 0.521 | 0.422(64) | 0.394(270) | −0.137 | 0.2134 |
| borsm | 0.928 | 0.309(306) | 0.310(759) | −0.01 | 0.1051 |
| boma3 | 0.0793 | 0.413(993) | 0.367(1539) | −0.117 | 0.0664 |
| **META** | **1.41e−08** | **0.594(12339)** | **0.599(1041717)** | **−0.081** | **0.0143** |
| Replication (AllofUs + borls) | 0.141 | 0.630(685) | 0.626(107750) | −0.082 | 0.0559 |
| **META (including replication)** | **4.3e−09** | **0.596(13024)** | **0.602(1149467)** | **−0.081** | **0.0139** |

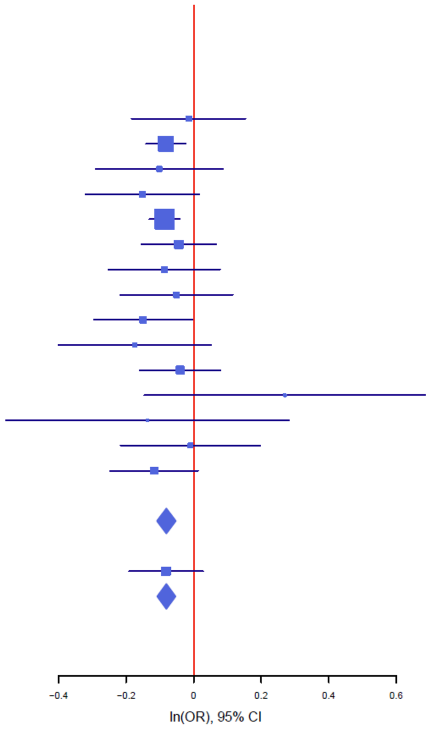


#### Figure S8: forest plot of rs2135029

The plot shows the size and direction of the SNP's effect in each analyzed cohort. Additionally, cohort-specific information and meta-analytic information on the SNP (imputed vs. genotyped SNP (ngt), info score (info), p value, frequency in cases and controls (f_ca(n), f_co(n)) effect size (ln(OR)) and standard error (STDerr)) are given in the table.

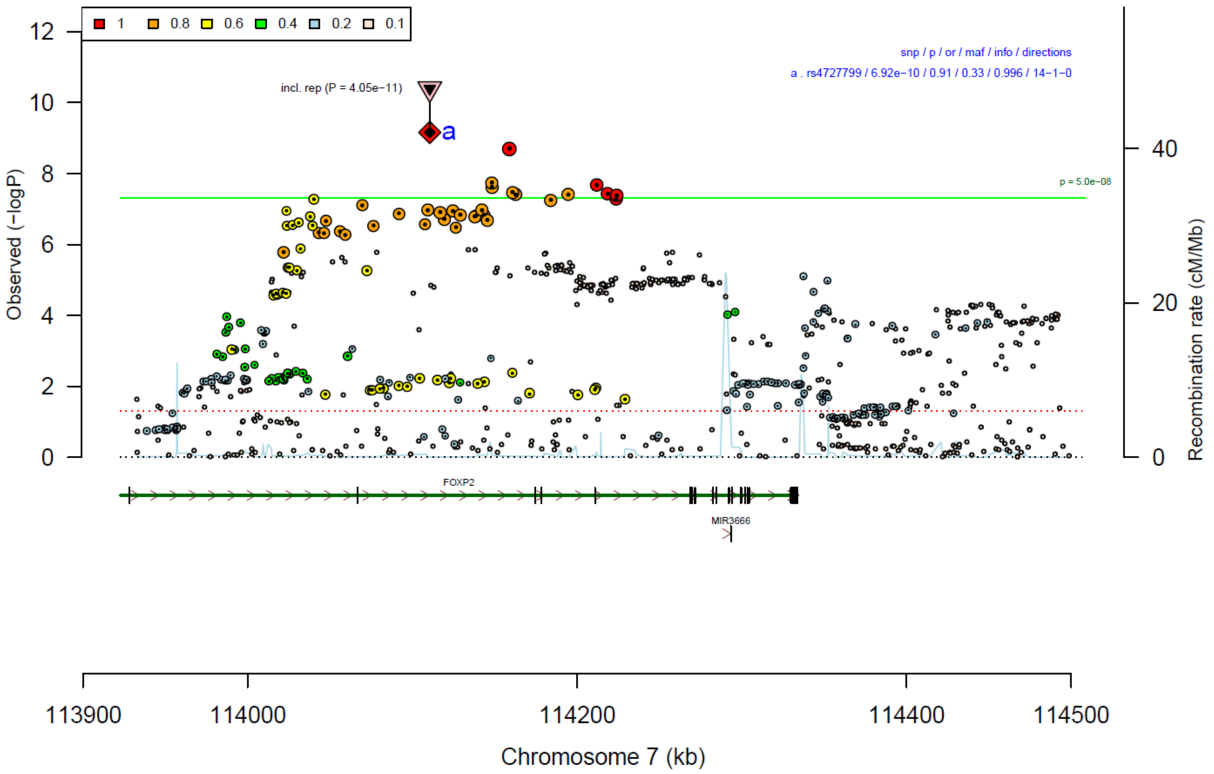


#### Figure S9: region plot of rs4727799

The lead SNP is indicated in diamond shape, and the linkage disequilibrium of the other SNPs ($r^2$) with the index SNP is represented by the colors. The downward-pointing triangle indicated the significance in the combined meta-analysis (discovery & replication). The green line indicates genome-wide significance ($P < 5*10^{-8}$).

| rs4727799 | T/C | | 7:114110568 | | |
|---|---|---|---|---|---|
| | +++++++++−+++++ | het_P: | 0.03764 | het_I: | 43.3 |
| | **p_value** | **f_ca(n)** | **f_co(n)** | **ln(OR)** | **STDerr** |
| BioVU | 0.541 | 0.665(315) | 0.653(3150) | 0.054 | 0.087 |
| Danish | 0.000928 | 0.655(2379) | 0.655(334766) | 0.105 | 0.032 |
| Decode | 0.187 | 0.684(294) | 0.684(259612) | 0.137 | 0.104 |
| EstBb | 0.707 | 0.639(351) | 0.630(1755) | 0.032 | 0.087 |
| FinGen | 0.0549 | 0.685(3812) | 0.677(332850) | 0.048 | 0.025 |
| GLAD | 0.171 | 0.646(1327) | 0.646(6703) | 0.077 | 0.056 |
| HUNT | 0.00142 | 0.686(295) | 0.686(42535) | 0.284 | 0.089 |
| McBb | 0.0854 | 0.680(293) | 0.642(46196) | 0.154 | 0.09 |
| MoBa | 0.166 | 0.700(386) | 0.672(7730) | 0.112 | 0.0809 |
| UKBb | 0.764 | 0.635(182) | 0.644(1820) | −0.035 | 0.116 |
| borge | 0.098 | 0.366(1285) | 0.392(1332) | 0.1 | 0.0603 |
| boro1 | 0.0707 | 0.254(57) | 0.331(700) | 0.428 | 0.2367 |
| boro2 | 0.000916 | 0.219(64) | 0.363(270) | 0.857 | 0.2585 |
| borsm | 0.122 | 0.399(306) | 0.433(759) | 0.155 | 0.1 |
| boma3 | 0.00198 | 0.353(993) | 0.397(1539) | 0.205 | 0.0663 |
| **META** | **6.92e−10** | **0.663(12339)** | **0.670(1041717)** | **0.093** | **0.0151** |
| Replication (AllofUs + borls) | 0.0147 | 0.634(685) | 0.631(107750) | 0.14 | 0.0575 |
| **META (including replication)** | **4.05e−11** | **0.661(13024)** | **0.666(1149467)** | **0.096** | **0.0146** |

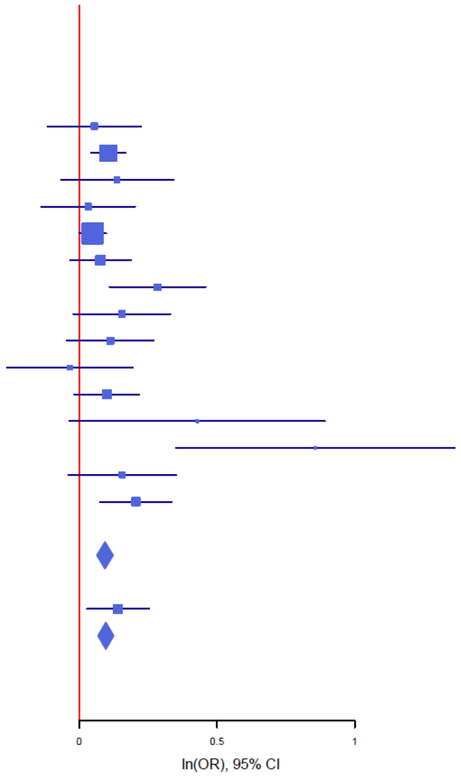


#### Figure S10: forest plot of rs4727799

The plot shows the size and direction of the SNP's effect in each analyzed cohort. Additionally, cohort-specific information and meta-analytic information on the SNP (imputed vs. genotyped SNP (ngt), info score (info), p value, frequency in cases and controls (f_ca(n), f_co(n)) effect size (ln(OR)) and standard error (STDerr)) are given in the table.

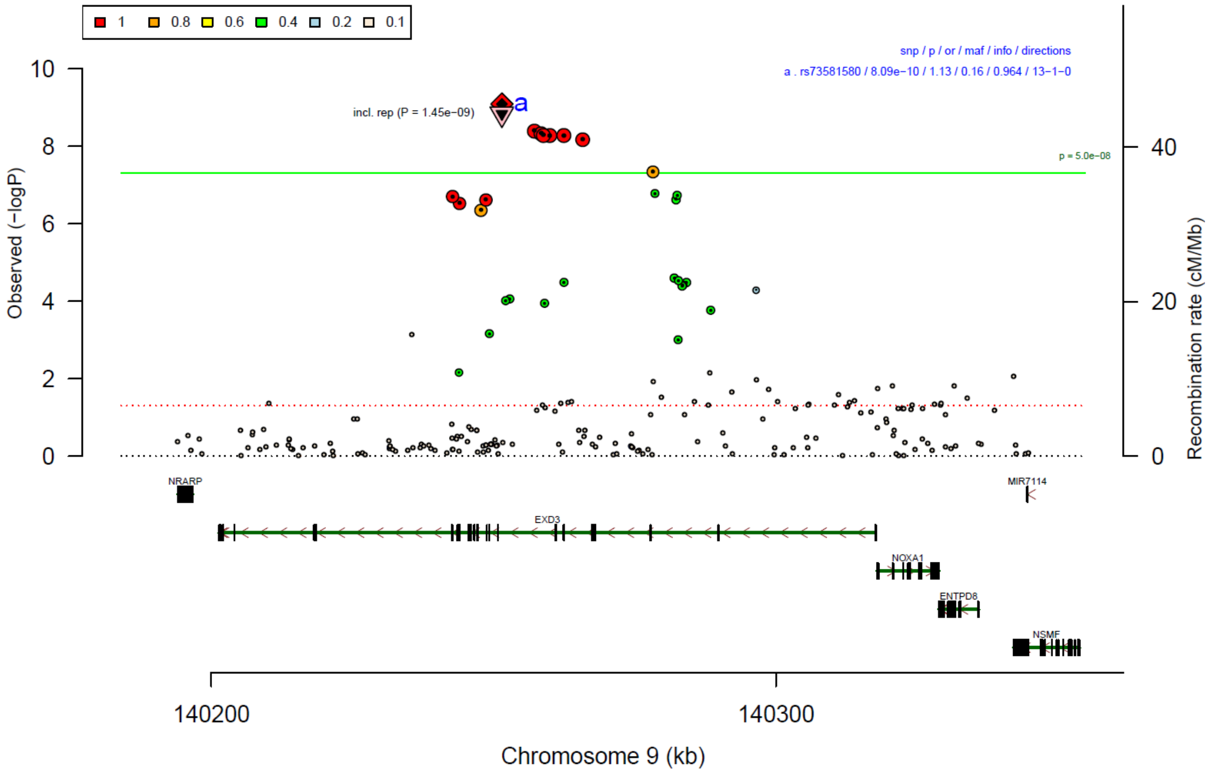


#### Figure S11: region plot of rs73581580

The lead SNP is indicated in diamond shape, and the linkage disequilibrium of the other SNPs ($r^2$) with the index SNP is represented by the colors. The downward-pointing triangle indicated the significance in the combined meta-analysis (discovery & replication). The green line indicates genome-wide significance ($P < 5*10^{-8}$).

| **rs73581580** | **G/A** | | **9:140251458** | | |
|---|---|---|---|---|---|
| | +++−+++++?+++++ | het_P: | 0.7786 | het_I: | 0.0 |
| | **p_value** | **f_ca(n)** | **f_co(n)** | **ln(OR)** | **STDerr** |
| BioVU | 0.583 | 0.865(315) | 0.871(3150) | −0.069 | 0.125 |
| Danish | 0.000386 | 0.852(2379) | 0.852(334766) | −0.149 | 0.042 |
| Decode | 0.296 | 0.865(294) | 0.865(259612) | −0.143 | 0.137 |
| EstBb | 0.452 | 0.872(351) | 0.861(1755) | 0.093 | 0.124 |
| FinGen | 0.000742 | 0.776(3812) | 0.795(332850) | −0.093 | 0.028 |
| GLAD | 0.378 | 0.882(1327) | 0.882(6703) | −0.071 | 0.081 |
| HUNT | 0.0998 | 0.870(295) | 0.870(42535) | −0.201 | 0.122 |
| McBb | 0.125 | 0.855(293) | 0.875(46196) | −0.179 | 0.117 |
| MoBa | 0.0619 | 0.830(237) | 0.859(4750) | −0.237 | 0.1269 |
| borge | 0.0707 | 0.868(1285) | 0.887(1332) | −0.161 | 0.0893 |
| boro1 | 0.509 | 0.866(57) | 0.875(700) | −0.202 | 0.306 |
| boro2 | 0.139 | 0.828(64) | 0.870(270) | −0.407 | 0.2752 |
| borsm | 0.0985 | 0.890(306) | 0.913(759) | −0.271 | 0.1639 |
| boma3 | 0.0748 | 0.858(993) | 0.884(1539) | −0.17 | 0.0952 |
| **META** | **8.09e−10** | **0.836(12008)** | **0.839(1036917)** | **−0.119** | **0.0193** |
| Replication (AllofUs + borls) | 0.798 | 0.886(685) | 0.884(107750) | −0.021 | 0.0834 |
| **META (including replication)** | **1.45e−09** | **0.839(12693)** | **0.843(1144667)** | **−0.114** | **0.0188** |

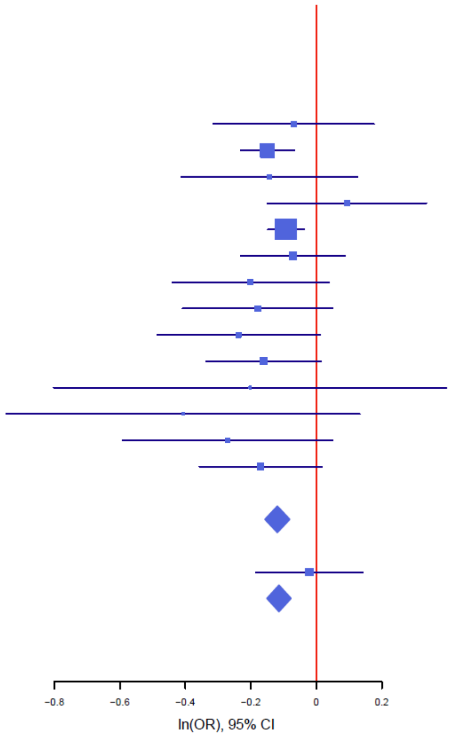


#### Figure S12: forest plot of rs73581580

The plot shows size and direction of the SNP's effect in each analyzed cohort. Additionally, cohort-specific information and meta-analytic information on the SNP (imputed vs. genotyped SNP (ngt), info score (info), p value, frequency in cases and controls (f_ca(n), f_co(n)) effect size (ln(OR)) and standard error (STDerr)) are given in the table.

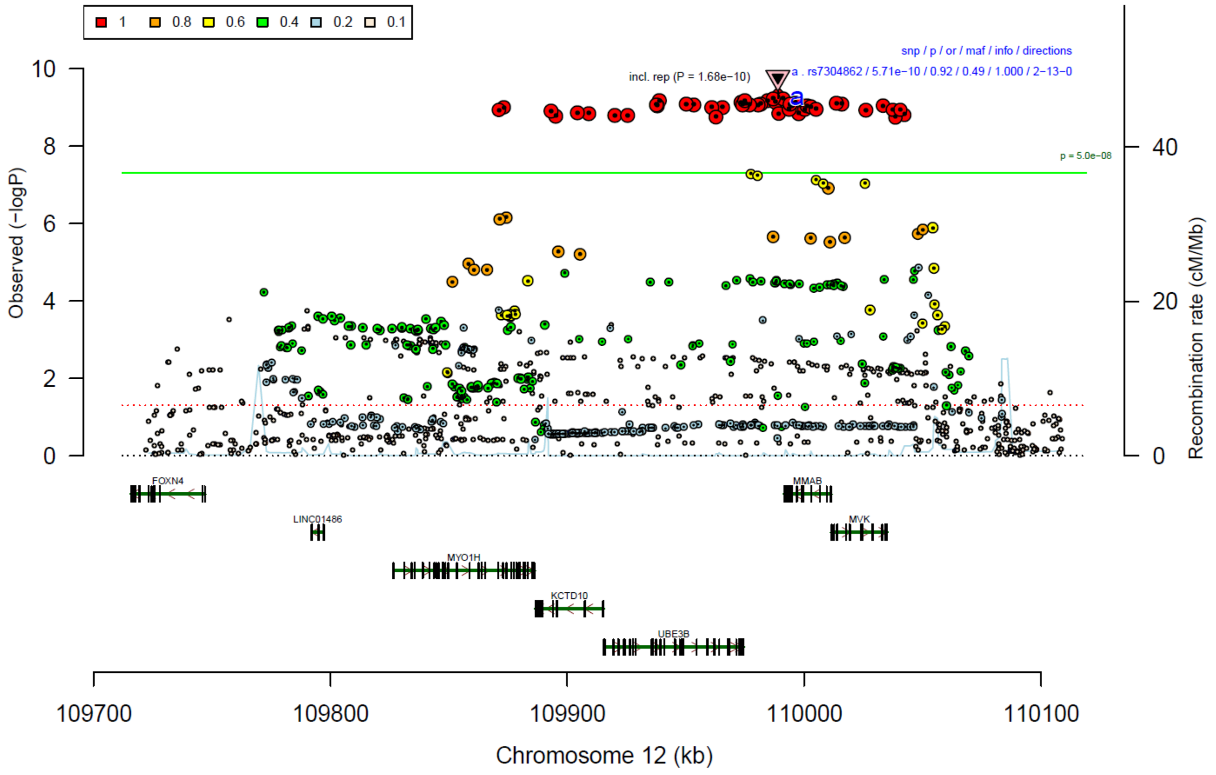


**Figure S13: region plot of rs7304862**

The lead SNP is indicated in diamond shape, and the linkage disequilibrium of the other SNPs ($r^2$) with the index SNP is represented by the colors. The downward-pointing triangle indicated the significance in the combined meta-analysis (discovery & replication). The green line indicates genome-wide significance ($P < 5*10^{-8}$).

| rs7304862 | G/T | | 12:109988891 | | |
|---|---|---|---|---|---|
| | −−−−−−−−−−++−−− | het_P: | 0.02254 | het_I: | 47.1 |
| | **p_value** | **f_ca(n)** | **f_co(n)** | **ln(OR)** | **STDerr** |
| BioVU | 0.75 | 0.544(315) | 0.537(3150) | 0.027 | 0.085 |
| Danish | 0.0109 | 0.537(2379) | 0.537(334766) | 0.077 | 0.03 |
| Decode | 0.033 | 0.522(294) | 0.522(259612) | 0.209 | 0.098 |
| EstBb | 0.45 | 0.583(351) | 0.566(1755) | 0.065 | 0.085 |
| FinGen | 0.00187 | 0.491(3812) | 0.474(332850) | 0.072 | 0.023 |
| GLAD | 1.46e−07 | 0.528(1327) | 0.528(6703) | 0.273 | 0.052 |
| HUNT | 0.277 | 0.526(295) | 0.526(42535) | 0.09 | 0.083 |
| McBb | 0.55 | 0.549(293) | 0.535(46196) | 0.05 | 0.083 |
| MoBa | 0.0804 | 0.558(386) | 0.534(7730) | 0.132 | 0.0756 |
| UKBb | 0.221 | 0.561(182) | 0.526(1820) | 0.137 | 0.112 |
| borge | 0.649 | 0.452(1285) | 0.451(1332) | 0.027 | 0.059 |
| boro1 | 0.183 | 0.517(57) | 0.458(700) | −0.27 | 0.2026 |
| boro2 | 0.0656 | 0.578(64) | 0.482(270) | −0.384 | 0.2086 |
| borsm | 0.903 | 0.409(306) | 0.409(759) | 0.012 | 0.099 |
| boma3 | 0.0413 | 0.415(993) | 0.451(1539) | 0.131 | 0.0642 |
| **META** | **5.71e−10** | **0.529(12339)** | **0.513(1041717)** | **0.088** | **0.0142** |
| Replication (AllofUs + borls) | 0.114 | 0.538(685) | 0.539(107750) | 0.087 | 0.0549 |
| **META (including replication)** | **1.68e−10** | **0.529(13024)** | **0.515(1149467)** | **0.088** | **0.0137** |

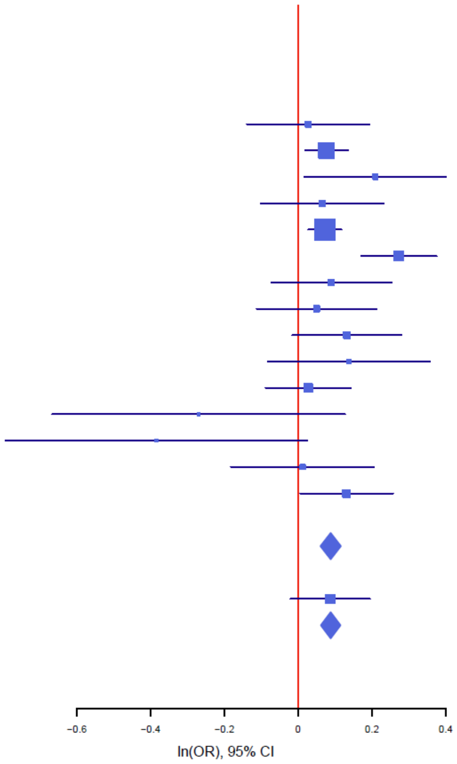


#### Figure S14: forest plot of rs7304862

The plot shows the size and direction of the SNP's effect in each analyzed cohort. Additionally, cohort-specific information and meta-analytic information on the SNP (imputed vs. genotyped SNP (ngt), info score (info), p value, frequency in cases and controls (f_ca(n), f_co(n)) effect size (ln(OR)) and standard error (STDerr)) are given in the table.

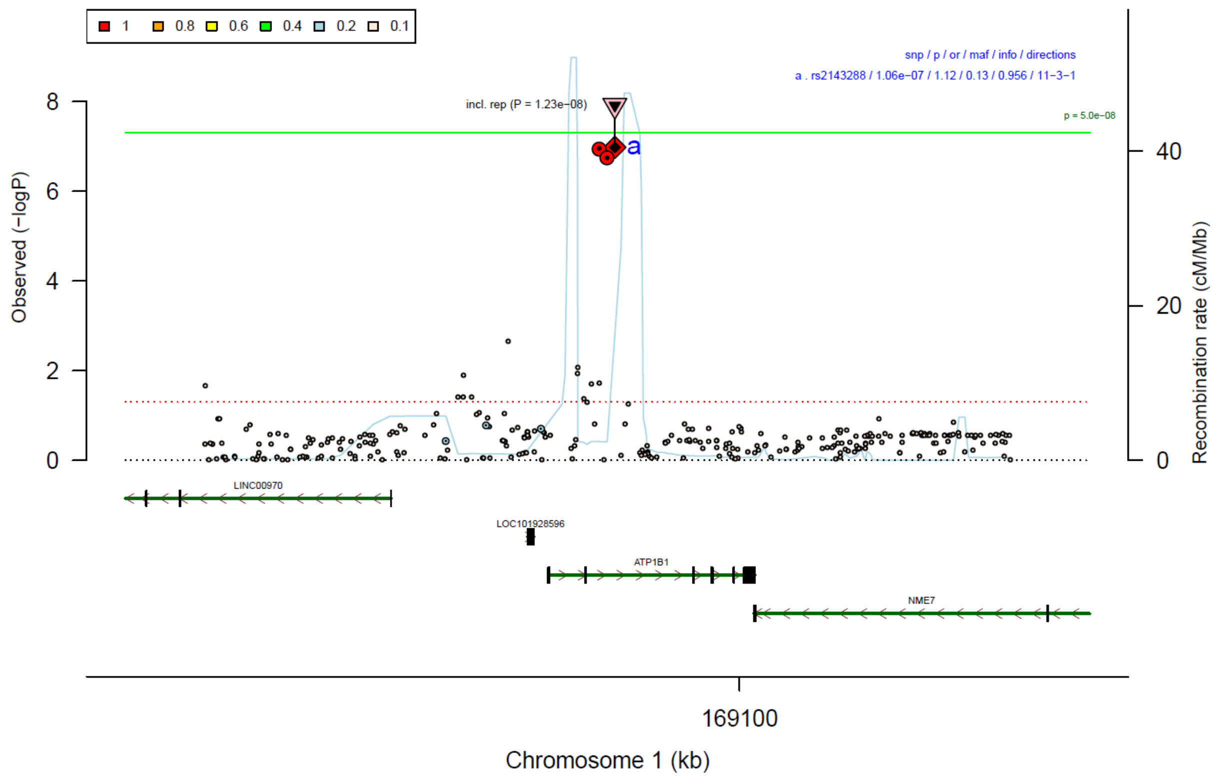


#### Figure S15: region plot of rs2143288

The lead SNP is indicated in diamond shape, and the linkage disequilibrium of the other SNPs ($r^2$) with the index SNP is represented by the colors. The downward-pointing triangle indicated the significance in the combined meta-analysis (discovery & replication). The green line indicates genome-wide significance ($P < 5*10^{-8}$).

| **rs2143288** | **G/A** | | **1:169084340** | | |
|---|---|---|---|---|---|
| | −++++?++−+++++− | het_P: | 0.0436 | het_I: | 43.1 |
| | **p_value** | **f_ca(n)** | **f_co(n)** | **ln(OR)** | **STDerr** |
| BioVU | 0.759 | 0.853(315) | 0.847(3150) | 0.036 | 0.117 |
| Danish | 0.012 | 0.863(2379) | 0.863(334766) | −0.111 | 0.044 |
| Decode | 0.442 | 0.864(294) | 0.864(259612) | −0.103 | 0.134 |
| EstBb | 0.000699 | 0.817(351) | 0.863(1755) | −0.44 | 0.13 |
| FinGen | 0.000645 | 0.867(3812) | 0.880(332850) | −0.117 | 0.034 |
| HUNT | 0.718 | 0.853(295) | 0.853(42535) | −0.046 | 0.127 |
| McBb | 0.089 | 0.839(293) | 0.862(46196) | −0.192 | 0.113 |
| MoBa | 0.492 | 0.889(237) | 0.880(4750) | 0.106 | 0.1546 |
| UKBb | 0.263 | 0.840(182) | 0.865(1820) | −0.179 | 0.16 |
| borge | 0.212 | 0.848(1285) | 0.855(1332) | −0.114 | 0.0916 |
| boro1 | 0.00208 | 0.784(57) | 0.884(700) | −0.836 | 0.2716 |
| boro2 | 0.0896 | 0.836(64) | 0.889(270) | −0.533 | 0.3137 |
| borsm | 0.174 | 0.825(306) | 0.848(759) | −0.199 | 0.1464 |
| boma3 | 0.561 | 0.855(993) | 0.856(1539) | 0.061 | 0.1058 |
| **META** | **1.06e−07** | **0.858(10863)** | **0.868(1032034)** | **−0.117** | **0.022** |
| Replication (AllofUs + borls) | 0.035 | 0.842(685) | 0.845(107750) | −0.154 | 0.073 |
| **META (including replication)** | **1.23e−08** | **0.857(11548)** | **0.866(1139784)** | **−0.12** | **0.0211** |

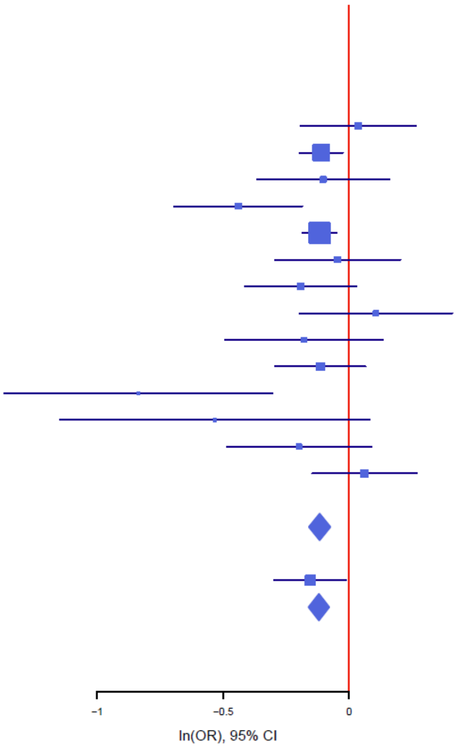


**Figure S16: forest plot of rs2143288**

The plot shows the size and direction of the SNP's effect in each analyzed cohort. Additionally, cohort-specific information and meta-analytic information on the SNP (imputed vs. genotyped SNP (ngt), info score (info), p value, frequency in cases and controls (f_ca(n), f_co(n)) effect size (ln(OR)) and standard error (STDerr)) are given in the table.

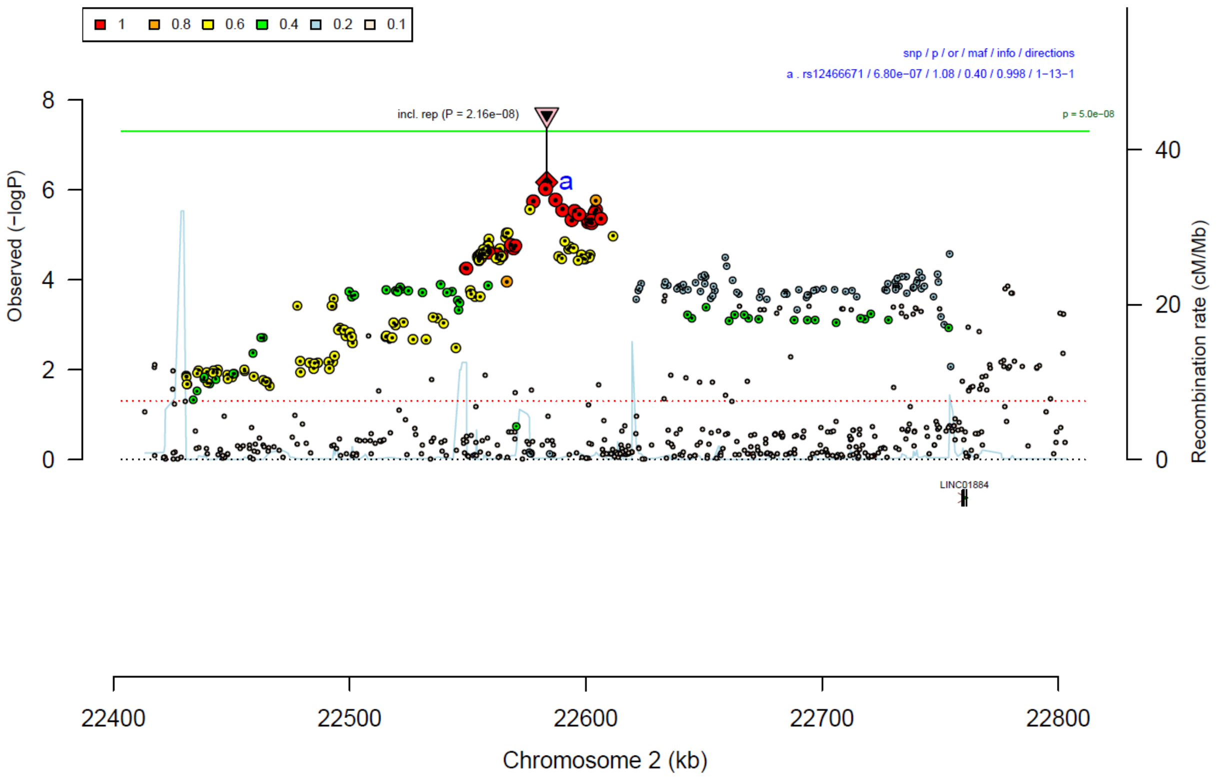


**Figure S17: region plot of rs12466671**

The lead SNP is indicated in diamond shape, and the linkage disequilibrium of the other SNPs ($r^2$) with the index SNP is represented by the colors. The downward-pointing triangle indicated the significance in the combined meta-analysis (discovery & replication). The green line indicates genome-wide significance ($P < 5*10^{-8}$).

| rs12466671 | A/G | | 2:22583435 | | |
|---|---|---|---|---|---|
|  | ?------+-------- | het_P: | 0.8165 | het_I: | 0.0 |
| | **p_value** | **f_ca(n)** | **f_co(n)** | **ln(OR)** | **STDerr** |
| Danish | 0.0021 | 0.605(2379) | 0.605(334766) | −0.093 | 0.03 |
| Decode | 0.748 | 0.632(294) | 0.632(259612) | −0.031 | 0.098 |
| EstBb | 0.734 | 0.605(351) | 0.610(1755) | −0.03 | 0.089 |
| FinGen | 0.00562 | 0.540(3812) | 0.560(332850) | −0.064 | 0.023 |
| GLAD | 0.00221 | 0.620(1327) | 0.620(6703) | −0.167 | 0.054 |
| HUNT | 0.405 | 0.625(295) | 0.625(42535) | −0.071 | 0.086 |
| McBb | 0.68 | 0.625(293) | 0.615(46196) | 0.035 | 0.085 |
| MoBa | 0.322 | 0.585(237) | 0.613(4750) | −0.095 | 0.096 |
| UKBb | 0.0877 | 0.591(182) | 0.634(1820) | −0.195 | 0.114 |
| borge | 0.546 | 0.605(1285) | 0.614(1332) | −0.037 | 0.0618 |
| boro1 | 0.833 | 0.575(57) | 0.605(700) | −0.043 | 0.2048 |
| boro2 | 0.444 | 0.600(64) | 0.644(270) | −0.17 | 0.2222 |
| borsm | 0.856 | 0.635(306) | 0.637(759) | −0.019 | 0.1048 |
| boma3 | 0.793 | 0.596(993) | 0.609(1539) | −0.017 | 0.0661 |
| **META** | **6.8e−07** | **0.587(11875)** | **0.599(1035587)** | **−0.073** | **0.0146** |
| Replication (AllofUs + borls) | 0.00237 | 0.616(685) | 0.619(107750) | −0.167 | 0.055 |
| **META (including replication)** | **2.16e−08** | **0.589(12560)** | **0.601(1143337)** | **−0.079** | **0.0141** |

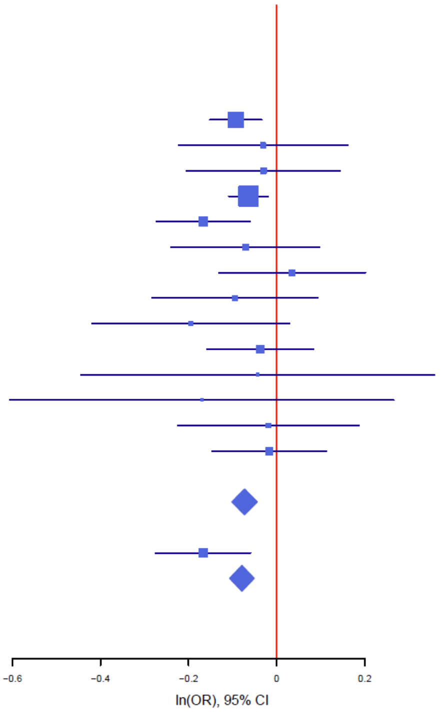


#### Figure S18: forest plot of rs12466671

The plot shows the size and direction of the SNP’s effect in each analyzed cohort. Additionally, cohort-specific information and meta-analytic information on the SNP (imputed vs. genotyped SNP (ngt), info score (info), p value, frequency in cases and controls (f_ca(n), f_co(n)) effect size (ln(OR)) and standard error (STDerr)) are given in the table.

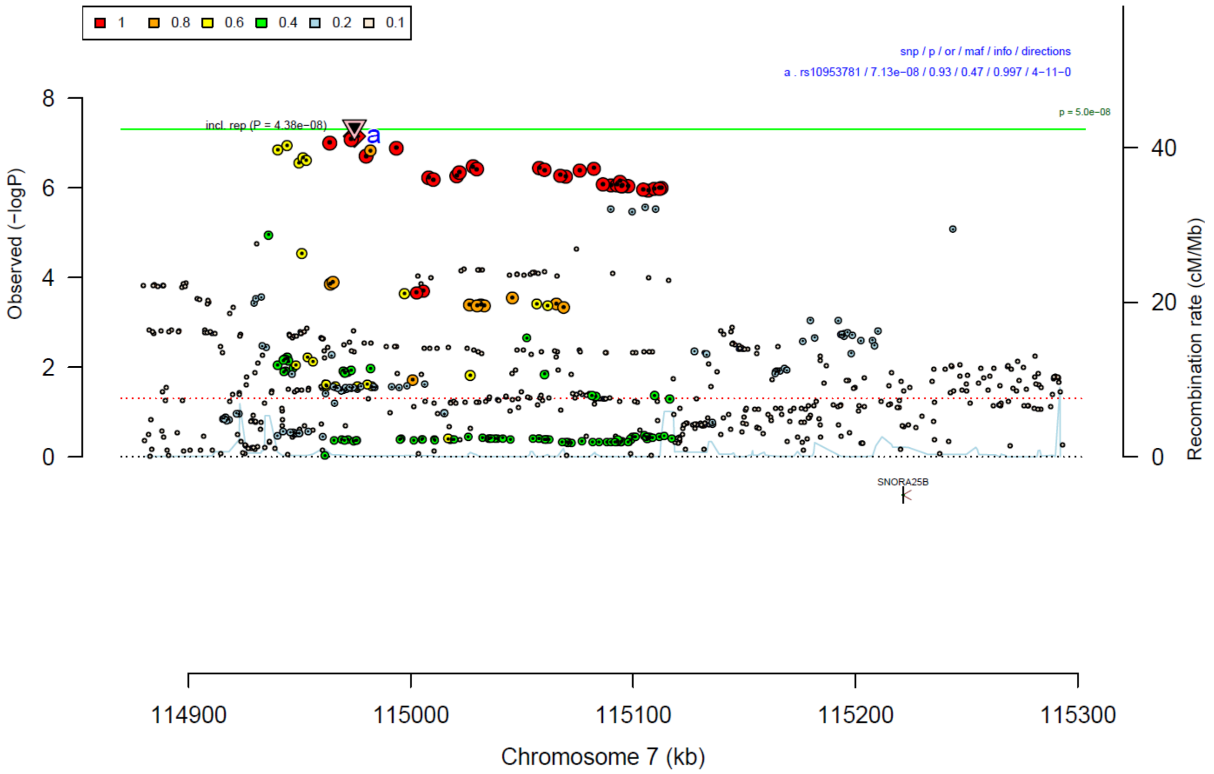


#### Figure S19: region plot of rs10953781

The lead SNP is indicated in diamond shape, and the linkage disequilibrium of the other SNPs ($r^2$) with the index SNP is represented by the colors. The downward-pointing triangle indicated the significance in the combined meta-analysis (discovery & replication). The green line indicates genome-wide significance ($P < 5*10^{-8}$).

**rs10953781** **C/T** **7:114974695**

--+--------+--++ het_P: 0.442 het_I: 0.7

| | p_value | f_ca(n) | f_co(n) | ln(OR) | STDerr |
|---|---|---|---|---|---|
| BioVU | 0.676 | 0.511(315) | 0.502(3150) | 0.035 | 0.085 |
| Danish | 0.00672 | 0.507(2379) | 0.507(334766) | 0.082 | 0.03 |
| Decode | 0.969 | 0.546(294) | 0.546(259612) | −0.004 | 0.103 |
| EstBb | 0.225 | 0.505(351) | 0.480(1755) | 0.103 | 0.086 |
| FinGen | 0.000106 | 0.587(3812) | 0.562(332850) | 0.091 | 0.023 |
| GLAD | 0.289 | 0.504(1327) | 0.504(6703) | 0.056 | 0.053 |
| HUNT | 0.00126 | 0.506(295) | 0.506(42535) | 0.268 | 0.083 |
| McBb | 0.601 | 0.517(293) | 0.501(46196) | 0.044 | 0.083 |
| MoBa | 0.423 | 0.523(386) | 0.510(7730) | 0.06 | 0.0746 |
| UKBb | 0.0737 | 0.531(182) | 0.476(1820) | 0.209 | 0.117 |
| borge | 0.753 | 0.484(1285) | 0.488(1332) | −0.018 | 0.0588 |
| boro1 | 0.563 | 0.500(57) | 0.519(700) | −0.119 | 0.2051 |
| boro2 | 0.871 | 0.520(64) | 0.509(270) | 0.035 | 0.2151 |
| borsm | 0.119 | 0.465(306) | 0.433(759) | 0.154 | 0.0987 |
| boma3 | 0.926 | 0.478(993) | 0.483(1539) | −0.006 | 0.0642 |
| **META** | **7.13e−08** | **0.528(12339)** | **0.534(1041717)** | **0.077** | **0.0142** |
| Replication (AllofUs + borls) | 0.313 | 0.481(685) | 0.492(107750) | 0.055 | 0.0546 |
| **META (including replication)** | **4.38e−08** | **0.526(13024)** | **0.530(1149467)** | **0.075** | **0.0137** |

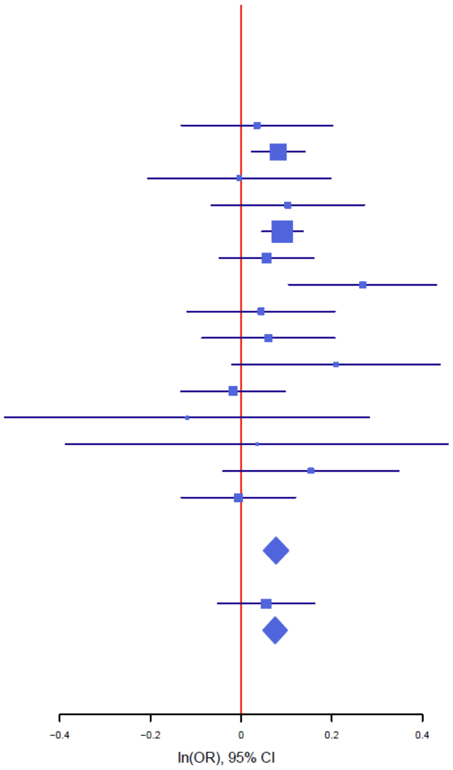


#### Figure S20: forest plot of rs10953781

The plot shows the size and direction of the SNP's effect in each analyzed cohort. Additionally, cohort-specific information and meta-analytic information on the SNP (imputed vs. genotyped SNP (ngt), info score (info), p value, frequency in cases and controls (f_ca(n), f_co(n)) effect size (ln(OR)) and standard error (STDerr)) are given in the table.

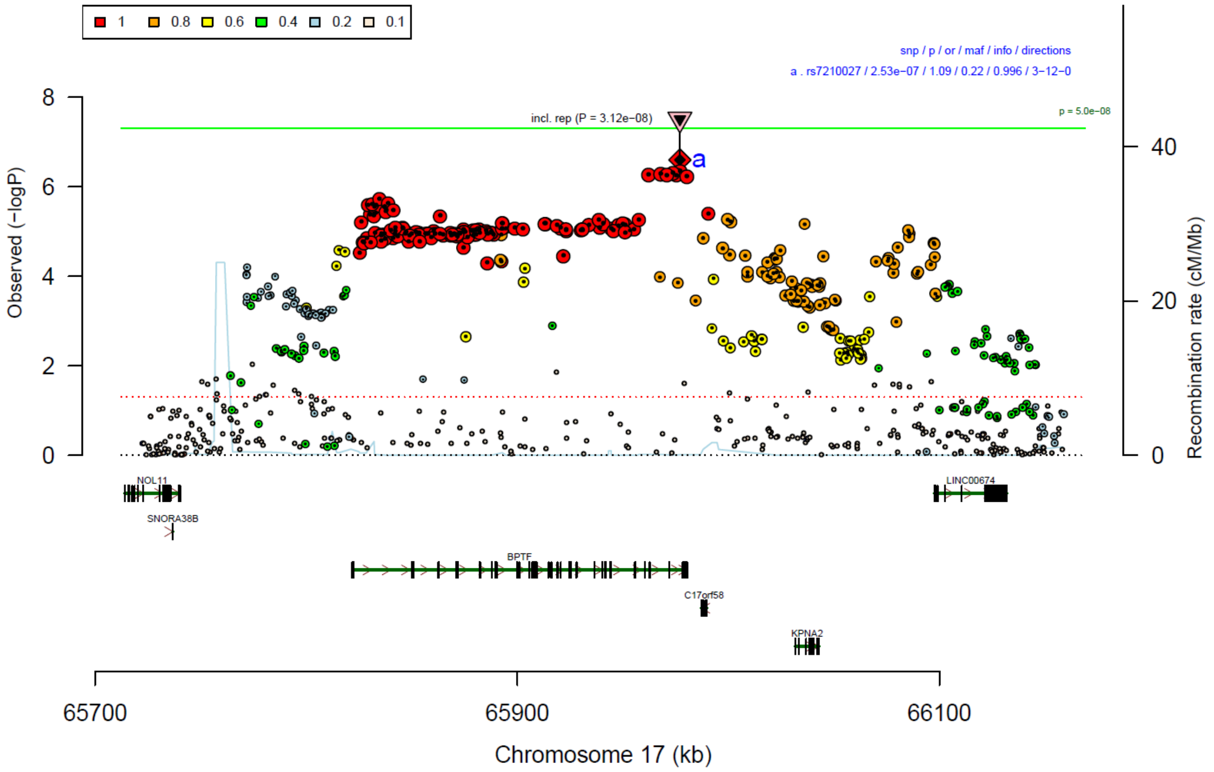


#### Figure S21: region plot of rs7210027

The lead SNP is indicated in diamond shape, and the linkage disequilibrium of the other SNPs ($r^2$) with the index SNP is represented by the colors. The downward-pointing triangle indicated the significance in the combined meta-analysis (discovery & replication). The green line indicates genome-wide significance ($P < 5*10^{-8}$).

| rs7210027 | T/C | | 17:65977053 | | |
|---|---|---|---|---|---|
|  | +--+------+----- | het_P: | 0.1773 | het_I: | 25.1 |
| | **p_value** | **f_ca(n)** | **f_co(n)** | **ln(OR)** | **STDerr** |
| BioVU | 0.104 | 0.802(315) | 0.775(3150) | 0.164 | 0.101 |
| Danish | 0.00572 | 0.781(2379) | 0.781(334766) | −0.099 | 0.036 |
| Decode | 0.095 | 0.814(294) | 0.814(259612) | −0.198 | 0.119 |
| EstBb | 0.323 | 0.788(351) | 0.771(1755) | 0.099 | 0.1 |
| FinGen | 0.00196 | 0.742(3812) | 0.759(332850) | −0.082 | 0.026 |
| GLAD | 0.0142 | 0.791(1327) | 0.791(6703) | −0.163 | 0.066 |
| HUNT | 0.716 | 0.787(295) | 0.787(42535) | −0.037 | 0.1 |
| McBb | 0.246 | 0.745(293) | 0.769(46196) | −0.112 | 0.096 |
| MoBa | 0.118 | 0.764(386) | 0.789(7730) | −0.137 | 0.0876 |
| UKBb | 0.241 | 0.829(182) | 0.804(1820) | 0.176 | 0.15 |
| borge | 0.187 | 0.224(1285) | 0.215(1332) | −0.095 | 0.0723 |
| boro1 | 0.172 | 0.247(57) | 0.203(700) | −0.333 | 0.2439 |
| boro2 | 0.313 | 0.242(64) | 0.209(270) | −0.258 | 0.2563 |
| borsm | 0.783 | 0.203(306) | 0.191(759) | −0.034 | 0.1247 |
| boma3 | 0.0219 | 0.230(993) | 0.211(1539) | −0.178 | 0.0775 |
| **META** | **2.53e−07** | **0.770(12339)** | **0.782(1041717)** | **−0.086** | **0.0167** |
| Replication (AllofUs + borls) | 0.0307 | 0.782(685) | 0.782(107750) | −0.138 | 0.064 |
| **META (including replication)** | **3.12e−08** | **0.771(13024)** | **0.782(1149467)** | **−0.089** | **0.0162** |

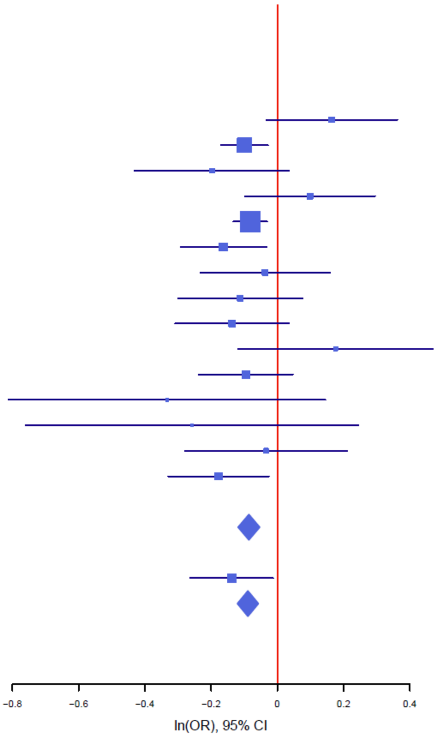


#### Figure S22: forest plot of rs7210027

The plot shows the size and direction of the SNP’s effect in each analyzed cohort. Additionally, cohort-specific information and meta-analytic information on the SNP (imputed vs. genotyped SNP (ngt), info score (info), p value, frequency in cases and controls (f_ca(n), f_co(n)) effect size (ln(OR)) and standard error (STDerr)) are given in the table.

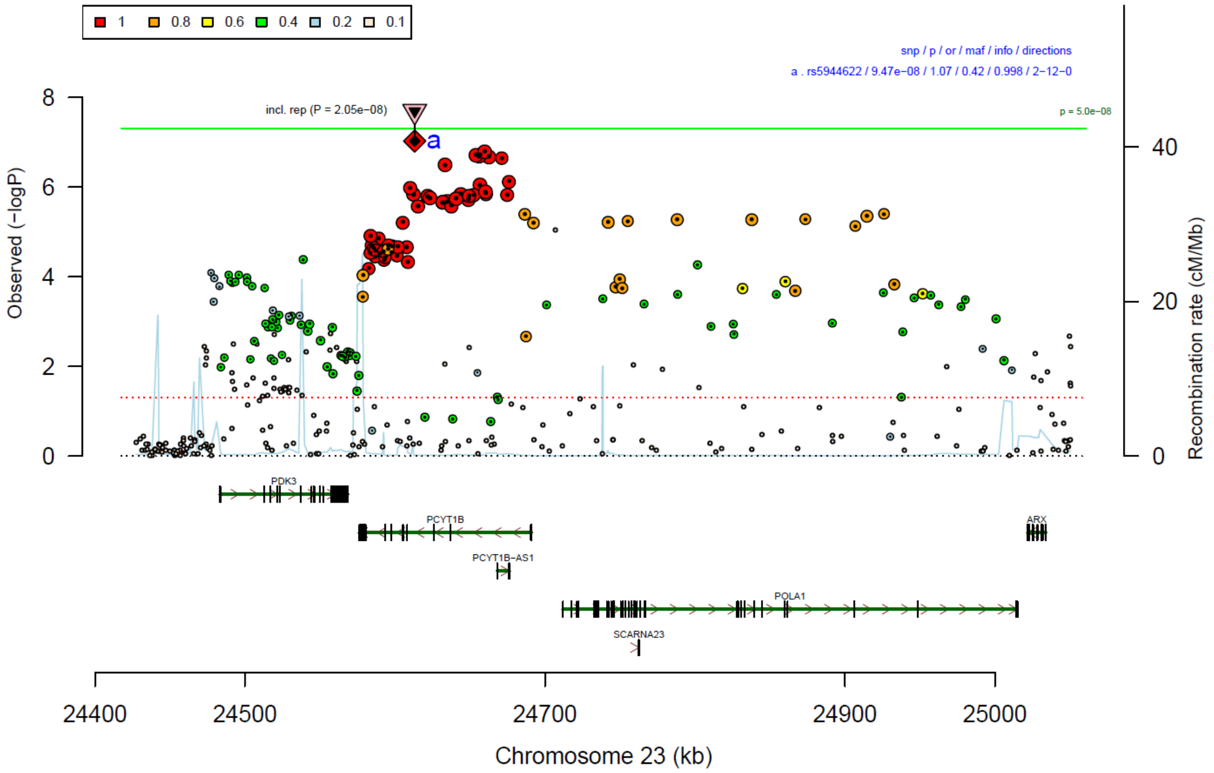


**Figure S23: region plot of rs5944622**

The lead SNP is indicated in diamond shape, and the linkage disequilibrium of the other SNPs ($r^2$) with the index SNP is represented by the colors. The downward-pointing triangle indicated the significance in the combined meta-analysis (discovery & replication). The green line indicates genome-wide significance ($P < 5*10^{-8}$).

| | | | | | |
|---|---|---|---|---|---|
| **rs5944622** | **T/C** | | **23:24613189** | | |
|  | ------+-----+-- | het_P: | 0.2926 | het_I: | 14.7 |
| | **p_value** | **f_ca(n)** | **f_co(n)** | **ln(OR)** | **STDerr** |
| BioVU | 0.0377 | 0.576(315) | 0.640(3150) | −0.123 | 0.059 |
| Danish | 0.00914 | 0.583(2379) | 0.583(334766) | −0.073 | 0.028 |
| Decode | 0.181 | 0.597(294) | 0.597(259612) | −0.132 | 0.099 |
| EstBb | 0.854 | 0.578(351) | 0.581(1755) | −0.014 | 0.077 |
| FinGen | 0.0121 | 0.546(3812) | 0.562(332850) | −0.052 | 0.021 |
| GLAD | 0.000761 | 0.603(1327) | 0.603(6703) | −0.155 | 0.046 |
| HUNT | 0.454 | 0.588(295) | 0.588(42535) | 0.051 | 0.068 |
| McBb | 0.0304 | 0.530(293) | 0.577(46196) | −0.161 | 0.074 |
| UKBb | 0.0648 | 0.538(182) | 0.592(1820) | −0.182 | 0.098 |
| borge | 0.19 | 0.435(1285) | 0.428(1330) | −0.075 | 0.0576 |
| boro1 | 0.546 | 0.472(57) | 0.446(700) | −0.145 | 0.2397 |
| boro2 | 0.251 | 0.461(64) | 0.361(270) | −0.256 | 0.223 |
| borsm | 0.427 | 0.355(306) | 0.376(759) | 0.075 | 0.094 |
| boma3 | 0.523 | 0.431(993) | 0.424(1539) | −0.041 | 0.065 |
| **META** | **9.47e−08** | **0.570(11947)** | **0.580(1033598)** | **−0.07** | **0.0132** |
| Replication (AllofUs) | 0.0749 | 0.582(639) | 0.582(107315) | −0.085 | 0.0477835 |
| **META (including replication)** | **2.05e−08** | **0.571(12586)** | **0.580(1140913)** | **−0.071** | **0.0127** |

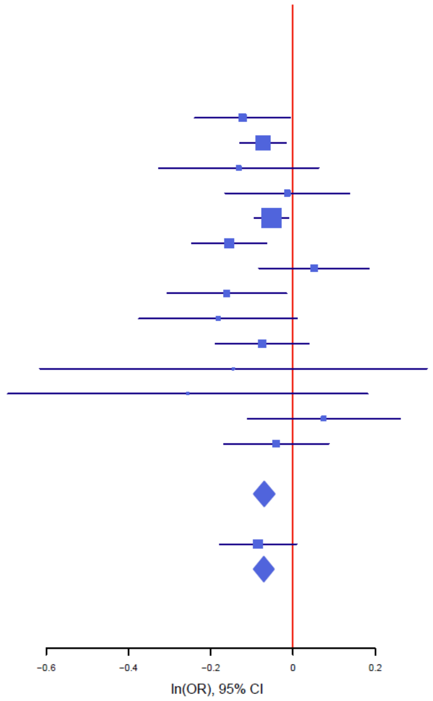


#### Figure S24: forest plot of rs5944622

The plot shows the size and direction of the SNP's effect in each analyzed cohort. Additionally, cohort-specific information and meta-analytic information on the SNP (imputed vs. genotyped SNP (ngt), info score (info), p value, frequency in cases and controls (f_ca(n), f_co(n)) effect size (ln(OR)) and standard error (STDerr)) are given in the table.

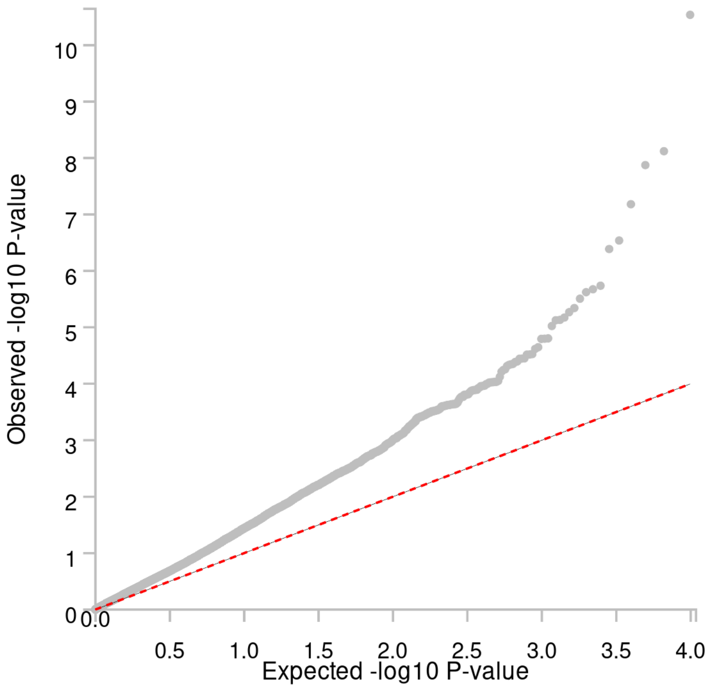


**Figure S25: Quantile–Quantile plot of the gene-based analysis (Ncases = 12,339, Ncontrols = 1,041,717)**

Observed –log10 *P-values* are shown on the y-axis, and expected –log10 *P-values* are shown on the x-axis.

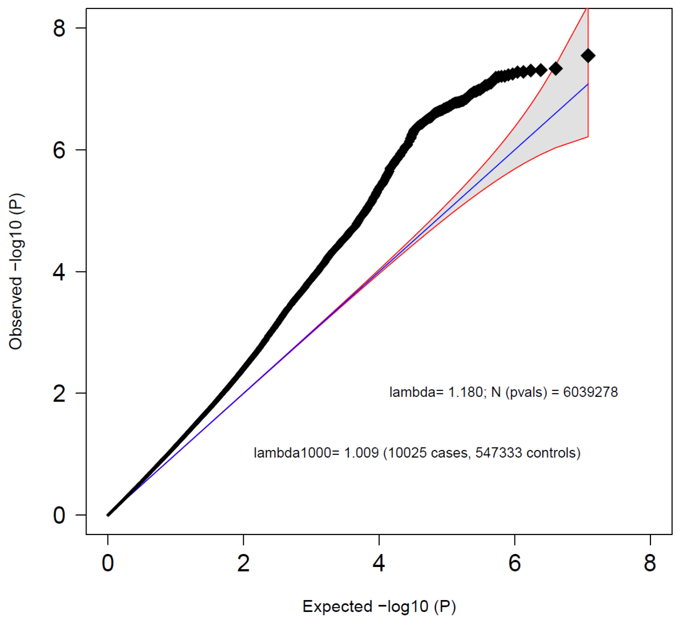


**Figure S26: Female subset - Quantile–Quantile plot of the GWAS meta-analysis of BPD (Ncases = 10,025, Ncontrols = 547,333)**

Observed –log10 *P-values* are shown on the y-axis, and expected –log10 *P-values* are shown on the x-axis. Lambda values were calculated based on the associations of the autosomal variants. The 95% confidence interval of expected P-values under the null hypothesis is indicated by the shaded region.

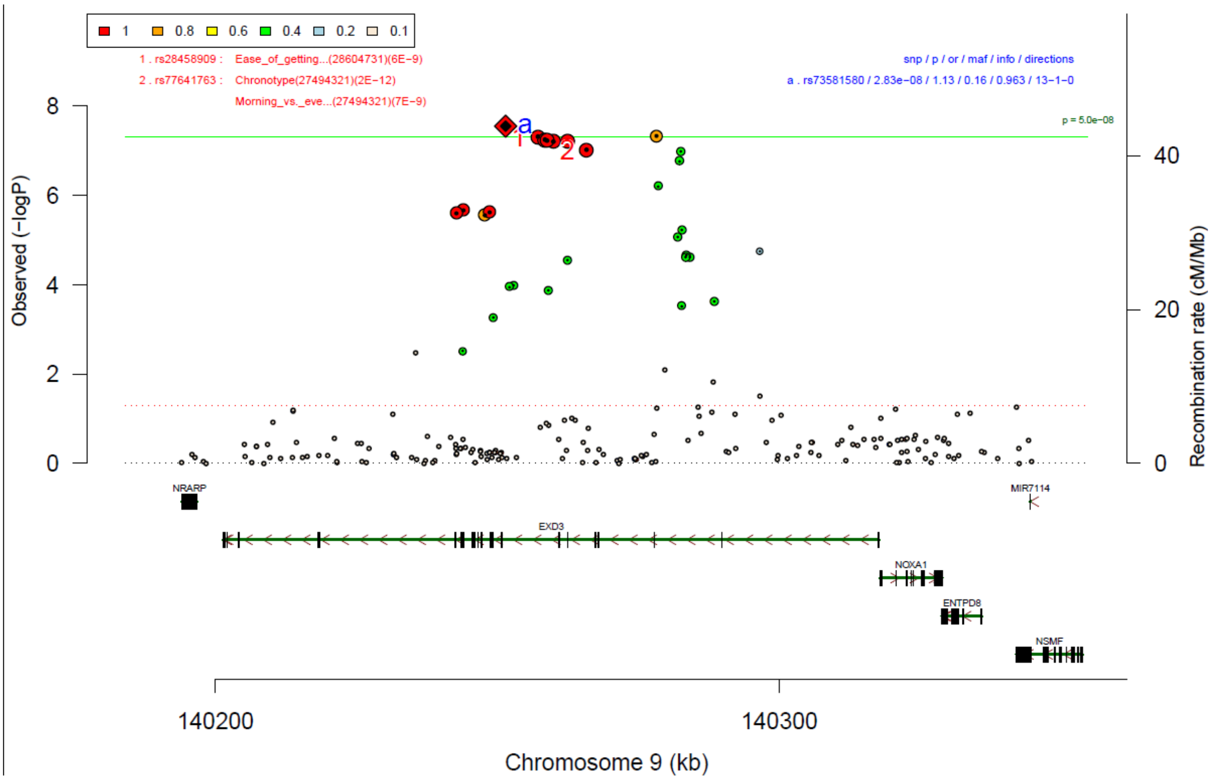


**Figure S27: Female subset - region plot of rs73581580**

The lead SNP is indicated in diamond shape, and the linkage disequilibrium of the other SNPs ($r^2$) with the index SNP is represented by the colors. The green line indicates genome-wide significance ($P < 5*10^{-8}$).

| rs73581580 | G/A | | 9:140251458 | | |
|---|---|---|---|---|---|
| | +++−++++++++++ | het_P: | 0.7482 | het_I: | 0.0 |
| | **p_value** | **f_ca(n)** | **f_co(n)** | **ln(OR)** | **STDerr** |
| BioVU (females) | 0.824 | 0.870(276) | 0.871(2758) | −0.029 | 0.134 |
| Danish (females) | 0.00227 | 0.852(1943) | 0.852(176216) | −0.142 | 0.046 |
| Decode (females) | 0.937 | 0.865(241) | 0.865(120968) | −0.012 | 0.151 |
| EstBb (females) | 0.742 | 0.864(243) | 0.859(1215) | 0.049 | 0.148 |
| FinGen (females) | 0.00415 | 0.777(2875) | 0.794(184845) | −0.092 | 0.032 |
| GLAD (females) | 0.332 | 0.883(1174) | 0.883(3521) | −0.092 | 0.094 |
| HUNT (females) | 0.153 | 0.870(230) | 0.870(20225) | −0.197 | 0.138 |
| McBb (females) | 0.296 | 0.859(216) | 0.874(26976) | −0.144 | 0.137 |
| MoBa (females) | 0.042 | 0.825(330) | 0.859(6610) | −0.268 | 0.1319 |
| borge (females) | 0.0176 | 0.869(1092) | 0.893(803) | −0.259 | 0.109 |
| boro1 (females) | 0.94 | 0.880(51) | 0.867(313) | −0.026 | 0.3481 |
| boro2 (females) | 0.313 | 0.809(55) | 0.837(147) | −0.306 | 0.3032 |
| borsm (females) | 0.0485 | 0.887(262) | 0.916(591) | −0.355 | 0.1797 |
| boma3 (females) | 0.0639 | 0.857(909) | 0.884(865) | −0.208 | 0.1124 |
| **META** | **2.83e−08** | **0.838(9780)** | **0.837(543713)** | **−0.122** | **0.0219** |

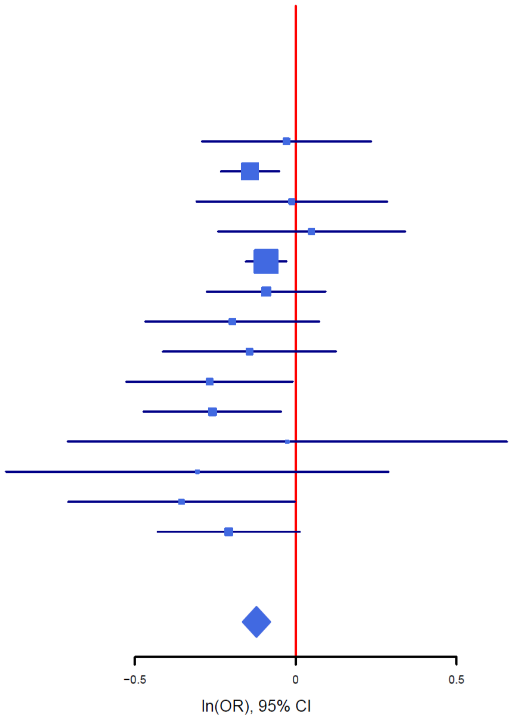


#### Figure S28: Female subset - forest plot of rs73581580

The plot shows the size and direction of the SNP's effect in each analyzed cohort. Additionally, cohort-specific information and meta-analytic information on the SNP (imputed vs. genotyped SNP (ngt), info score (info), p value, frequency in cases and controls (f_ca(n), f_co(n)) effect size (ln(OR)) and standard error (STDerr)) are given in the table.

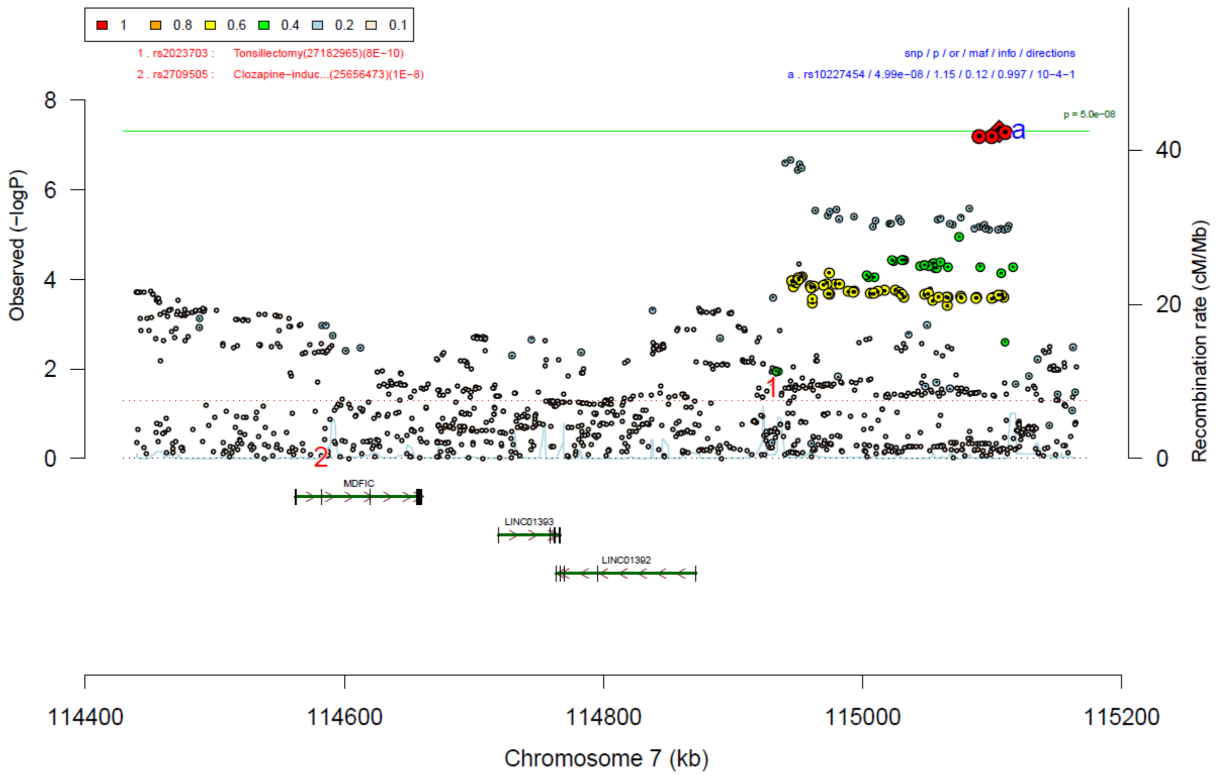


**Figure S29: Female subset - region plot of rs10227454**

The lead SNP is indicated in diamond shape, and the linkage disequilibrium of the other SNPs ($r^2$) with the index SNP is represented by the colors. The green line indicates genome-wide significance ($P < 5*10^{-8}$).

| rs10227454 | G/T | | 7:115105520 | | |
|---|---|---|---|---|---|
| | ?++−++++++−−+−+ | het_P: | 0.03931 | het_I: | 44.0 |
| | **p_value** | **f_ca(n)** | **f_co(n)** | **ln(OR)** | **STDerr** |
| Danish (females) | 0.000211 | 0.890(1943) | 0.890(176216) | −0.188 | 0.051 |
| Decode (females) | 0.58 | 0.863(241) | 0.863(120968) | −0.08 | 0.146 |
| EstBb (females) | 0.0517 | 0.940(243) | 0.914(1215) | 0.36 | 0.185 |
| FinGen (females) | 0.00195 | 0.863(2875) | 0.879(184845) | −0.12 | 0.039 |
| GLAD (females) | 0.00716 | 0.877(1174) | 0.877(3521) | −0.248 | 0.092 |
| HUNT (females) | 0.00462 | 0.879(230) | 0.879(20225) | −0.411 | 0.145 |
| McBb (females) | 0.261 | 0.867(216) | 0.885(26976) | −0.159 | 0.142 |
| MoBa (females) | 0.0311 | 0.852(330) | 0.877(6610) | −0.248 | 0.1149 |
| UKBb (females) | 0.943 | 0.880(128) | 0.882(1280) | −0.015 | 0.209 |
| borge (females) | 0.51 | 0.901(1092) | 0.896(803) | 0.078 | 0.1182 |
| boro1 (females) | 0.3 | 0.902(51) | 0.869(313) | 0.407 | 0.3932 |
| boro2 (females) | 0.177 | 0.891(55) | 0.847(147) | 0.504 | 0.3739 |
| borsm (females) | 0.76 | 0.906(262) | 0.913(591) | −0.058 | 0.188 |
| boma3 (females) | 0.27 | 0.898(909) | 0.904(865) | −0.14 | 0.1271 |
| **META** | **4.99e−08** | **0.881(9749)** | **0.879(544575)** | **−0.136** | **0.0249** |

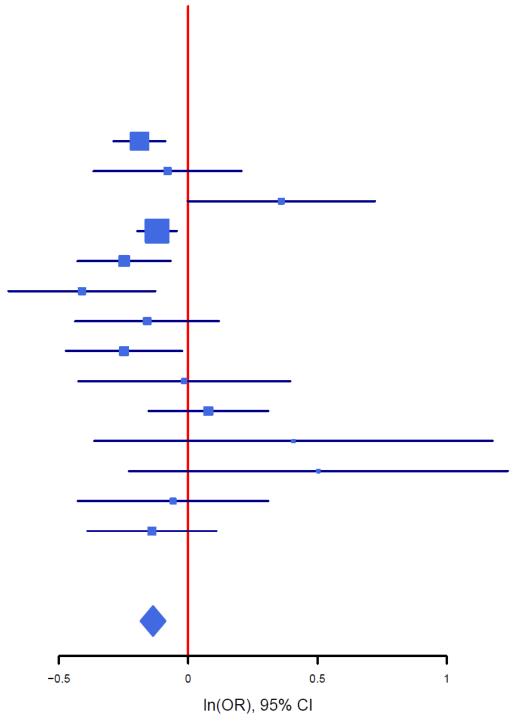


#### Figure S30: Female subset - forest plot of rs10227454

The plot shows the size and direction of the SNP's effect in each analyzed cohort. Additionally, cohort-specific information and meta-analytic information on the SNP (imputed vs. genotyped SNP (ngt), info score (info), p value, frequency in cases and controls (f_ca(n), f_co(n)) effect size (ln(OR)) and standard error (STDerr)) are given in the table.

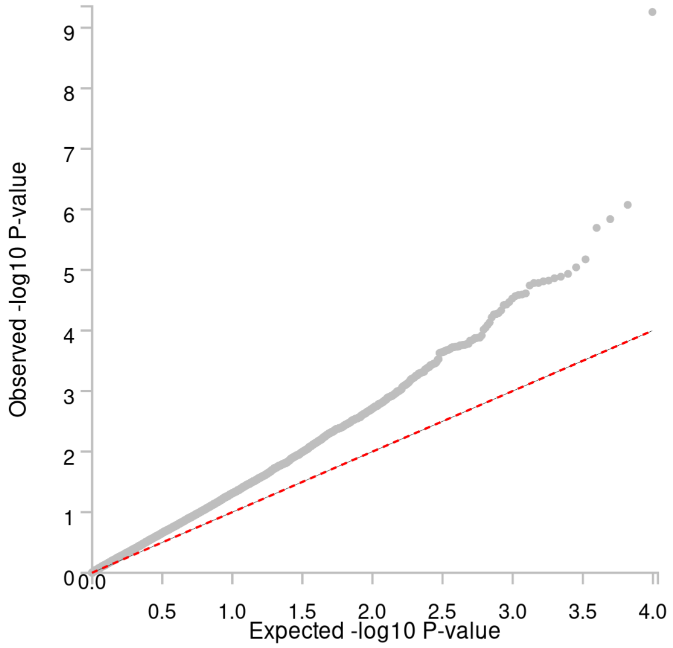


**Figure S31: Female subset - Quantile–Quantile plot of the gene-based analysis (Ncases = 10,025, Ncontrols = 547,333)**

Observed –log10 *P-values* are shown on the y-axis, and expected –log10 *P-values* are shown on the x-axis.

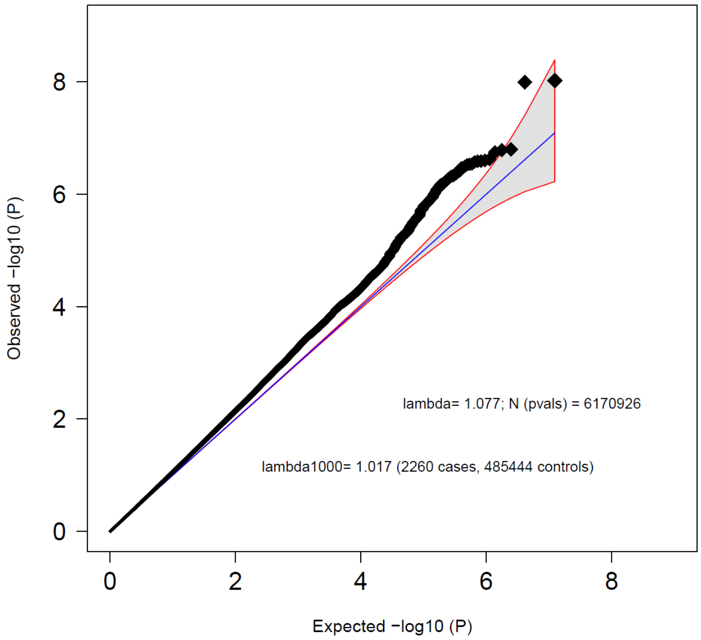


**Figure S32: Male subset - Quantile–Quantile plot of the GWAS meta-analysis of BPD (Ncases = 2260, Ncontrols = 485,444)**

Observed –log10 *P-values* are shown on the y-axis, and expected –log10 *P-values* are shown on the x-axis. Lambda values were calculated based on the associations of the autosomal variants. The 95% confidence interval of expected P-values under the null hypothesis is indicated by the shaded region.

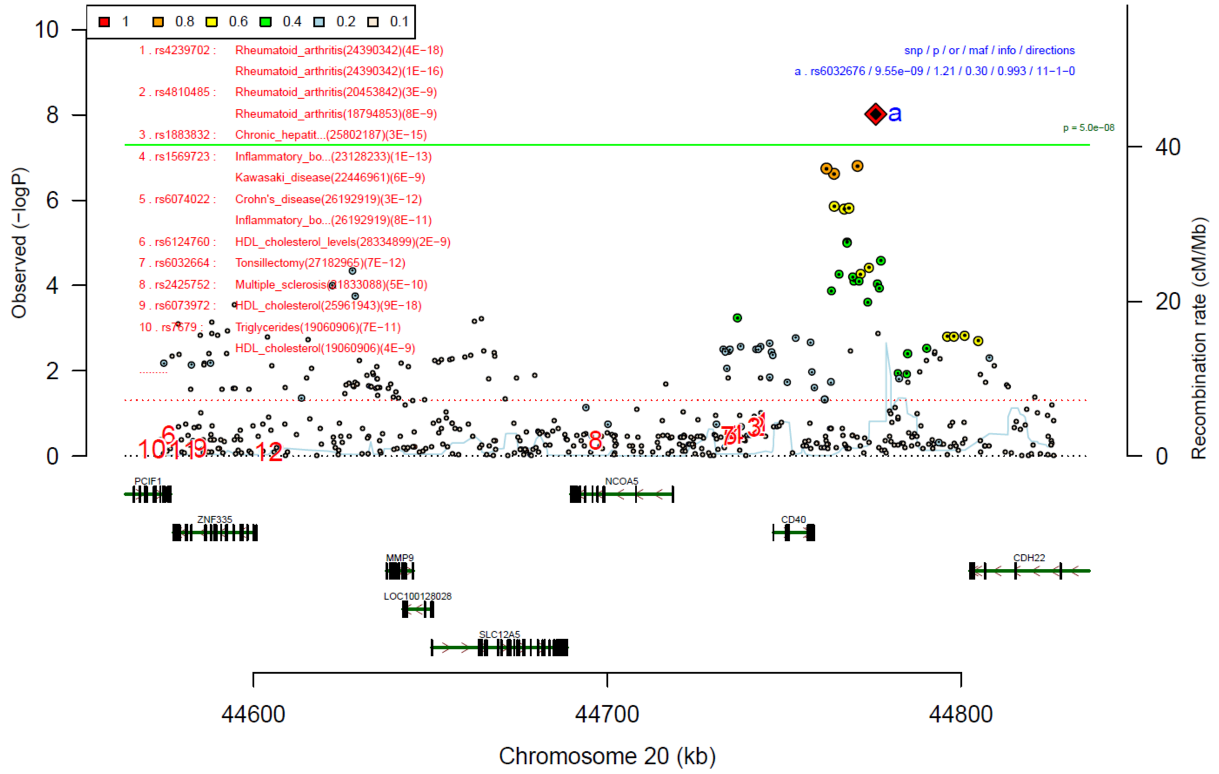


#### Figure S33: Male subset - region plot of rs6032676

The lead SNP is indicated in diamond shape, and the linkage disequilibrium of the other SNPs ($r^2$) with the index SNP is represented by the colors. The green line indicates genome-wide significance ($P < 5*10^{-8}$).

| rs6032676 | G/C | | 20:44775863 | | |
|---|---|---|---|---|---|
| | +++++++++++− | het_P: | 0.6669 | het_I: | 0.0 |
| | **p_value** | **f_ca(n)** | **f_co(n)** | **ln(OR)** | **STDerr** |
| Danish (males) | 3.42e−06 | 0.681(436) | 0.681(158105) | −0.329 | 0.071 |
| Decode (males) | 0.257 | 0.713(53) | 0.713(131053) | −0.281 | 0.248 |
| EstBb (males) | 0.377 | 0.648(108) | 0.684(540) | −0.15 | 0.169 |
| FinGen (males) | 0.00104 | 0.667(937) | 0.702(148005) | −0.161 | 0.049 |
| GLAD (males) | 0.51 | 0.700(153) | 0.700(3182) | −0.092 | 0.14 |
| HUNT (males) | 0.198 | 0.691(65) | 0.691(22310) | −0.245 | 0.19 |
| McBb (males) | 0.194 | 0.648(77) | 0.691(19220) | −0.221 | 0.17 |
| MoBa (males) | 0.664 | 0.697(56) | 0.690(1120) | −0.097 | 0.2234 |
| UKBb (males) | 0.086 | 0.644(54) | 0.701(540) | −0.4 | 0.233 |
| borge (males) | 0.833 | 0.278(193) | 0.288(527) | −0.03 | 0.1438 |
| borsm (males) | 0.705 | 0.285(44) | 0.273(168) | −0.108 | 0.2867 |
| boma3 (males) | 0.893 | 0.333(84) | 0.300(674) | 0.026 | 0.1919 |
| **META** | **9.55e−09** | **0.678(2260)** | **0.697(485444)** | **−0.191** | **0.0332** |

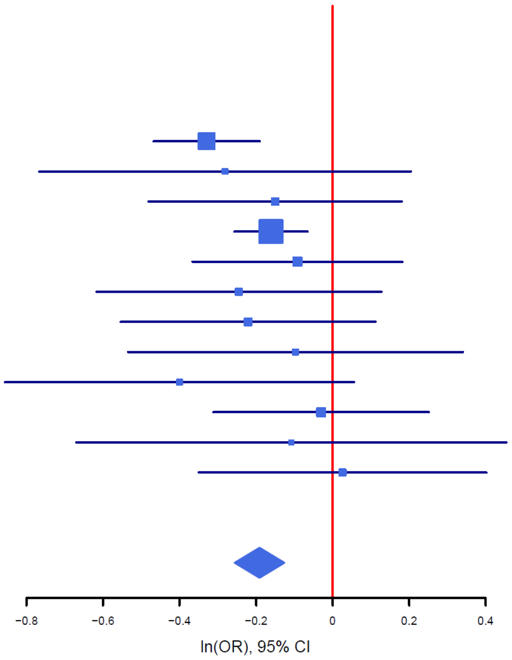


#### Figure S34: Male subset - forest plot of rs6032676

The plot shows the size and direction of the SNP’s effect in each analyzed cohort. Additionally, cohort-specific information and meta-analytic information on the SNP (imputed vs. genotyped SNP (ngt), info score (info), p value, frequency in cases and controls (f_ca(n), f_co(n)) effect size (ln(OR)) and standard error (STDerr)) are given in the table.

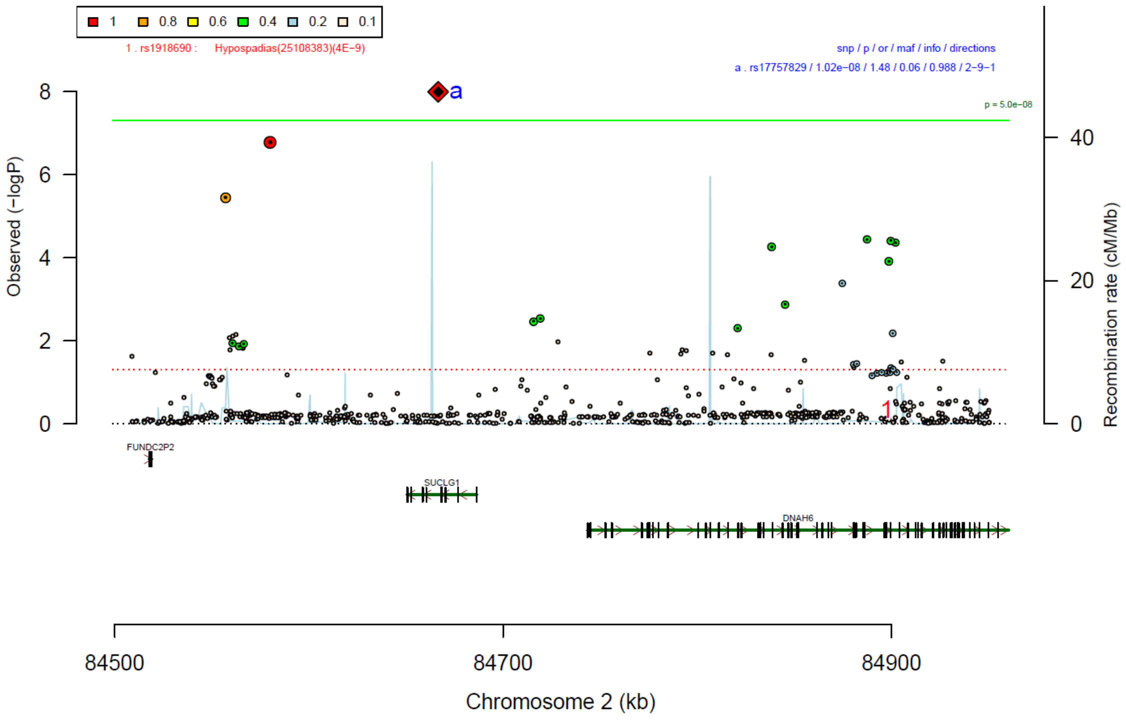


**Figure S35: Male subset - region plot of rs17757829**

The lead SNP is indicated in diamond shape, and the linkage disequilibrium of the other SNPs ($r^2$) with the index SNP is represented by the colors. The green line indicates genome-wide significance ($P < 5*10^{-8}$).

| rs17757829 | T/C | | 2:84666634 | | |
|---|---|---|---|---|---|
| | −−−−−−−?−++− | het_P: | 0.08505 | het_I: | 39.6 |
| | **p_value** | **f_ca(n)** | **f_co(n)** | **ln(OR)** | **STDerr** |
| Danish (males) | 0.000384 | 0.938(436) | 0.938(158105) | −0.446 | 0.126 |
| Decode (males) | 0.794 | 0.934(53) | 0.934(131053) | −0.12 | 0.458 |
| EstBb (males) | 0.00209 | 0.920(108) | 0.967(540) | −1.217 | 0.396 |
| FinGen (males) | 0.0685 | 0.956(937) | 0.963(148005) | −0.206 | 0.113 |
| GLAD (males) | 0.0631 | 0.936(153) | 0.936(3182) | −0.48 | 0.258 |
| HUNT (males) | 0.0633 | 0.930(65) | 0.930(22310) | −0.671 | 0.361 |
| McBb (males) | 0.00491 | 0.891(77) | 0.941(19220) | −0.724 | 0.257 |
| UKBb (males) | 0.0226 | 0.907(54) | 0.952(540) | −0.87 | 0.382 |
| borge (males) | 0.639 | 0.947(193) | 0.946(527) | 0.16 | 0.3406 |
| borsm (males) | 0.639 | 0.926(44) | 0.917(168) | 0.254 | 0.5407 |
| boma3 (males) | 0.0615 | 0.934(84) | 0.959(674) | −0.82 | 0.4383 |
| **META** | **1.02e−08** | **0.942(2204)** | **0.944(484324)** | **−0.391** | **0.0683** |

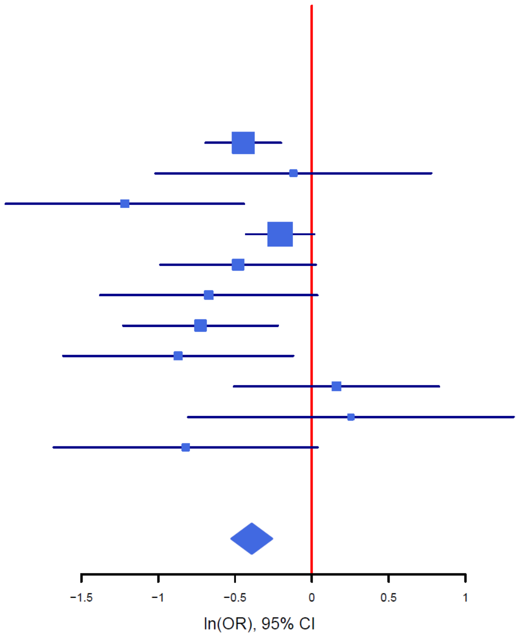


#### Figure S36: Male subset - forest plot of rs17757829

The plot shows the size and direction of the SNP’s effect in each analyzed cohort. Additionally, cohort-specific information and meta-analytic information on the SNP (imputed vs. genotyped SNP (ngt), info score (info), p value, frequency in cases and controls (f_ca(n), f_co(n)) effect size (ln(OR)) and standard error (STDerr)) are given in the table.

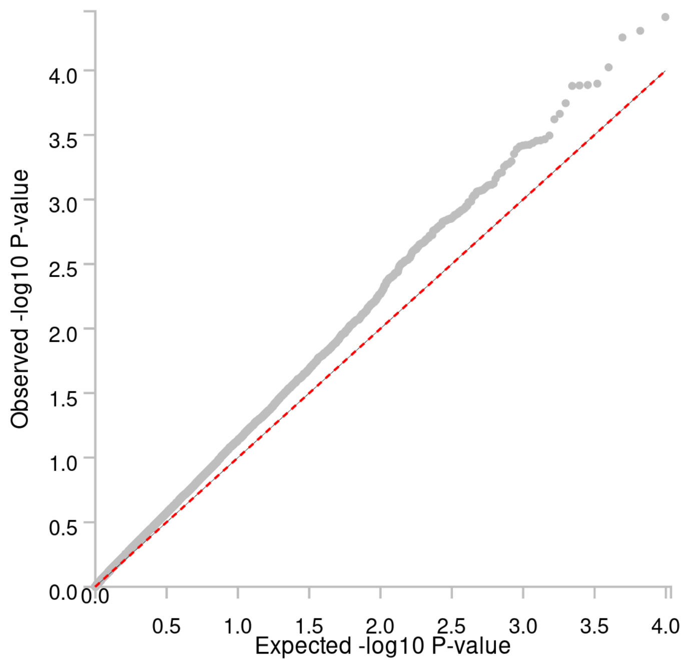


**Figure S37: Male subset - Quantile–Quantile plot of the gene-based analysis (Ncases = 2260, Ncontrols = 485,444)**

Observed –log10 *P-values* are shown on the y-axis, and expected –log10 *P-values* are shown on the x-axis.

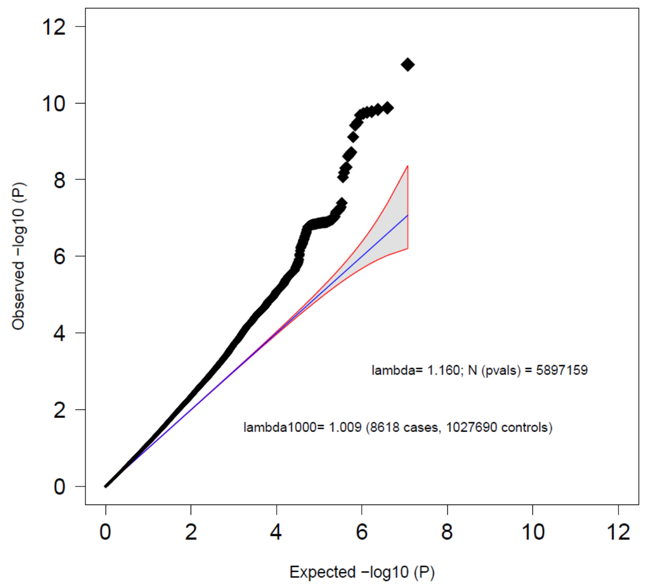

**Figure S38: Sensitivity analysis - Quantile–Quantile plot of the GWAS meta-analysis of BPD excluding subjects with schizophrenia or bipolar disorder (Ncases = 8,618, Ncontrols = 1,027,690)**

Observed –log10 *P-values* are shown on the y-axis, and expected –log10 *P-values* are shown on the x-axis. Lambda values were calculated based on the associations of the autosomal variants. The 95% confidence interval of expected P-values under the null hypothesis is indicated by the shaded region.

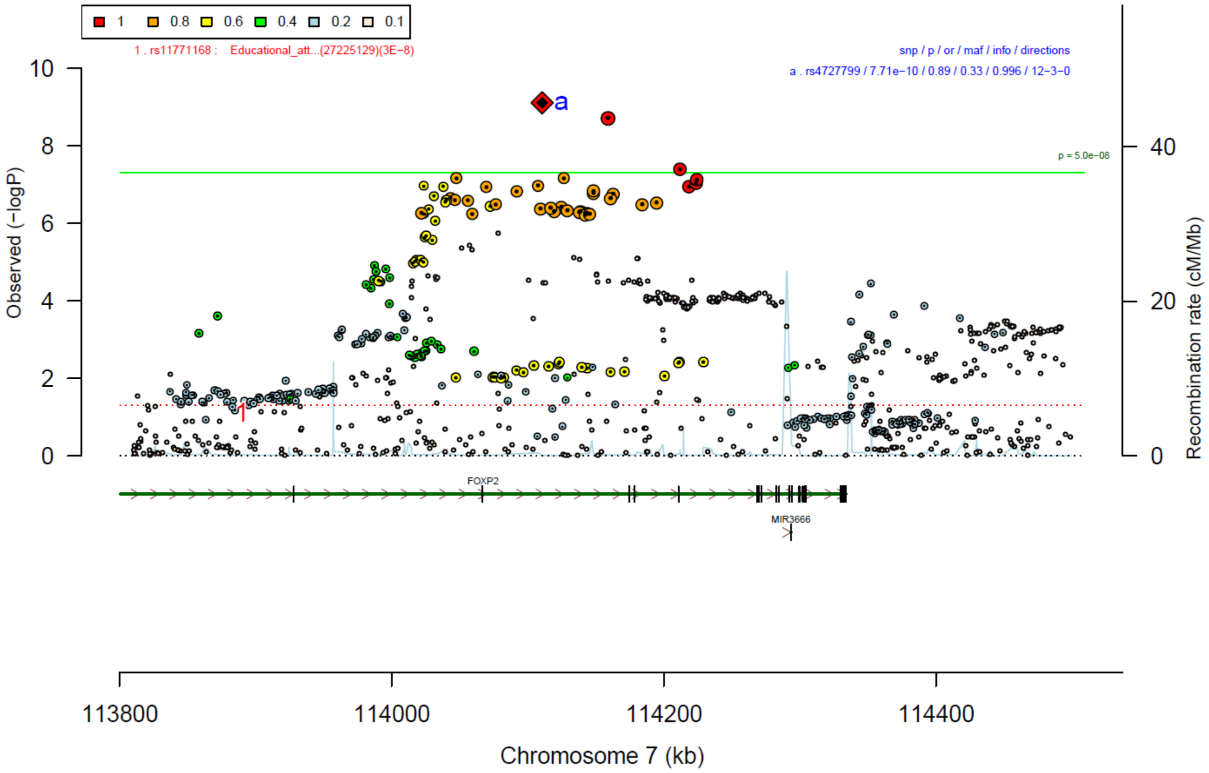


**Figure S39: Sensitivity analysis - region plot of rs4727799**

The lead SNP is indicated in diamond shape, and the linkage disequilibrium of the other SNPs ($r^2$) with the index SNP is represented by the colors. The green line indicates genome-wide significance ($P < 5*10^{-8}$).

| rs4727799 | T/C | | 7:114110568 | | |
|---|---|---|---|---|---|
| | −++−+++++−+++++ | het_P: | 0.01704 | het_I: | 48.9 |
| | **p_value** | **f_ca(n)** | **f_co(n)** | **ln(OR)** | **STDerr** |
| BioVU (ex.BIP.SCZ) | 0.636 | 0.646(99) | 0.660(990) | −0.076 | 0.161 |
| Danish (ex.BIP.SCZ) | 0.000296 | 0.655(1553) | 0.655(334766) | 0.142 | 0.039 |
| Decode (ex.BIP_SCZ) | 0.0574 | 0.684(194) | 0.684(261778) | 0.247 | 0.13 |
| EstBb (ex.BIP_SCZ) | 0.777 | 0.624(243) | 0.630(1215) | −0.029 | 0.103 |
| FinGen (ex.BIP_SCZ) | 0.051 | 0.689(2083) | 0.677(324515) | 0.066 | 0.034 |
| GLAD (ex.BIP_SCZ) | 0.226 | 0.645(1019) | 0.645(6703) | 0.075 | 0.062 |
| HUNT (ex.BIP_SCZ) | 0.0121 | 0.686(224) | 0.686(42535) | 0.258 | 0.103 |
| McBb (ex.BIP_SCZ) | 0.0455 | 0.697(151) | 0.642(44788) | 0.252 | 0.126 |
| MoBa (ex.BIP_SCZ) | 0.208 | 0.722(233) | 0.671(4660) | 0.119 | 0.0947 |
| UKBb (ex.BIP_SCZ) | 0.157 | 0.592(114) | 0.635(1140) | −0.207 | 0.146 |
| borge (ex.BIP_SCZ) | 0.098 | 0.366(1285) | 0.392(1332) | 0.1 | 0.0603 |
| boro1 (ex.BIP_SCZ) | 0.0707 | 0.254(57) | 0.331(700) | 0.428 | 0.2367 |
| boro2 (ex.BIP_SCZ) | 0.000916 | 0.219(64) | 0.363(270) | 0.857 | 0.2585 |
| borsm (ex.BIP_SCZ) | 0.122 | 0.399(306) | 0.433(759) | 0.155 | 0.1 |
| boma3 (ex.BIP_SCZ) | 0.00198 | 0.353(993) | 0.397(1539) | 0.205 | 0.0663 |
| **META** | **7.71e−10** | **0.660(8618)** | **0.670(1027690)** | **0.114** | **0.0185** |

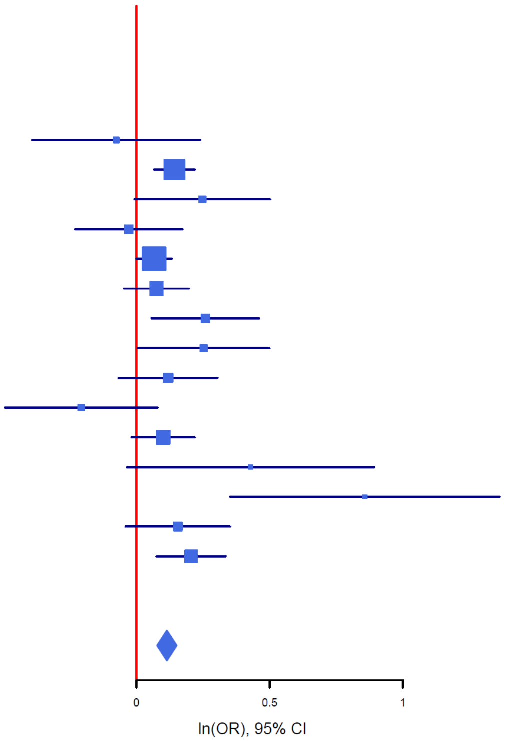


**Figure S40: Sensitivity analysis - forest plot of rs4727799**

The plot shows the size and direction of the SNP's effect in each analyzed cohort. Additionally, cohort-specific information and meta-analytic information on the SNP (imputed vs. genotyped SNP (ngt), info score (info), p value, frequency in cases and controls (f_ca(n), f_co(n)) effect size (ln(OR)) and standard error (STDerr)) are given in the table.

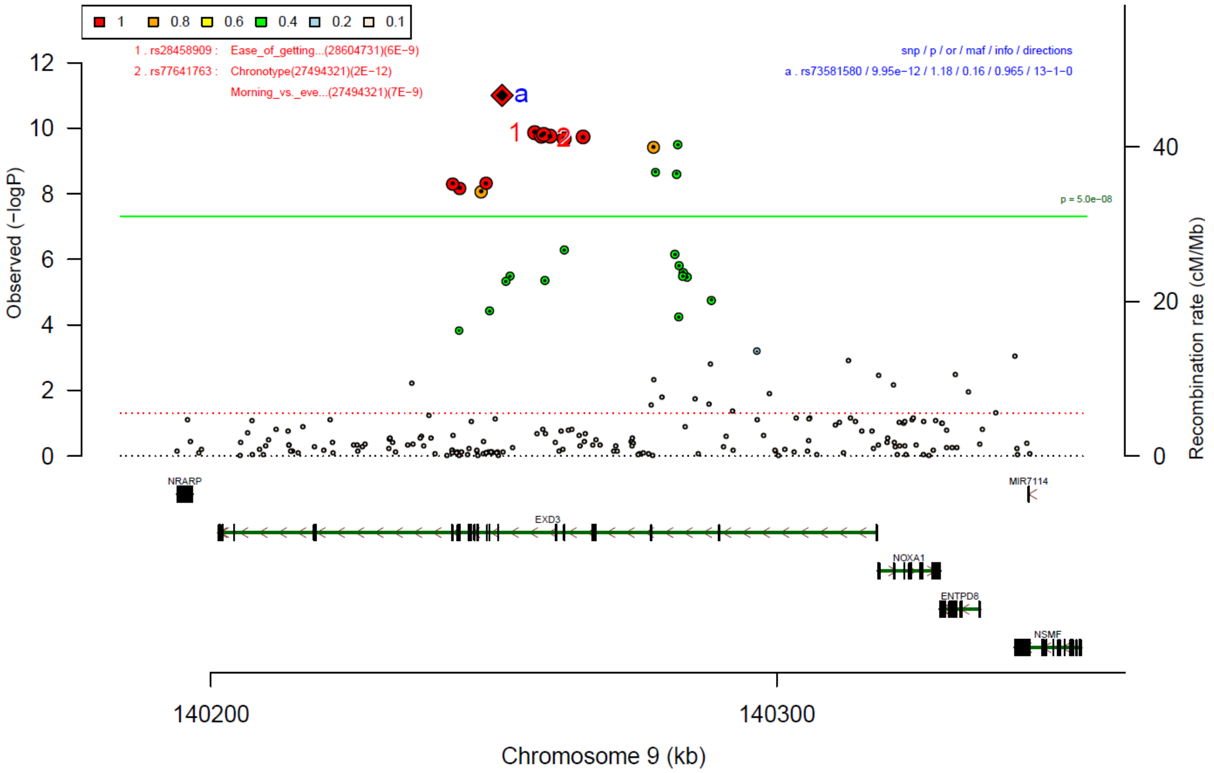


**Figure S41: Sensitivity analysis - region plot of rs73581580**

The lead SNP is indicated in diamond shape, and the linkage disequilibrium of the other SNPs ($r^2$) with the index SNP is represented by the colors. The green line indicates genome-wide significance ($P < 5*10^{-8}$).

| rs73581580 | G/A | | 9:140251458 | | |
|---|---|---|---|---|---|
| | +++−+++++++++++ | het_P: | 0.4493 | het_I: | 0.0 |
| | **p_value** | **f_ca(n)** | **f_co(n)** | **ln(OR)** | **STDerr** |
| BioVU (ex.BIP.SCZ) | 0.346 | 0.854(99) | 0.879(990) | −0.209 | 0.222 |
| Danish (ex.BIP.SCZ) | 0.00035 | 0.852(1553) | 0.852(334766) | −0.183 | 0.051 |
| Decode (ex.BIP_SCZ) | 0.582 | 0.865(194) | 0.865(261778) | −0.092 | 0.168 |
| EstBb (ex.BIP_SCZ) | 0.0908 | 0.885(243) | 0.857(1215) | 0.245 | 0.145 |
| FinGen (ex.BIP_SCZ) | 3.74e−06 | 0.763(2083) | 0.795(324515) | −0.17 | 0.037 |
| GLAD (ex.BIP_SCZ) | 0.584 | 0.883(1019) | 0.883(6703) | −0.049 | 0.089 |
| HUNT (ex.BIP_SCZ) | 0.0167 | 0.870(224) | 0.870(42535) | −0.337 | 0.141 |
| McBb (ex.BIP_SCZ) | 0.328 | 0.858(151) | 0.875(44788) | −0.159 | 0.163 |
| MoBa (ex.BIP_SCZ) | 0.102 | 0.833(233) | 0.858(4660) | −0.242 | 0.148 |
| borge (ex.BIP_SCZ) | 0.0707 | 0.868(1285) | 0.887(1332) | −0.161 | 0.0893 |
| boro1 (ex.BIP_SCZ) | 0.509 | 0.866(57) | 0.875(700) | −0.202 | 0.306 |
| boro2 (ex.BIP_SCZ) | 0.139 | 0.828(64) | 0.870(270) | −0.407 | 0.2752 |
| borsm (ex.BIP_SCZ) | 0.0985 | 0.890(306) | 0.913(759) | −0.271 | 0.1639 |
| boma3 (ex.BIP_SCZ) | 0.0748 | 0.858(993) | 0.884(1539) | −0.17 | 0.0952 |
| **META** | **9.95e−12** | **0.840(8452)** | **0.839(1025510)** | **−0.162** | **0.0237** |

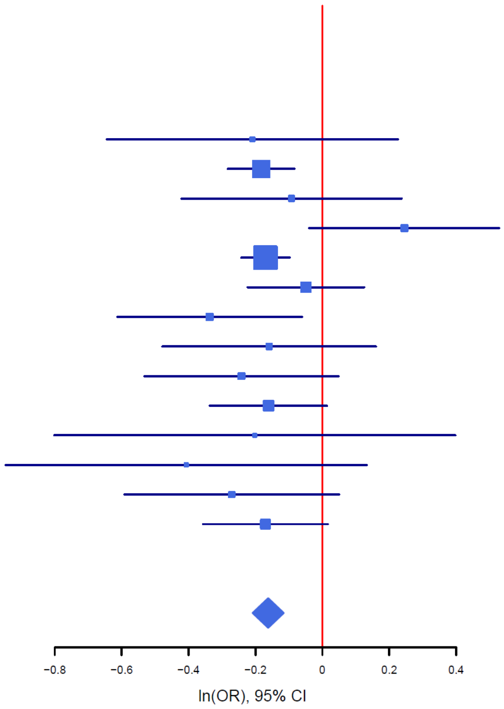


**Figure S42: Sensitivity analysis - forest plot of rs73581580**

The plot shows the size and direction of the SNP’s effect in each analyzed cohort. Additionally, cohort-specific information and meta-analytic information on the SNP (imputed vs. genotyped SNP (ngt), info score (info), p value, frequency in cases and controls (f_ca(n), f_co(n)) effect size (ln(OR)) and standard error (STDerr)) are given in the table.

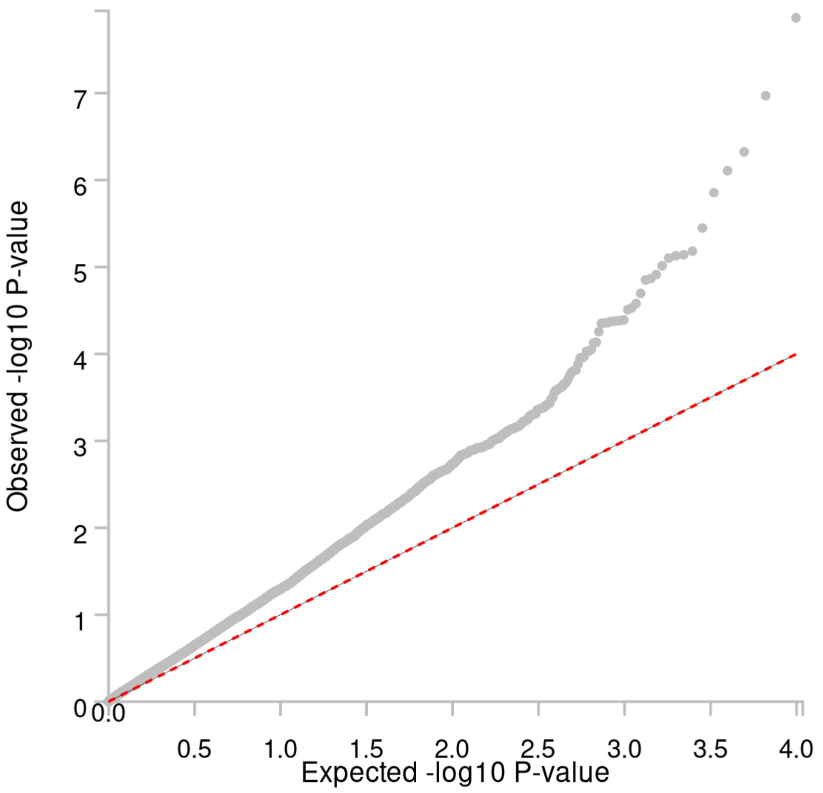


**Figure S43: Sensitivity analysis - Quantile–Quantile plot of the gene-based analysis of subset excluding subjects with schizophrenia or bipolar disorder (Ncases = 8,618, Ncontrols = 1,027,690)**

Observed –log10 *P-values* are shown on the y-axis, and expected –log10 *P-values* are shown on the x-axis.

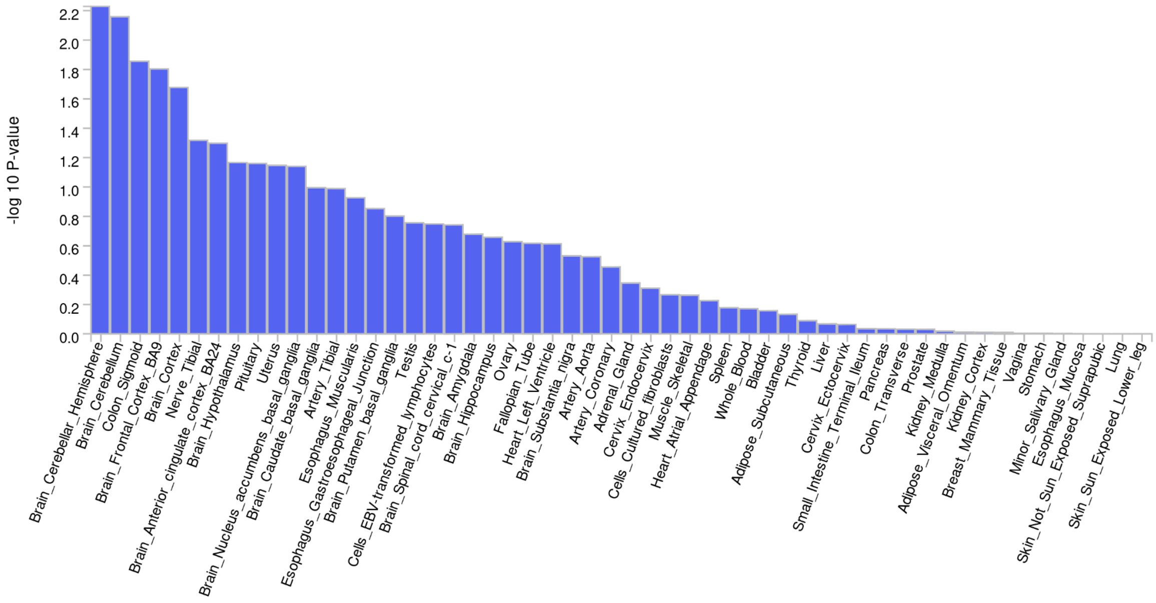


**Figure S44: GTEx tissue enrichment expression of genes associated with BPD as implemented in FUMA**

-log10 p-values are indicated on the y-axis.

---

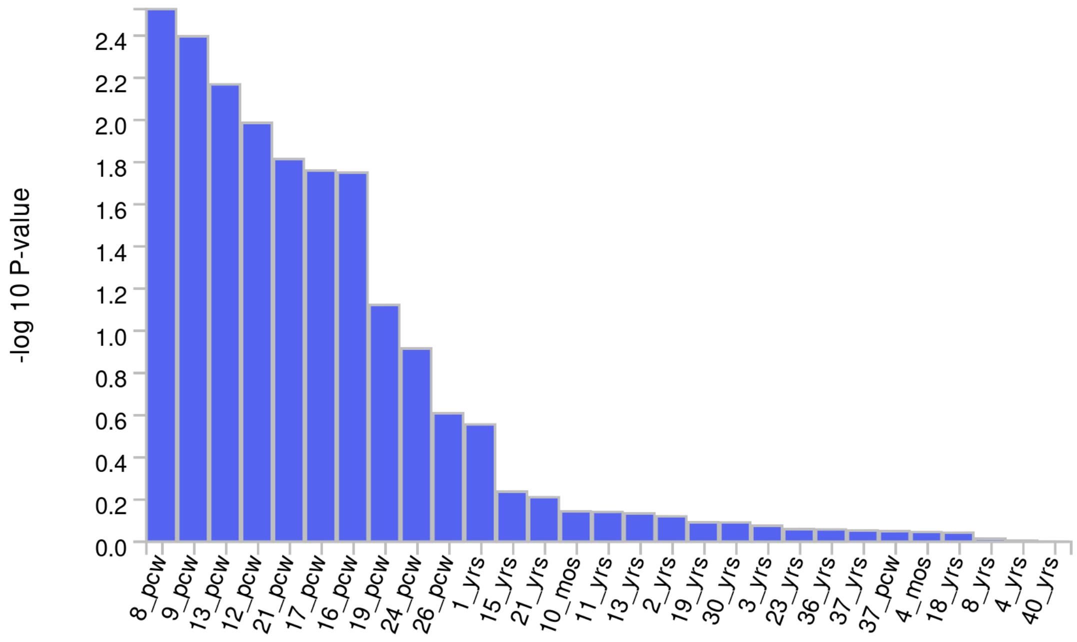


**Figure S45: BrainSpan age groups: enrichment of expression of genes associated with BPD as implemented in FUMA**

-log10 p-values are indicated on the y-axis.; pcw = post-conceptional Weeks; mos = months; yrs = years

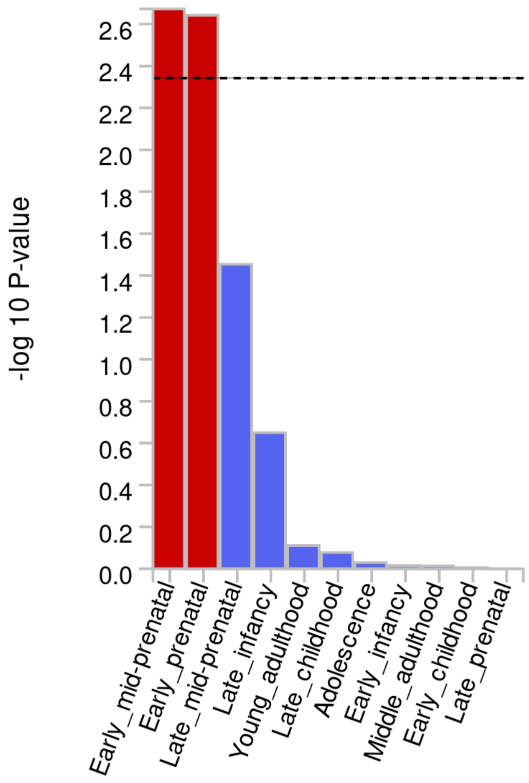


**Figure S46: BrainSpan developmental stages: enrichment of expression of genes associated with BPD as implemented in FUMA**

-log10 p-values are indicated on the y-axis. Red bars indicate significance after correction for multiple testing.

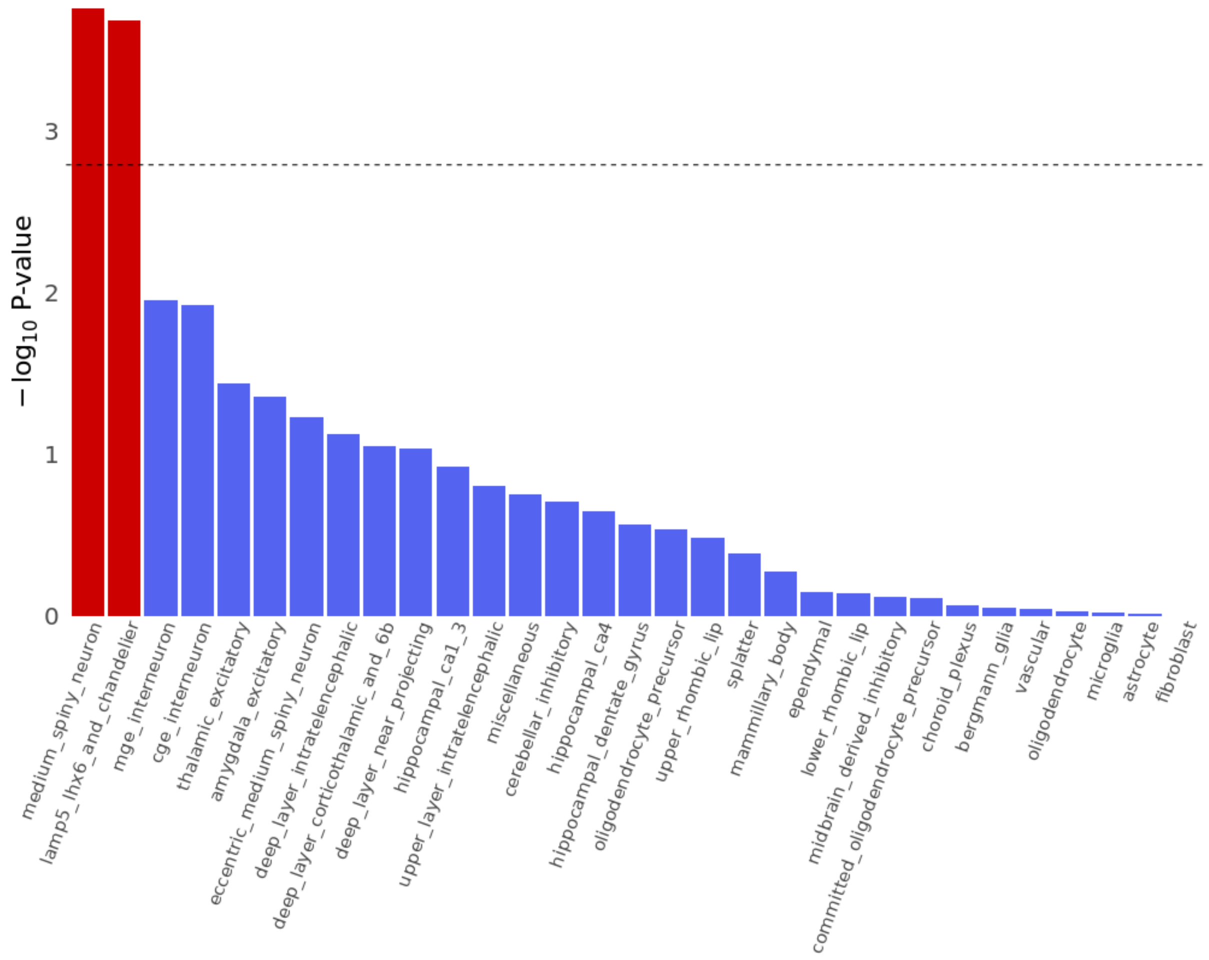


**Figure S47: SNP-h² enrichment for BPD on the supercluster-level based on Human Brain Atlas single-nucleus RNA sequencing data**

-log10 p-values are indicated on the y-axis. Red bars indicate significance after correction for multiple testing.

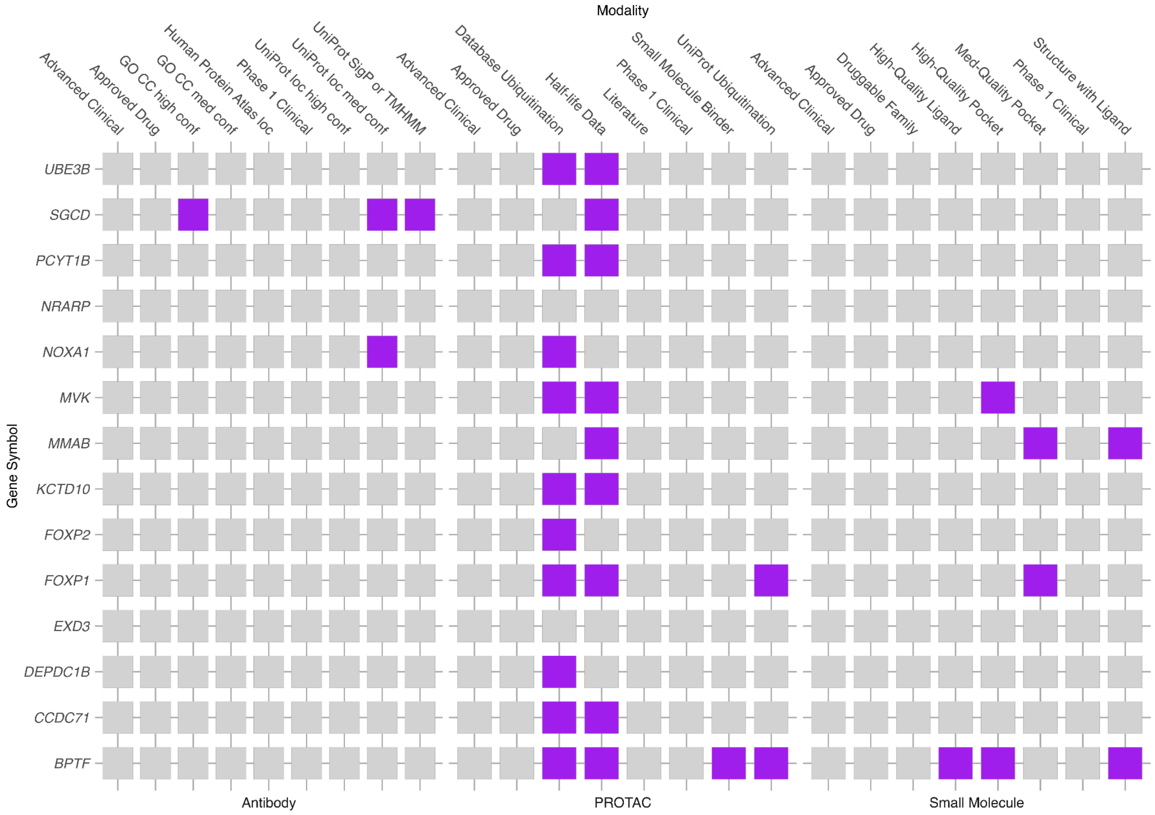


**Figure S48: Drug target analysis**

Only prioritized genes with available information in the database are displayed.

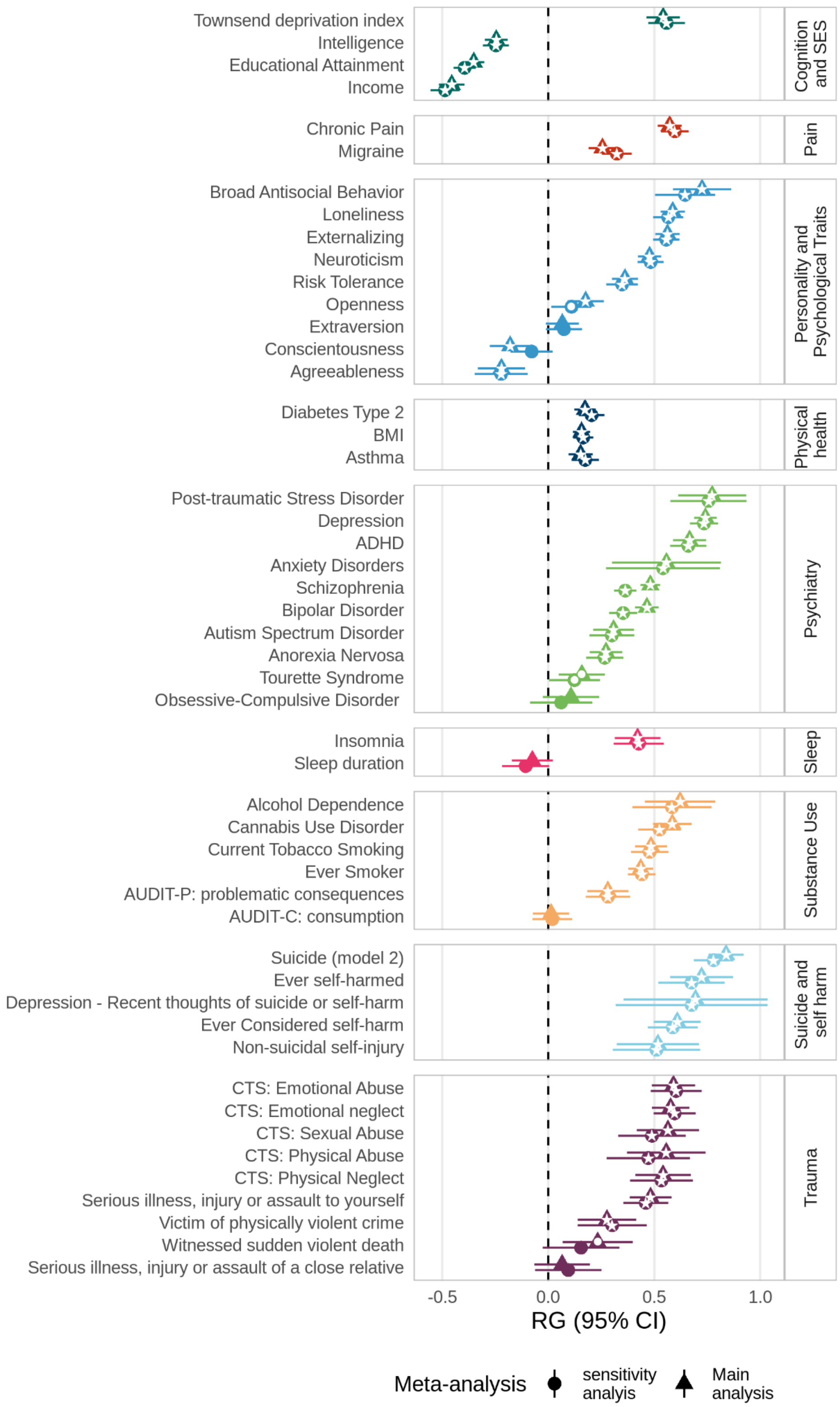


**Figure S49: Genetic correlations of BPD with other phenotypes**

Genetic correlations of BPD main analysis ($N_{cases}$=12,339, $N_{controls}$=1,041,717, triangle shape) and BPD sensitivity analysis (excl BIP/SCZ; $N_{cases}$= 8,618, $N_{controls}$= 1,027,690, sphere shape) with 51 disorders and traits; within each category, disorders and traits are sorted by their genetic correlation. • *p*<0.05, * *p*<0.00098 (0.05/51 tested correlations); 95% CI = 95% confidence interval, AUDIT = Alcohol Use Disorders Identification Test, BMI = body mass index, CTQ = childhood trauma questionnaire, RG = genetic correlation.
